# A Multimodal Sleep Foundation Model for Disease Prediction

**DOI:** 10.1101/2025.02.04.25321675

**Authors:** Rahul Thapa, Magnus Ruud Kjaer, Bryan He, Ian Covert, Hyatt Moore, Umaer Hanif, Gauri Ganjoo, M. Brandon Westover, Poul Jennum, Andreas Brink-Kjaer, Emmanuel Mignot, James Zou

## Abstract

Sleep is a fundamental biological process with broad implications for physical and mental health, yet its complex relationship with disease remains poorly understood. Polysomnography (PSG), the gold standard for sleep analysis, captures rich physiological signals but remains underutilized due to challenges in standardization, generalizability, and multimodal integration. To address these limitations, we developed SleepFM, a multimodal sleep foundation model trained with a novel contrastive learning approach that accommodates multiple PSG montages—the specific arrangements of electrodes and sensors used to record physiological signals during sleep. Trained on a curated dataset of over 585,000 hours of PSG recordings from approximately 65,000 participants across multiple cohorts, SleepFM produces latent sleep representations that capture the physiological and temporal structure of sleep and enable accurate prediction of future disease risk. SleepFM achieved a C-Index of at least 0.75 (Bonferroni-corrected p < 0.01) for 130 conditions, including all-cause mortality (C-Index: 0.84), dementia (0.85), myocardial infarction (0.81), heart failure (0.80), chronic kidney disease (0.79), stroke (0.78), and atrial fibrillation (0.78). Moreover, the model demonstrates strong transfer learning performance on a dataset from the Sleep Heart Health Study (SHHS), a dataset that was excluded from pretraining, and performs competitively with specialized sleep-staging models such as U-Sleep and YASA on common sleep analysis tasks, achieving mean F1 scores of 0.70–0.78 for sleep staging and accuracies of 0.69 and 0.87 for classifying sleep apnea severity and presence. This work shows that foundation models can extract clinically meaningful features from multi-modal sleep recordings, enabling scalable, label-efficient analysis and disease prediction.

## Introduction

Sleep is a complex process characterized by intricate interactions across physiological systems, including brain, heart, respiratory, and muscle activity [1]. Polysomnography (PSG), the gold standard for sleep evaluation, captures these interactions through recordings of multiple modalities including brain activity signals (BAS, including EEG and EOG), electrocardiography (ECG), electromyography (EMG), and respiratory signals [2].

Sleep disorders affect millions of people and are increasingly recognized as indicators of and contributors to various health conditions [3]. Sleep disturbances often precede the clinical onset of numerous conditions, such as psychiatric disorders [4], neurodegenerative diseases [5] and cardiovascular disorders [6]. These associations highlight an important role sleep plays in maintaining overall health and underscores its predictive potential across a wide spectrum of diseases. However, most existing studies have focused on identifying links between sleep and specific diseases using isolated metrics or manual annotations, leaving much of the complexity of sleep physiology, as captured in PSG, underutilized.

Recent advances in deep learning have enabled the use of PSG’s multimodal data for tasks ranging from sleep staging and apnea detection to predicting conditions like atrial fibrillation, biological aging, and narcolepsy [3, 7, 8, 9, 10]. Despite this progress, current approaches face key limitations: they focus on individual outcomes, depend on supervised learning with expert-labeled data, and are trained on relatively small datasets (2,500–15,913 recordings) [3, 7, 9, 10, 11]. Manual annotations are time-consuming and prone to inter-rater variability, making scaling difficult. Moreover, existing models lack flexibility across recording environments, generalize poorly across cohorts, and often fail to exploit the richness of multimodal sleep signals. There remains a need for robust, generalizable architectures and systematic evaluation of sleep’s predictive value across a broad range of health conditions.

Foundation models have emerged as a transformative approach in machine learning, enabling robust representation learning from large-scale, unlabeled data [12]. By leveraging self-supervised learning, these models can be fine-tuned efficiently for diverse applications. In biomedicine, foundation models have demonstrated remarkable capabilities in analyzing complex, heterogeneous datasets, driving advances in disease prediction, patient stratification, and therapeutic discovery [13, 14]. Their ability to extract meaningful patterns from large-scale data has addressed many challenges associated with the diverse and high-dimensional nature of clinical datasets.

Despite these successes, their application to sleep remains limited. Sleep data, particularly from PSG, presents unique challenges due to its complexity and variability, including differences in the number and types of recording channels across clinical cohorts. Most sleep studies have narrowly focused on sleep-specific outcomes, constraining the broader potential of foundation models for disease prediction. In preliminary work, we explored self-supervised learning on PSG data in a smaller cohort of participants [11]. While this effort highlighted the potential of foundation models for analyzing sleep data, it primarily targeted sleep-specific outcomes and lacked the flexibility to accommodate the diverse configurations of PSG recordings. These limitations emphasize the need for models that can generalize across heterogeneous datasets and systematically uncover sleep’s role in predicting a wider range of diseases.

In this paper, we present SleepFM, a foundation model trained on over 585,000 hours of PSG data from 65,000+ participants. SleepFM captures the diverse information present in multimodal sleep recordings—integrating EEG, ECG, EMG, and respiratory signals. Its channel-agnostic architecture enables joint learning across multiple modalities, producing representations that generalize across environments. We also introduce a new leave-one-out contrastive learning algorithm that aligns information across modalities during pretraining while remaining resilient to missing or heterogeneous channels during inference. Our model uses 5–25 times more data than previously trained supervised sleep [3, 7, 9, 10] or biosignal models [15, 16].

Inspired by phenome-wide association studies (PheWAS) [17], we examined whether sleep characteristics, as captured by SleepFM, can predict the onset of a wide range of diseases. Leveraging EHR disease codes, we develop a framework to systematically explore predictive associations between multimodal sleep and diverse health conditions.

## Dataset and SleepFM architecture

We describe our dataset and training procedures in detail in Methods section. Briefly, we used PSG data from four primary cohorts: Stanford Sleep Clinic (SSC) [11], BioSerenity [18, 19], the Multi-Ethnic Study of Atherosclerosis (MESA) [20, 21], and the Outcomes of Sleep Disorders in Older Men (MrOS) [20, 22]. SSC includes 35,052 studies from participants aged 1–100 years; BioSerenity adds 18,900 studies from individuals aged 7–90; MESA and MrOS contribute 2,237 and 3,930 PSGs, respectively, from older adults. Together, these cohorts span 65,000 participants and over 585,000 hours of sleep recordings. We further evaluated generalization using the Sleep Heart Health Study (SHHS) [20, 23], a multi-center dataset of 6,441 adults aged 40+, held out from pretraining and used solely for transfer learning. Dataset distributions post-filtering are shown in Table 1. Demographics for SSC and BioSerenity appear in Extended Table 1 and Extended Table 2, while details for SHHS, MrOS, and MESA are available in their respective publications.

**Table 1:** Distribution of polysomnography recordings across cohorts and data splits. The model was first pre-trained on SSC, BioSerenity, MESA, and MROS data, following which these same recordings were used for task-specific fine-tuning. The SHHS dataset is reserved exclusively for evaluating transfer learning capabilities and was used only during fine-tuning not during pre-training. The temporal test set consists of SSC recordings from 2020 onwards, used to evaluate model robustness to temporal distribution shifts. Dashes (-) indicate that no data is available for that split.

| Split | SSC | BioSerenity | MESA | MROS | SHHS | Total |
| --- | --- | --- | --- | --- | --- | --- |
| Train | 24,137 | 18,869 | 1,747 | 3,340 | 3,291 | 51,384 |
| Validation | 764 | 100 | 10 | 18 | 500 | 1,392 |
| Test | 5,019 | — | 150 | 286 | 2,000 | 7,455 |
| Temporal Test | 5,132 | — | — | — | — | 5,132 |
| Total | 35,052 | 18,969 | 1,907 | 3,644 | 5,791 | 65,363 |

Our pre-processing pipeline begins by resampling all signals to 128 Hz for consistency across cohorts. Signals are then segmented into 5-second windows, which serve as the model’s fundamental input tokens. The architecture includes 1D convolutional layers for feature extraction, followed by channel-agnostic attention pooling to address variability in channel number and order across cohorts. A transformer block captures temporal dependencies over a 5-minute context window. During pre-training, we use a multimodal contrastive learning objective to align representations across all modalities. The model’s robustness stems from its channel-agnostic design, enabling it to accommodate missing channels, varying channel counts, and heterogeneous signal types.

For downstream tasks, we leverage the pre-trained model’s embeddings through lightweight fine-tuning. The token embeddings from different modalities are pooled again and processed by a 2-layer Long Short-Term Memory (LSTM) network before passing through task-specific output heads. For patient-level prediction tasks (e.g., disease prediction), an additional temporal pooling layer before the output layer compresses all token embeddings into a single 128-dimensional embedding.

To evaluate model performance across tasks, we use appropriate task-specific metrics. For classification tasks such as gender classification, we report AUROC and AUPRC; for sleep apnea classification we show confusion matrices and report accuracy; for age estimation, we use mean absolute error (MAE) and Pearson correlation. Sleep staging is evaluated using the F1 score, which is well-suited for class-imbalanced settings. For disease prediction, we report AUROC and Harrell’s concordance index (C-Index), a standard survival analysis metric that measures the proportion of correctly ranked risk pairs. All metrics range from 0 to 1, with higher values indicating better performance. Confidence intervals (95%) are computed using bootstrapping.

## SleepFM Supports Standard Sleep Analysis Tasks

After pre-training SleepFM, we assessed the general utility of its learned representations by fine-tuning on four common benchmark tasks: age estimation, gender classification, sleep stage classification, and sleep apnea classification. While these tasks are not the main focus of our work, they are useful validations showing that the model captures fundamental sleep patterns. For all tasks, we trained lightweight LSTM-based heads on top of the frozen multimodal embeddings derived from entire nights of PSG data.

For age estimation, we assessed the model’s ability to predict chronological age. Overall performance is shown in Extended Figure 1, with the model achieving a MAE of 7.33 years and a correlation coefficient of 0.88. Performance varied across age groups, with higher accuracy in pediatric and middle-aged participants and greater error in elderly adults, suggesting that age prediction is more challenging at the extremes of the age spectrum. Gender classification yielded an AUROC of 0.86 [0.85–0.87] and AUPRC of 0.90 [0.89–0.91]. For sleep stage classification, we fine-tuned a LSTM-based classifier to distinguish Wake, Stage 1, Stage 2, Stage 3, and REM using 5-second windows—a more granular resolution than the standard 30-second epochs, which has been shown to improve precision in certain conditions (e.g., narcolepsy [10]). As shown in Supplementary Figure 1, SleepFM performs well on Wake, Stage 2, and REM, with expected confusion in transitional stages like Stage 1—consistent with known human scoring variability. We report results across SSC, MESA, MrOS, and SHHS, where SleepFM achieves competitive performance compared to U-Sleep [7], YASA [24], GSSC [25], and STAGES [10], state-of-the-art sleep staging models, as shown in Extended Table 3 and Extended Table 4. Additionally, we compare SleepFM to three PhysioEx [26] models on the public datasets DCSM [27] and HMC [28] in a fully external validation setting, achieving an F1-score of 0.68 on DCSM—outperforming all models—and 0.55 on HMC (see Supplementary Table 1). While the source alone has little impact, using multiple datasets for pretraining and fine-tuning improves generalization, boosting macro F1 by around 0.1 (see Supplementary Tables 2, 3, and 4), consistent with prior work [26].

For sleep apnea classification, we performed patient-level severity classification to distinguish between four commonly used severity groups based on the apnea-hypopnea index (AHI): none (AHI *<* 5), mild (5 *≤* AHI *<* 15), moderate (15 *≤* AHI *<* 30), and severe (AHI *≥* 30). Across MESA, MrOs, and SHHS, we observe competitive performance, with a severity classification accuracy of 0.69 and a presence classification accuracy (none/mild vs. moderate/severe) of 0.87. The confusion matrix for apnea classification is shown in Figure 1.

**Figure 1:**
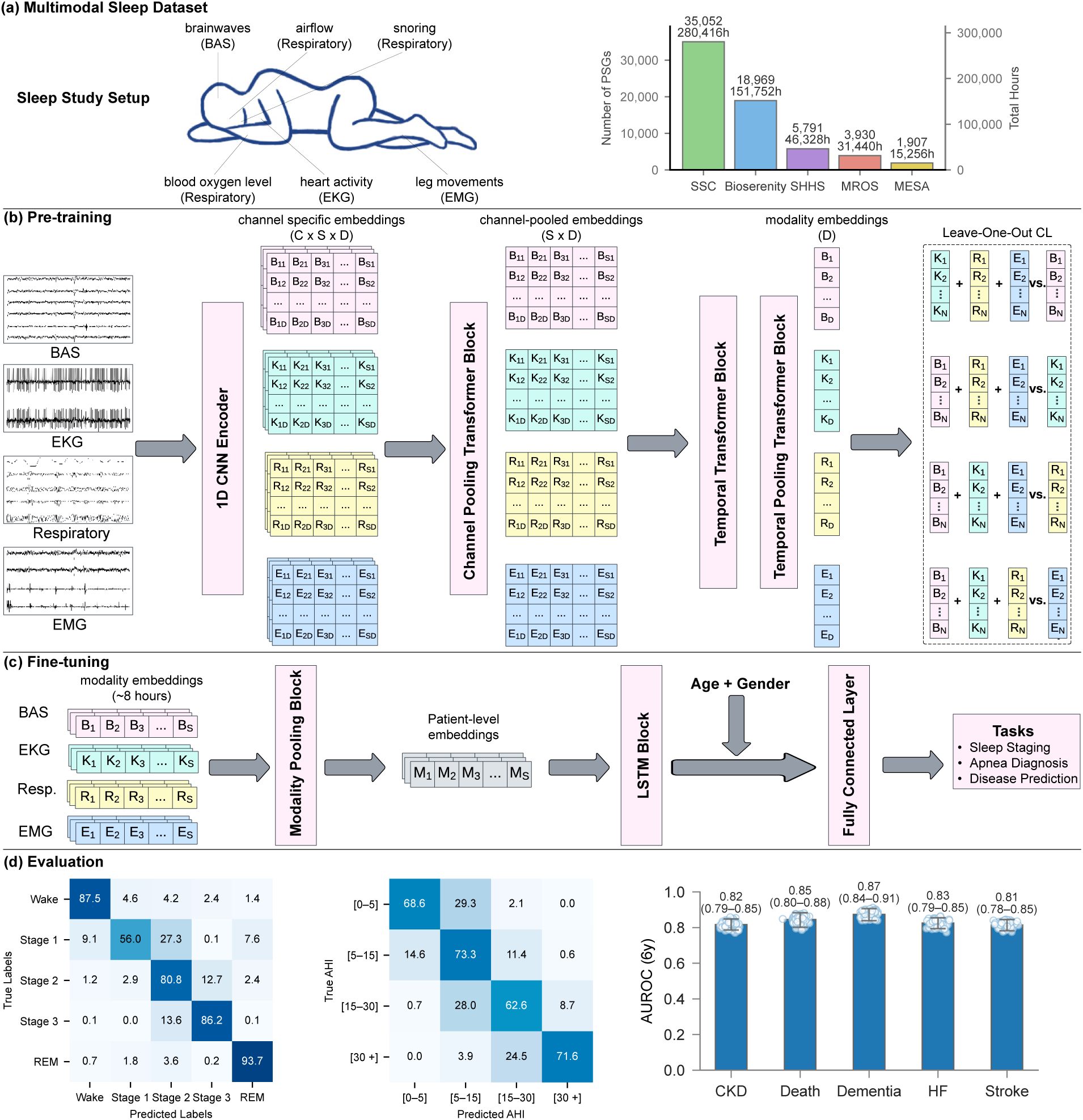
Overview of the SleepFM framework. (a) Polysomnography (PSG) setup and dataset statistics across multiple sleep centers. Bars show the number of independent PSG recordings (participants) per cohort and the corresponding total recording hours. (b) Multimodal contrastive pre-training: raw signals from each modality are encoded by a CNN, channel embeddings are pooled within modality, and a temporal transformer with temporal pooling yields sequence-level representations for leave-one-out contrastive learning (CL). *C*: channels, *S*: sequence length, *D*: embedding dimension. (c) Fine-tuning using frozen embeddings for downstream tasks (sleep staging, apnea detection, disease prediction). Eight hours of multimodal embeddings are aggregated to patient-level representations, concatenated with age and gender, and passed to an LSTM followed by a fully connected layer. (d) Evaluation across representative tasks and clinical applications. Left and middle, confusion matrices for sleep staging (SHHS) and AHI categories (SSC) shown as row-normalized percentages. Right, disease-prediction performance on the Stanford cohort (n=5,019 participants). Box plots summarize 1,000 patient-level bootstrap resamples: faint dots (individual bootstrap draws), and vertical line with end caps (95% bootstrap percentile CI). Numeric labels are means. The number of positive samples for each disease is as follows: CKD (354), Death (224), Dementia (221), HF (283), and Stroke (297). **Abbreviations:** BAS, brain activity signal; EKG, electrocardiogram; EMG, electromyogram; SSC, Stanford Sleep Cohort; SHHS, Sleep Heart Health Study; MROS, Outcomes of Sleep Disorders in Older Men; MESA, Multi-Ethnic Study of Atherosclerosis; CKD, chronic kidney disease; HF, heart failure.

To enable disease prediction, we paired SSC data with EHRs, extracting all diagnostic codes (ICD-9 and ICD-10) and their timestamps. These codes were mapped to phecodes—a hierarchical system of 1,868 disease categories designed for PheWAS [29]. Each phecode’s timestamp was defined as the earliest among its corresponding ICD codes. Positive cases were defined as patients whose first phecode instance occurred more than 7 days after the sleep study, avoiding trivial associations. We excluded phecodes with prevalence below 1.5% to ensure statistical power, resulting in 1,041 phecodes for evaluation. For model fine-tuning, we used a multi-label extension of the Cox proportional hazards (CoxPH) loss, averaging independent losses computed for each label.

Figure 2 illustrates SleepFM’s performance across disease categories on the test set. While performance varies across categories, SleepFM demonstrates strong results in several areas, including neoplasms, pregnancy complications, circulatory conditions, and mental disorders. Overall, 130 future diseases achieved a C-Index and AUROC of at least 0.75 on held-out participants (and Bonferroni-corrected *p <* 0.01), as summarized in Supplementary Table 5. AUROC was calculated using a six-year horizon, meaning a condition is considered positive if the patient develops the disease within six years of their PSG study. The six-year horizon for AUROC calculation was chosen to balance performance and account for both long-term and short-term conditions. Supplementary Figure 2 shows AUROC values across 1–6 year horizons for multiple conditions.

**Figure 2:**
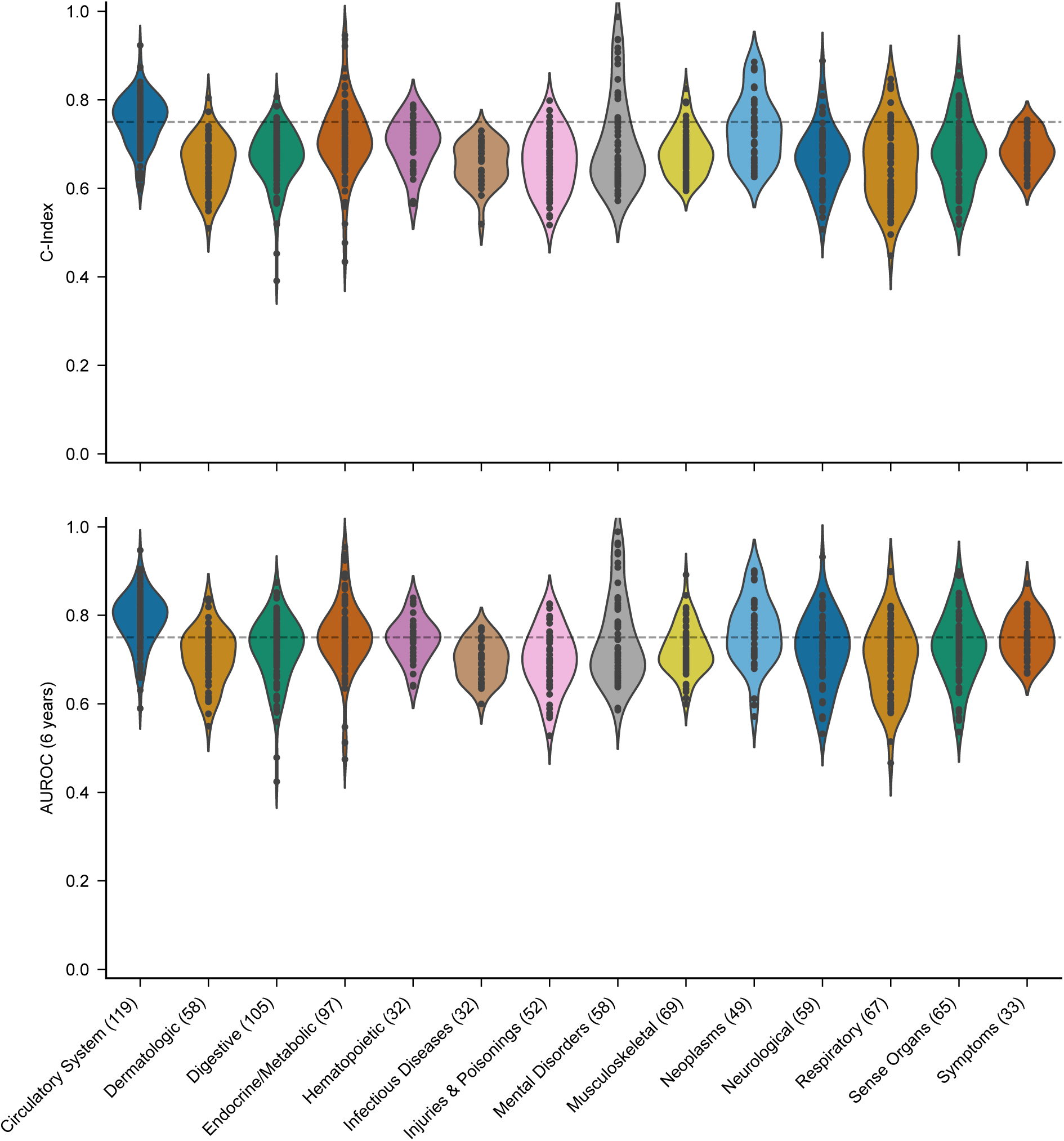
Performance of SleepFM on the held-out test set (n=5,019) as stratified by disease categories. Individual dots represents a disease within a category. The results are evaluated using two metrics: the C-Index, which measures the model’s ability to rank patient risk accurately, and the 6-year AUROC, which assesses the model’s discrimination performance by evaluating its ability to distinguish between patients who experience the event of interest and those who do not within a 6-year prediction window. For reference, the horizontal dashed line indicates a threshold of 0.75.

The model showed high accuracy for mild cognitive impairment (AUROC: 0.84 [0.80–0.88]), aligning with studies showing sleep disturbances as early markers of cognitive decline [30]. Strong performance was observed for Parkinson’s disease (0.93 [0.89–0.96]), where sleep disorders are increasingly recognized as potential early indicators [31], and developmental delays and disorders (0.84 [0.79–0.87]). Among circulatory conditions, the model effectively predicted hypertensive heart disease (0.88 [0.85–0.91]) and intracranial hemorrhage (0.82 [0.73–0.90]), consistent with established links between sleep disorders and cardiovascular risk [32]. In the Neoplasm category, the model showed strong predictive performance for several cancers: prostate cancer (0.90 [0.87–0.93]), breast cancer (0.90 [0.86–0.93]), and melanomas of skin (0.83 [0.76–0.90]). These findings align with existing literature linking sleep patterns to cancer risk [33, 34].

Drawing on sleep expertise and prior literature, we identified 14 conditions with strong potential links to sleep patterns. Prior studies associate sleep regularity with mortality [35], prolonged sleep with early neurodegeneration [36], and sleep disturbances with dementia [37], stroke [38], and cardiovascular outcomes [9]. Related phecodes were grouped into unified disease categories in consultation with a medical doctor (see Supplementary Table 6). Results for selected conditions—including death, stroke, heart failure, and dementia—are shown in Extended Figure 2. SleepFM demonstrates strong predictive performance, with particularly high accuracy for death (AUROC: 0.84 [0.80–0.88]), heart failure (0.83 [0.79–0.86]), chronic kidney disease (0.82 [0.79–0.85]), dementia (0.87 [0.84–0.91]), and stroke (0.81 [0.78–0.85]). All reported associations are statistically significant (p < 0.01, Bonferroni-corrected).

To better understand the physiological basis of disease prediction, we analyzed model performance stratified by both sleep stages and signal modalities. We found that while most sleep stages contribute similarly to disease prediction, certain stages such as Stage 1/2 and REM can offer slightly better predictive power for specific conditions, including cardiovascular and neurodegenerative diseases. Likewise, different signal modalities showed nuanced differences, with BAS signals better capturing mental and neurological conditions, RESP signals more predictive of respiratory and metabolic disorders, and EKG signals more informative for circulatory diseases. While these differences align with known physiology, the overall predictive performance was highest when combining all modalities. Full results and condition-specific breakdowns are provided in Supplementary Figures 3 and 4, and Supplementary Tables 7 and 8. Additionally, we trained separate SleepFM models on each modality to directly assess modality-level importance. Performance comparisons stratified by disease category, presented in Supplementary Tables 9 and 10, further confirm that combining all modalities yields the optimal performance.

**Figure 3:**
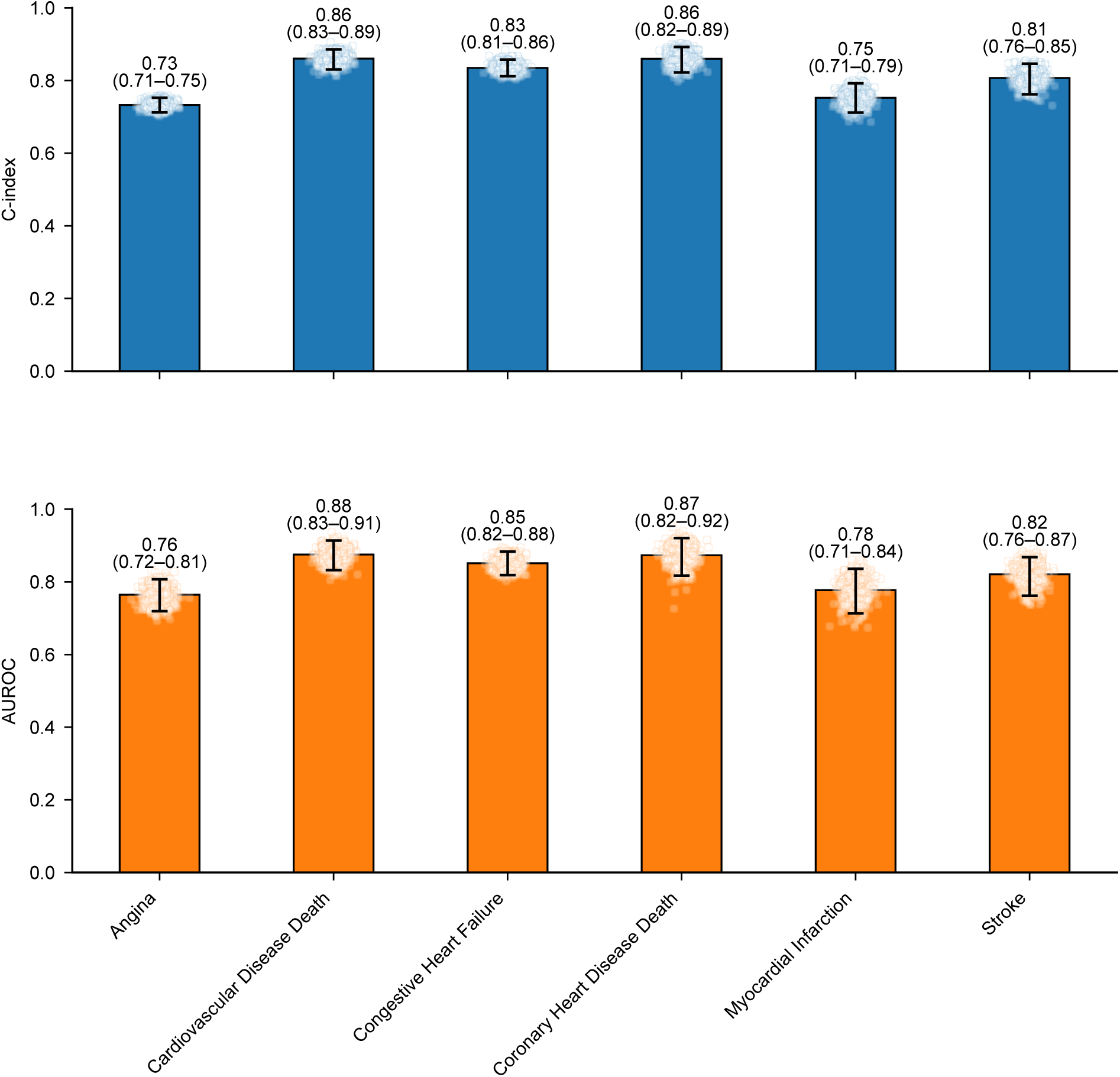
SleepFM prediction performance on the Sleep Heart Health Study (SHHS) test set (n=2,000 participants). Due to differences in available outcome data between SHHS and Stanford datasets, evaluation was limited to a subset of conditions. Results demonstrate transfer learning capabilities across these key clinical outcomes, including stroke, congestive heart failure, and cardiovascular disease-related mortality. Each panel uses bar plots derived from 1000 patient-level bootstrapping: faint points are individual bootstrap draws, and the vertical line with end caps marks the 95% bootstrap percentile CI. Numbers above bars report the mean. Metrics are C-index (top) and AUROC at 6 years (bottom). The number of positive samples for each outcome is as follows: Angina (704), Cardiovascular disease death (128), Congestive heart failure (190), Coronary heart disease death (80), Myocardial infarction (103), and Stroke (95). All conditions are statistically significant with a p-value < 0.01 after Bonferroni correction.

**Figure 4:**
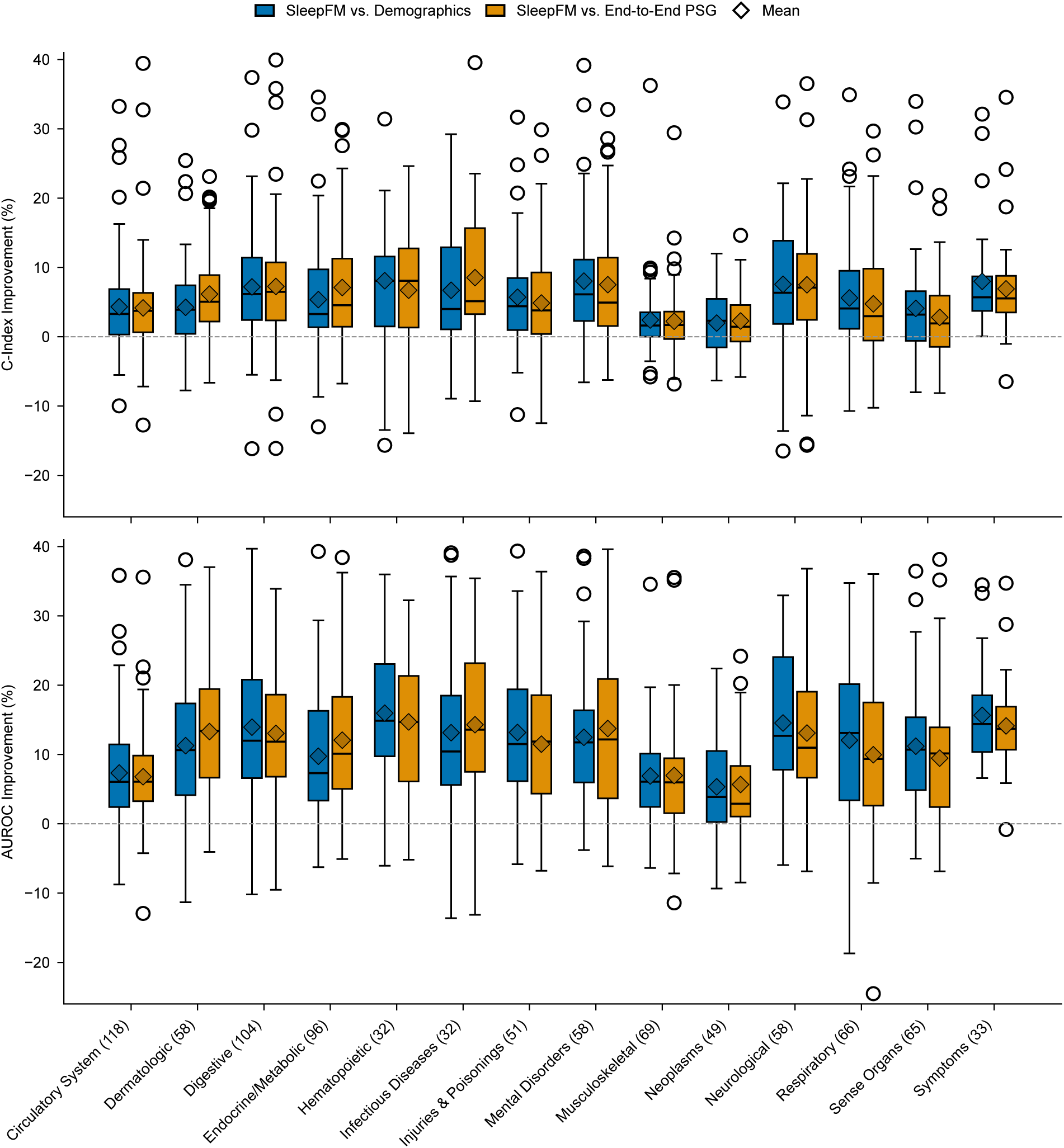
Performance improvements of SleepFM over baseline models across disease categories on Stanford test set (n=5,019 participants). SleepFM and the End-to-End PSG model include age and gender demo-graphic features, whereas the demographics-only model includes age, gender, body mass index (BMI), and race/ethnicity. Each box shows the distribution of disease-level percentage improvements of SleepFM relative to each baseline within the indicated disease category. Improvements are shown for both C-index (top) and 6-year AUROC (bottom) metrics. Boxes represent the interquartile range (IQR), with whiskers extending to 1.5 × IQR and outliers shown as points. Diamonds denote the mean improvement within each category. The horizontal dashed line at zero indicates no improvement.

## SleepFM Demonstrates Robust Generalization Across Time and cohorts

We evaluate SleepFM’s generalization capabilities across temporal distribution shifts and external site validation. For temporal generalization, we test the model on a separate cohort comprising Stanford patients from 2020 onwards. All model pre-training and training was done on data prior to 2020. Despite the limited follow-up period, SleepFM maintains strong predictive performance. Extended Figure 3 shows results for our 14 selected conditions, with particularly robust and statistically significant performance (Bonferroni-corrected p < 0.01) for death (0.83 [0.73–0.91]), heart failure (0.80 [0.75–0.85]), and dementia (0.83 [0.76–0.89]). Comprehensive temporal-split performance across all disease phenotypes and categories is provided in Supplementary Figures 5 and 6. Supplementary Figure 7 further reports temporal-split performance comparisons with baseline models, stratified by disease category.

To assess cross-site generalization, we evaluate SleepFM’s transfer learning capabilities on SHHS, a dataset entirely excluded from the pre-training phase. We use the pretrained model to extract embeddings and then fine-tune it on a subset of SHHS. Specifically, the SHHS fine-tuning set includes 3,291 participants, and the test set includes 2,000 participants. Due to differences in task availability between SSC and SHHS, our evaluation focuses on six overlapping cardiovascular conditions. This setup mimics real-world deployment scenarios where foundation models must be adapted to new clinical sites with minimal supervision.

As shown in Figure 3, SleepFM demonstrates strong transfer learning performance across key outcomes. For example, the model achieves statistically significant predictive accuracy (Bonferroni-corrected p < 0.01) for stroke (0.82 [0.76–0.87]), congestive heart failure (0.85 [0.82–0.88]), and cardiovascular disease-related mortality (0.88 [0.83–0.91]).

## SleepFM Surpasses Supervised Baselines in Disease Prediction

We compare SleepFM against two supervised baselines: Demographics and End-to-End PSG. The demographics baseline is a multi-layer perceptron (MLP) trained on structured clinical features (age, gender, race/ethnicity, and BMI). This baseline includes more demographic variables than the SleepFM-based models, which only use age and gender. The End-to-End PSG model is trained directly on raw PSG data using the same architecture and parameter count as SleepFM, and it includes age and gender but does not use any pretraining. From Figure 4, we observe that the percentage difference in AUROC between SleepFM and both baseline models ranges from 5% to 17%. The magnitude of improvement varies across disease categories; for example, gains are more pronounced in neurological and hematopoietic conditions, whereas in neoplasm-related conditions, the improvements are comparatively modest. Supplementary Figure 8 reports the overall test-set performance comparison between SleepFM and the baseline models across all disease phenotypes.

Next, we evaluate three different variants of SleepFM using identical training configurations, as shown in Table 2 and Extended Table 5. SleepFM-LSTM (w/o Demo) uses SleepFM embeddings with a 2-layer LSTM fine-tuning head but no demographic features. SleepFM-Linear uses SleepFM embeddings with a simple linear prediction head and includes age and gender. Finally, SleepFM-LSTM, combines the pretrained SleepFM embeddings with a 2-layer LSTM head and includes age and gender.

**Table 2:** Comparison of category-averaged AUROC across SleepFM variants and baselines. Category-averaged 6-year AUROC (mean with 95% CI) comparing SleepFM variants with two baselines across disease categories on Stanford cohort (n=5,019). The Demographics baseline (“Demo”) uses only structured clinical features (age, gender, BMI, and race/ethnicity). The End-to-End PSG baseline (“E2E-PSG”) is trained directly on raw PSG signals with age and gender, without any pretraining. SleepFM-1 denotes SleepFM-LSTM (w/o Demo), using two LSTM layers in the fine-tuning prediction module and no demographic features. SleepFM-2 denotes SleepFM-Linear, a linear prediction module on SleepFM embeddings with age and gender. SleepFM-3 denotes SleepFM-LSTM, which uses two LSTM layers in the fine-tuning prediction module with age and gender. Values are averaged within each category across conditions. Uncertainty is estimated via nonparametric bootstrapping (n=1000 resamples): for each resample, conditions within a category are sampled with replacement and the category mean is computed; 95% CIs are the 2.5th–97.5th percentiles across resamples.

| Category | Demo | E2E-PSG | SleepFM-1 | SleepFM-2 | SleepFM-3 |
| --- | --- | --- | --- | --- | --- |
| Circulatory System | 0.74<sub>(0.73, 0.74)</sub> | 0.74<sub>(0.73, 0.75)</sub> | 0.78<sub>(0.77, 0.78)</sub> | 0.79<sub>(0.78, 0.80)</sub> | 0.79<sub>(0.78, 0.80)</sub> |
| Dermatologic | 0.64<sub>(0.63, 0.65)</sub> | 0.63<sub>(0.62, 0.64)</sub> | 0.68<sub>(0.67, 0.69)</sub> | 0.71<sub>(0.70, 0.72)</sub> | 0.70<sub>(0.70, 0.71)</sub> |
| Digestive | 0.63<sub>(0.62, 0.64)</sub> | 0.64<sub>(0.63, 0.65)</sub> | 0.69<sub>(0.69, 0.70)</sub> | 0.72<sub>(0.71, 0.73)</sub> | 0.72<sub>(0.71, 0.73)</sub> |
| Endocrine/Metabolic | 0.68<sub>(0.68, 0.69)</sub> | 0.67<sub>(0.66, 0.68)</sub> | 0.74<sub>(0.73, 0.75)</sub> | 0.75<sub>(0.74, 0.76)</sub> | 0.75<sub>(0.74, 0.76)</sub> |
| Hematopoietic | 0.64<sub>(0.63, 0.66)</sub> | 0.66<sub>(0.64, 0.67)</sub> | 0.73<sub>(0.72, 0.75)</sub> | 0.75<sub>(0.73, 0.76)</sub> | 0.74<sub>(0.73, 0.76)</sub> |
| Infectious Diseases | 0.62<sub>(0.61, 0.64)</sub> | 0.62<sub>(0.60, 0.63)</sub> | 0.67<sub>(0.65, 0.69)</sub> | 0.70<sub>(0.68, 0.71)</sub> | 0.70<sub>(0.68, 0.71)</sub> |
| Injuries & Poisonings | 0.62<sub>(0.61, 0.63)</sub> | 0.63<sub>(0.61, 0.64)</sub> | 0.68<sub>(0.67, 0.69)</sub> | 0.70<sub>(0.69, 0.71)</sub> | 0.70<sub>(0.69, 0.71)</sub> |
| Mental Disorders | 0.66<sub>(0.65, 0.67)</sub> | 0.66<sub>(0.66, 0.67)</sub> | 0.72<sub>(0.71, 0.73)</sub> | 0.74<sub>(0.73, 0.75)</sub> | 0.74<sub>(0.74, 0.75)</sub> |
| Musculoskeletal | 0.68<sub>(0.67, 0.68)</sub> | 0.68<sub>(0.67, 0.69)</sub> | 0.70<sub>(0.69, 0.71)</sub> | 0.72<sub>(0.72, 0.73)</sub> | 0.72<sub>(0.71, 0.73)</sub> |
| Neoplasms | 0.73<sub>(0.71, 0.74)</sub> | 0.73<sub>(0.71, 0.74)</sub> | 0.73<sub>(0.72, 0.74)</sub> | 0.76<sub>(0.75, 0.77)</sub> | 0.76<sub>(0.75, 0.77)</sub> |
| Neurological | 0.62<sub>(0.61, 0.63)</sub> | 0.63<sub>(0.62, 0.64)</sub> | 0.70<sub>(0.69, 0.71)</sub> | 0.72<sub>(0.71, 0.73)</sub> | 0.72<sub>(0.71, 0.73)</sub> |
| Respiratory | 0.63<sub>(0.62, 0.64)</sub> | 0.64<sub>(0.63, 0.65)</sub> | 0.69<sub>(0.68, 0.70)</sub> | 0.69<sub>(0.69, 0.70)</sub> | 0.70<sub>(0.69, 0.71)</sub> |
| Sense Organs | 0.66<sub>(0.65, 0.67)</sub> | 0.67<sub>(0.66, 0.68)</sub> | 0.71<sub>(0.70, 0.72)</sub> | 0.73<sub>(0.72, 0.74)</sub> | 0.73<sub>(0.72, 0.74)</sub> |
| Symptoms | 0.65<sub>(0.64, 0.66)</sub> | 0.66<sub>(0.64, 0.67)</sub> | 0.72<sub>(0.71, 0.73)</sub> | 0.75<sub>(0.74, 0.76)</sub> | 0.75<sub>(0.74, 0.76)</sub> |

As seen in Table 2, the demographics-only baseline performs well, reflecting the fact that many diseases are strongly associated with age, gender, BMI, and race/ethnicity. For example, in the Neoplasm category, older age is a strong predictor of cancer risk. Nevertheless, all SleepFM-based models, including the SleepFM-LSTM (w/o Demo) variant, consistently outperform the demographics and End-to-End PSG baselines across most disease categories. This demonstrates the benefit of using pretrained SleepFM embeddings for disease prediction. Additionally, SleepFM-LSTM (w/o Demo) achieves over +5 AUROC points in 9 out of 14 conditions, while SleepFM-Linear and SleepFM-LSTM achieve over +5 AUROC points in 12 out of 14 conditions, compared to supervised demographics baseline. Importantly, as seen from the 95% CI bars, these improvements are robust, with most differences being larger than the uncertainty intervals. Finally, SleepFM-Linear performs comparably to SleepFM-LSTM, suggesting that the strength of the model lies in the pretrained embeddings rather than the complexity of the downstream head. Percentage improvement comparisons across models are provided in Supplementary Figure 9, and a scatter-plot comparison of all disease phenotypes across different fine-tuning architectures on top of SleepFM is shown in Supplementary Figure 10.

To further examine disease-specific performance, full results are provided in Supplementary Tables 11, 12, and 13, and clinician-selected conditions are presented in Supplementary Figure 11. These comparisons show that SleepFM achieves substantial gains across multiple neurological, mental, circulatory, endocrine/metabolic, and respiratory conditions. For neurological and mental disorders, SleepFM attains higher C-Index scores for senile dementia (0.99 [0.98–1.00] vs. 0.87 [0.75–0.96]), myoneural disorders (0.81 [0.73–0.88] vs. 0.42 [0.28–0.55]), and developmental delays (0.80 [0.77–0.84] vs. 0.58 [0.51–0.64]). For circulatory diseases, SleepFM outperforms in atherosclerosis (0.92 [0.88–0.95] vs. 0.74 [0.64–0.89]) and acute pulmonary heart disease (0.80 [0.75–0.85] vs. 0.74 [0.68–0.80]). Improvements in endocrine/metabolic conditions include diabetes type 2 with circulatory complications (0.87 [0.83–0.91] vs. 0.79 [0.74–0.85]) and diabetic retinopathy (0.81 [0.77–0.85] vs. 0.75 [0.69–0.80]). For respiratory conditions, SleepFM achieves higher C-Index in respiratory insufficiency (0.79 [0.72–0.85] vs. 0.59 [0.51–0.67]) and failure (0.77 [0.73–0.80] vs. 0.70 [0.65–0.74]). These findings highlight the versatility of SleepFM in predicting a broad range of diseases beyond what is captured by demographics alone.

Similarly, full comparisons with the End-to-End PSG model are provided in Supplementary Table 14. This comparison highlights the value of foundation model pre-training: although both models share similar architecture and input signals, SleepFM benefits from self-supervised pre-training, enabling more robust and informative representations. This advantage is reflected in consistent performance gains across neurological, circulatory, endocrine/metabolic, and respiratory conditions. For neurological and mental disorders, SleepFM outperforms the end-to-end model in myoneural disorders (0.84 [0.75–0.91] vs. 0.54 [0.40–0.69]), developmental delays (0.84 [0.79, 0.87] vs. 0.61 [0.52, 0.69]), and speech/language disorders (0.83 [0.74, 0.90] vs. 0.71 [0.60, 0.83]). For circulatory conditions, improvements are observed in atherosclerosis of native arteries of the extremities (0.95 [0.92, 0.98] vs. 0.65 [0.61, 0.69]), atherosclerosis of the extremities (0.84 [0.75, 0.90] vs. 0.78 [0.71, 0.85]), and acute pulmonary heart disease (0.84 [0.77, 0.90] vs. 0.76 [0.69, 0.83]). In endocrine/metabolic disorders, SleepFM demonstrates stronger performance for predicting diabetes with circulatory complications (0.89 [0.85, 0.93] vs. 0.79 [0.70, 0.87]), neurological manifestations (0.86 [0.81, 0.90] vs. 0.73 [0.67, 0.78]), and diabetic retinopathy (0.84 [0.79, 0.89] vs. 0.76 [0.69, 0.82]). Respiratory conditions also benefit, with better performance in predicting respiratory insufficiency (0.82 [0.72, 0.91] vs. 0.64 [0.54, 0.73]) and respiratory failure (0.76 [0.71, 0.82] vs. 0.68 [0.62, 0.74]). In predicting all-cause mortality, SleepFM achieves a AUROC of 0.85 [0.80–0.89], outperforming both the demographic baseline and end-to-end PSG model, which achieve AUROC of 0.78 [0.72–0.82].

Finally, we compare fine-tuning scalability by evaluating SleepFM alongside two baseline models as we increase the amount of fine-tuning data and measure performance on the same test set. These results are shown in Extended Figure 4 for SHHS and Extended Figure 5 and Supplementary Figure 12 for SSC. In both plots, the key observation is that SleepFM consistently outperforms the supervised baselines, with its performance steadily improving as more data is used, remaining above the baseline curves for nearly all conditions. For SHHS, SleepFM surpasses the Demographics baseline in 5 out of 6 conditions across all data percentages, with particularly large improvements in smaller dataset splits. For example, SleepFM trained on just 10% of the data outperforms the Demographics baseline trained on five times more data across all conditions in SSC and 4 out of 6 conditions in SHHS (e.g., cardiovascular disease death, congestive heart failure, myocardial infarction, and stroke). SleepFM also outperforms the End-to-End PSG baseline in 5 out of 6 conditions, although the gap is slightly smaller than with the Demographics baseline. Importantly, SleepFM exhibits stable scaling behavior across data percentages, with smoother performance improvements, while the baseline models show greater variability.

## Discussion

This study presents a large-scale foundation model for sleep analysis, developed on over 585,000 hours of PSG data from 65,000 subjects. Our work makes several contributions. First, we address challenges in sleep analysis by leveraging self-supervised learning to train a foundation model that learns from unlabeled data and is agnostic to channel type and number, enabling broad exploration of sleep data across diverse clinical settings. Second, through extensive evaluation across 1,041 disease phenotypes, we demonstrate sleep’s broad predictive power for diverse health outcomes. The model shows strong performance in predicting death (C-Index: 0.84), dementia (0.85), heart failure (0.80), and chronic kidney disease (0.79). Third, we demonstrated transfer learning capabilities through strong performance on the SHHS dataset. Despite SHHS being entirely excluded from pre-training, our model maintains robust predictive power for key outcomes such as stroke (C-Index: 0.81), congestive heart failure (0.83), and death related to cardiovascular disease (0.86). Finally, SleepFM achieves competitive performance on standard sleep analysis tasks, including sleep staging and apnea detection, with mean F1 scores ranging from 0.70 to 0.78 across cohorts—comparable to state-of-the-art models such as U-Sleep [7], GSSC [25], STAGES [10], and YASA [24]. Additionally, in a fully external validation setting, SleepFM outperforms all models on DCSM (F1 = 0.68) and is competitive with the PhysioEx [26] models. For apnea classification, SleepFM achieves 87% accuracy in MESA, MrOS, and SHHS, comparable to state-of-the-art [8].

SleepFM predicts all-cause mortality more accurately than both the demographics-based model and the end-to-end PSG model, achieving a higher C-Index of 0.84 [0.81–0.87], compared to 0.79 [0.75–0.82]. This indicates that pre-training efficiently captures subtle mortality-related signals in the PSG data. Research shows strong association between all-cause mortality and sleep-related factors, including high arousal burden [39], low REM sleep [40], sleep-disordered breathing [41], hypoxemia, and low sleep efficiency [42]. Increased “brain age” derived from EEG has also been identified as a significant predictor of mortality [3]. SleepFM likely integrates these multifactorial contributors, capturing respiratory events, sleep fragmentation, arousal burden, and sleep efficiency, along with markers of cardiovascular, metabolic, and other diseases.

Predictive and prognostic models for neurological and mental disorders are rapidly advancing, offering the potential for earlier and more individualized treatment. Among the top conditions predicted by SleepFM were Alzheimer’s disease and Parkinson’s disease, with C-indices of 0.91 [0.87–0.98] and 0.89 [0.85–0.92], respectively. Sleep disorders are strongly associated with preclinical Alzheimer’s disease [43], including abnormalities in Non-REM sleep, such as reduced slow-wave activity [44], REM sleep disturbances [45], and decreased spindle activity [46]. In early Alzheimer’s disease, REM sleep abnormalities have been linked to basal forebrain cholinergic lesions, which likely contribute to cognitive decline [47]. Similarly, Parkinson’s disease is frequently preceded by REM sleep behavior disorder (RBD), characterized by REM sleep without atonia and abnormalities in BAS and ECG patterns [48]. Recent studies have also shown that respiratory signals can capture phenotypes specific to Parkinson’s disease [49].

Consistent with these findings, SleepFM identified BAS as the strongest predictor of neurological and mental disorders, while respiratory signals were particularly effective in predicting senile dementia. Most studies in this domain rely on imaging modalities such as MRI and fMRI to predict dementia. For example, one study using hippocampal MRI achieved a C-index of 0.86 [50], while another using fMRI reported an AUROC of 0.82 for predicting dementia up to nine years in advance [51]. Although direct performance comparisons are challenging due to differences in sample distributions, the ability of SleepFM to leverage PSG data to predict neurological and mental disorders underscores its potential as an alternative to imaging-based approaches.

Other established biomarkers for Alzheimer’s disease—such as amyloid PET, decreased CSF β-amyloid_42_, and increased CSF phosphorylated tau (e.g., p-tau_129_) [52, 53]—have been widely used for diagnosis and prognosis. More recently, plasma p-tau_217_ has emerged as a promising less invasive marker [54]. Sleep biomarkers from PSG data offer a complementary, non-invasive tool for the prognosis of dementia and mild cognitive impairment.

SleepFM accurately modeled cardiovascular disease in both the SSC and SHHS datasets, leveraging data-driven methods commonly used in prognostic modeling of cardiovascular disease, particularly with ECG data [55] and lead II ECG from PSG studies [9]. Foundation models have demonstrated state-of-the-art performance with ECG data in various cross-sectional tasks [15]. For predicting cardiovascular mortality over 10 years, a previous study reported an AUROC of 0.84 [0.78–0.89] in a subset of SHHS participants with sleep apnea, whereas SleepFM achieved a slightly higher AUROC of 0.88 [0.83–0.91]. Similarly, for atrial fibrillation, earlier work reported an AUROC of 0.82 [9], which aligns with SleepFM’s performance of 0.81 [0.78–0.84]. Our ablation study further demonstrated that both ECG and respiratory signals contribute to the prediction of circulatory system phenotypes, suggesting that SleepFM integrates information on sleep apnea and heart activity in ways that are consistent with known disease mechanisms [56].

Most disease categories, including neurological, circulatory, hematopoietic, mental disorders, and endocrine/metabolic, were predicted with significantly improved performance by SleepFM compared to the demographics-based and end-to-end PSG baseline models. Many of these diseases are either associated with sleep (e.g., type 2 diabetes [57]) or directly influenced by the signal modalities (e.g., heart arrhythmia). Disrupted and unhealthy sleep contributes to dysfunction across multiple physiological systems, increasing the risk of diseases such as obesity, type 2 diabetes, hypertension, stroke, and cardiovascular disease [58]. Sleep-specific conditions, including sleep apnea [56] and less conclusively periodic leg movements [59], are also linked to cardiovascular outcomes. Additionally, specific EEG waveforms, such as coupled slow-wave and spindle activity, have been identified as markers of next-day blood glucose regulation [60].

Despite these promising results, several limitations should be acknowledged. While our dataset is large, it primarily consists of patients referred for sleep studies due to suspected sleep disorders or other medical conditions requiring overnight monitoring. This selection bias means our cohort is not representative of the general population, as individuals without sleep complaints or those with limited access to specialized sleep clinics are underrepresented. The model’s performance shows some degradation in temporal test sets, highlighting the challenge of maintaining predictive accuracy over time as clinical practices and patient populations evolve. Additionally, interpreting the predictions made by SleepFM is inherently challenging due to the complexity of the learned features during training by a deep model. To mitigate this, we stratified the model’s performance across sleep stages and data modalities, and conducted evaluations on temporal test sets and unseen datasets to gain insights into its behavior. However, further work is needed to enhance case-level interpretability and understand the specific sleep patterns and features driving these predictions.

In building our model, we selected hyperparameters for SleepFM based on prior work and ensured all training converged in loss; more extensive hyperparameter searches may further boost performance. Furthermore, while we evaluated SleepFM’s transfer learning performance on an independent dataset, SHHS, only a subset of the full 1,041 conditions could be assessed in this sample due to limited diagnostic overlap with SSC; this prevented a comprehensive evaluation of generalization across the full spectrum of diseases. Our sleep apnea analysis was limited to binary and four-class classification based on AHI thresholds; we did not explore more granular formulations such as AHI regression or event detection, we leave this for future research. Similarly, while SleepFM achieves competitive results on sleep staging tasks across most datasets, it lags behind specialized sleep staging models on certain external validation datasets (e.g., HMC). Further specialized modeling may be necessary to optimize SleepFM for sleep staging.

This study underscores the potential of sleep-based foundation models for risk stratification and longitudinal health monitoring. By integrating multiple physiological signals and leveraging large-scale pretraining, SleepFM performs consistently well across diverse disease categories and outperforms supervised baselines. Its stable performance across fine-tuning splits suggests that pretraining may improve model generalizability, particularly in clinical contexts with limited labeled data. These results suggest that SleepFM can complement existing risk assessment tools and help identify early signs of diseases. As wearable sleep technologies continue to advance, models like SleepFM may offer opportunities for non-invasive, real-time health monitoring. Future efforts should explore how combining sleep-based models with data from electronic health records, omics, and imaging can further enhance their utility.

## Data Availability

Part of the data used in this study is publicly available (MESA, MrOS, SHHS). Data from Stanford Sleep Clinic will be available in near future. Data from Bioserenity is proprietary.

https://github.com/zou-group/sleepfm-clinical

## Acknowledgements

The Multi-Ethnic Study of Atherosclerosis (MESA) Sleep Ancillary study was funded by NIH-NHLBI Association of Sleep Disorders with Cardiovascular Health Across Ethnic Groups (RO1 HL098433). MESA is supported by NHLBI funded contracts HHSN268201500003I, N01-HC-95159, N01-HC-95160, N01-HC-95161, N01-HC-95162, N01-HC-95163, N01-HC-95164, N01-HC-95165, N01-HC-95166, N01-HC-95167, N01-HC-95168 and N01-HC-95169 from the National Heart, Lung, and Blood Institute, and by cooperative agreements UL1-TR-000040, UL1-TR-001079, and UL1-TR-001420 funded by NCATS. The National Sleep Research Resource was supported by the National Heart, Lung, and Blood Institute (R24 HL114473, 75N92019R002).

The National Heart, Lung, and Blood Institute provided funding for the ancillary MrOS Sleep Study, "Outcomes of Sleep Disorders in Older Men," under the following grant numbers: R01 HL071194, R01 HL070848, R01 HL070847, R01 HL070842, R01 HL070841, R01 HL070837, R01 HL070838, and R01 HL070839.

The National Sleep Research Resource was supported by the National Heart, Lung, and Blood Institute (R24 HL114473, 75N92019R002).

The Sleep Heart Health Study (SHHS) was supported by National Heart, Lung, and Blood Institute cooperative agreements U01HL53916 (University of California, Davis), U01HL53931 (New York University), U01HL53934 (University of Minnesota), U01HL53937 and U01HL64360 (Johns Hopkins University), U01HL53938 (University of Arizona), U01HL53940 (University of Washington), U01HL53941 (Boston University), and U01HL63463 (Case Western Reserve University). The National Sleep Research Resource was supported by the National Heart, Lung, and Blood Institute (R24 HL114473, 75N92019R002).

R.T. is supported by the Knight-Hennessy Scholars funding. E.M. and B.W. are supported by a grant from the National Heart, Lung, and Blood Institute of the NIH (R01HL161253). J.Z. is supported by funding from the Chan-Zuckerberg Biohub.

We would also like to acknowledge Ethan Steinberg for his valuable insights into survival analysis.

## Author Contributions Statement

R.T. and M.R.K. contributed equally to brainstorming the project, running experiments, and writing the manuscript. B.H., I.C., P.J., A.B.K., and B.W. provided high-level brainstorming and contributed to writing and editing the paper. U.H., G.G. and H.M. assisted with data access. E.M. and J.Z., as senior co-authors, conceived the project and provided overall guidance. All authors reviewed and approved the final manuscript.

## Competing Interests Statement

B.W. is a co-founder, scientific advisor, consultant to, and has personal equity interest in Beacon Biosignals. The remaining authors declare no competing interests.

## Methods

### Dataset and Preprocessing

Our dataset includes PSG recordings from four different sites: SSC, BioSerenity, MESA [20, 21], and MROS [20, 22], with SHHS [20, 23] serving as an external validation dataset. Among these, MESA, MROS, and SHHS are publicly available datasets, while SSC is our proprietary dataset. The BioSerenity dataset, provided by the BioSerenity company, contains 18,869 overnight recordings lasting 7-11 hours each. This dataset is a subset of a larger collection from SleepMed and BioSerenity sleep laboratories, gathered between 2004 and 2019 across 240 US facilities [19]. At the time of this study, approximately 20K de-identified PSGs were available for analysis. The dataset distribution across different splits is shown in Figure 1, with SSC constituting the largest cohort. To prevent data leakage, participants with multiple PSG recordings were assigned to a single split. For MESA, MROS, and SHHS details, we refer readers to their original publications. Below, we describe our internal SSC dataset in more detail.

The SSC dataset comprises 35,052 recordings, each lasting approximately 8 hours overnight. It includes diverse waveforms such as BAS, ECG, EMG, and respiratory channels, making it a high-quality resource for sleep-related research. The dataset spans recordings from 1999 to 2024 and includes participants aged 2 to 96. The patient demographic statistics for SSC and BioSerenity are summarized in Extended Table 1 and Extended Table 2 respectively.

Our preprocessing strategy minimizes alterations to preserve raw signal characteristics crucial for identifying nuanced patterns. Each recording contains up to four modalities: BAS, ECG, EMG, and respiratory, with variable numbers of channels. For BAS, we allowed up to 10 channels, for ECG 2 channels, for EMG 4 channels, and for respiratory 7 channels. The number and type of channels vary across sites and even between patients within the same site, depending on the study type. The types of channels available across sites are described in Supplementary Tables 15, 16, 17, 18, and 19. BAS includes channels that measure brain activity from different regions (frontal, central, occipital) as well as EOG for eye movements. EMG records electrical activity in muscles, while ECG captures cardiac electrical function. Respiratory channels measure chest and abdominal movements, pulse readings, and nasal/oral airflow. These channels were selected based on their relevance to sleep studies, guided by sleep experts [1].

Each PSG recording is resampled to 128 Hz to standardize sampling rates across participants and sites. Before downsampling, we utilized a 4^th^-order low-pass Butterworth filter to prevent aliasing, applied in a zero-phase setting to avoid phase distortion. Finally, we standardized the signal to have zero mean and unit variance. For any signals that needed to be upsampled, this was done using linear interpolation. Due to the channel agnostic model design, we did not need any other data-harmonization. Signals are segmented into 5-second patches, with each segment embedded into a vector representation for transformer model processing. To prevent data leakage, PSGs were split into pre-train, train, validation, test, and temporal test sets early in the preprocessing pipeline. While there is overlap between the pre-training and training sets, no overlap exists with the validation, test, or temporal test sets. The SHHS serves as an independent dataset not used during pre-training, instead being used to evaluate the model’s ability to adapt to a new site through lightweight fine-tuning.

During pre-training, the only required labels are the modality types of the signals. A self-supervised contrastive learning (CL) objective is employed for pre-training. For downstream evaluations, we consider canonical tasks such as age/gender prediction, sleep stage classification, sleep apnea classification, and various patient conditions extracted from EHR data. Sleep staging and apnea labels for SSC, MESA, MROS, and SHHS were annotated by sleep experts. To ensure consistency across and within datasets, R&K labels were converted to AASM by mapping R&K stages 3 and 4 to AASM N3. SHHS also includes diagnostic information for conditions such as myocardial infarction, stroke, angina, congestive heart failure, and death. For SSC, we paired PSG data with Stanford EHR data using de-identified patient IDs to extract demographic and diagnostic information. As BioSerenity lacks associated labels, it was used exclusively for pre-training.

### SleepFM Model Architecture

Our model architecture is illustrated in Figure 1. The architecture includes several key components that differ slightly between the pre-training and fine-tuning stages. During pre-training, we employ contrastive learning (CL) as the objective function for representation learning. A single model processes all four modalities.

The first component of the architecture is the *Encoder*, a 1D convolutional neural network (CNN) that processes raw signal data for each modality separately. The encoder takes raw input vectors, where the length of each vector corresponds to a 5-second segment of the signal, referred to as a token. The input dimensions are (*B, T, C*), where *B* is the batch size, *T* is the raw temporal length of the input, and *C* is the number of channels for each modality. These inputs are reshaped into (*B, C, S, L*), where *S* is the sequence length representing the number of tokens (*S = T/L*), and *L* corresponds to the raw vector length for a single token (e.g., 640 samples). Each token is then processed individually through a stack of six convolutional layers, each followed by normalization and ELU activation layers. These layers progressively reduce the temporal resolution while increasing the number of feature channels, converting the input from one channel to 128 channels. After this, adaptive average pooling further reduces the temporal dimensions, and a fully connected layer compresses the representation into a 128-dimensional embedding for each token. The final output of the encoder has dimensions (*B, C, S, D*), where *D* = 128.

Following the encoder, a sequence of transformer-based operations is applied to extract and aggregate modality-specific and temporal features. The first step is *channel pooling*, which aggregates token embeddings from all channels within a given modality. This operation uses an attention pooling mechanism based on a transformer layer to compute attention scores for each channel and produces a single aggregated embedding per time segment by averaging over the channel dimension. The resulting embeddings, with dimensions (*B, S, D*), are then passed through a *temporal transformer*, which operates along the temporal dimension to capture dependencies between tokens. The temporal transformer applies sinusoidal positional encoding to the token embeddings, followed by 2 transformer blocks consisting of self-attention and feedforward layers, enabling the model to learn contextual relationships across the sequence. After temporal modeling, the embeddings are processed through *temporal pooling*, which aggregates token embeddings over the sequence length (S) for each modality. Similar to channel pooling, *temporal pooling* uses an attention mechanism to compute weighted averages, generating a compact representation of size (*B*, 128) per modality. These steps collectively ensure that the model captures both spatial and temporal dependencies while reducing dimensionality for computational efficiency.

The final output is a single 128-dimensional embedding for each modality, used for contrastive learning during pre-training. While the 5-minute recordings are used exclusively for pre-training, we retain the 5-second-level embeddings for each modality for downstream tasks such as sleep staging and disease classification.

### Baseline Models

We evaluate SleepFM against two carefully chosen baseline approaches to demonstrate the value of our foundation model methodology.

The first baseline is a simple demographic model that processes only patient characteristics, including age, gender, body mass index (BMI), and race/ethnicity information. This demographic baseline is implemented as a one-layer multi-layer perceptron (MLP) to establish a minimum performance threshold using only basic patient data available in most clinical settings.

The second baseline is a more sophisticated end-to-end PSG model that directly processes raw sleep recordings. This model uses the same architecture as SleepFM, including the 1D CNN encoder, channel pooling transformer block, temporal transformer block, temporal pooling transformer block, and the LSTM layers, and is trained from scratch on the same dataset used for downstream evaluation. It also includes age and gender demographic features to ensure a fair comparison, but does not leverage any pretraining—serving to isolate the benefit of task-specific supervised learning on PSG signals without a foundation model.

All baseline models were trained using dataset splits shown in Table 1. The foundation model was first pretrained on the training dataset using a self-supervised objective, and subsequently fine-tuned on the same data. In contrast, the supervised baseline models were trained end-to-end without any pretraining. Although all models share identical architectures, training objectives, and data splits, SleepFM consistently outperforms both baselines across a range of clinical prediction tasks. While this may seem counterintuitive—given that the supervised PSG baseline is trained on the same data—these results align with well-established benefits of pretraining in representation learning. Self-supervised pretraining enables the model to learn more generalizable physiological representations, facilitates better convergence through improved initialization, and makes more efficient use of sparse or noisy supervision during fine-tuning, as demonstrated in prior work [11].

### Model Training

Model training can be categorized into two segments: pre-training and fine-tuning. Pre-training stage involves self-supervised representation learning with a CL objective and fine-tuning involves training the model with supervised learning objective for specific tasks such as sleep stage classification, sleep apnea classification, and disease prediction. We describe these in more details below.

### Pre-training

Model pre-training is performed using a self-supervised learning objective called CL. Specifically, we employ a CL objective for multiple modalities, referred to as leave-one-out contrastive learning (LOO-CL). The key idea behind CL is to bring positive pairs of embeddings from different modalities closer in the latent space while pushing apart negative pairs. Positive pairs are derived from temporally aligned 5-minute aggregated embeddings, obtained after temporal pooling, across four different modalities. All other non-matching instances within a training batch are treated as negative pairs.

In LOO-CL, we define a predictive task where an embedding from one modality attempts to identify the corresponding embeddings from the remaining modalities. For each modality *i*, we construct an embedding *x̄^−i^_k_* by averaging over embeddings from all other modalities, excluding modality *i*. We then apply a contrastive loss between the embedding of modality i and this leave-one-out representation:

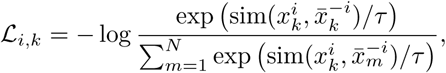

where *L_i,k_* is the loss for a sample *k* from modality *i* in a given batch, sim(·) represents a similarity function (e.g., cosine similarity), and *τ* is a temperature scaling parameter. The numerator computes the similarity between the embedding of modality *i* and the leave-one-out representation of the corresponding sample, while the denominator sums the similarities across all samples within the batch. The motivation behind the leave-one-out method is to encourage each embedding to align semantically with all other modalities.

### Fine-tuning

After pre-training with the CL objective, we extract 5-second embeddings for all patient PSG data across modalities. We standardize the temporal context to 9 hours for all patients - longer recordings are cropped and shorter ones are zero-padded to ensure consistent input dimensions. For example, for a patient’s standardized 9-hour sleep data, the resulting patient matrix has dimensions (4 × 6480 × 128), where 4 represents the number of modalities, 6480 is the number of 5-second embeddings for 9 hours of sleep, and 128 is the embedding vector dimension.

During fine-tuning, we first apply a channel pooling operation across different modalities, reducing the dimensions to (6480 × 128) for our example patient matrix. The pooled embeddings are then processed through a 2-layer LSTM block, which is designed to handle temporal sequences. For sleep staging tasks, these 5-second embeddings are directly passed through a classification layer. For all other tasks, the embeddings are first pooled along the temporal dimension before being passed through an output layer.

For disease classification, we append age and gender features to the mean-pooled embedding vector after the LSTM block, before passing it to the final output layer. This addition empirically improves performance and surpasses the demographic baseline.

The fine-tuning objective for disease prediction uses the CoxPH loss function, a standard approach in survival analysis for modeling time-to-event data. The CoxPH loss maximizes the partial likelihood and is defined for a single label as:

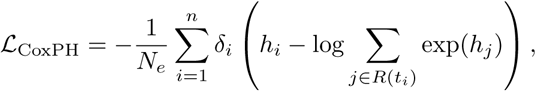

where *h_i_* is the predicted hazard for the i-th patient, *δ_i_* is the event indicator (1 for event occurrence, 0 otherwise), *t_i_* is the event or censoring time, *R(t_i_)* represents the risk set of all patients with event times greater than or equal to *t_i_*, *n* is the total number of patients, and 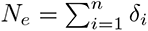 is the number of events. For our multi-label setup with 1,041 labels, we extend the CoxPH loss by computing it independently for each label and summing the results:

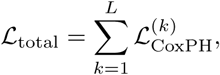

where *L* is the total number of labels.

Given the large dataset size, computing the loss for all patients in a single batch is computationally infeasible. Therefore, we calculate the loss in smaller batches of 32 samples, with patients sorted by event time in descending order to ensure correct computation of the partial likelihood. This batching strategy, combined with the summation of per-label losses, provides an efficient and scalable approach for multi-label time-to-event modeling.

### Architectural Details

We provide additional implementation-level details to clarify how SleepFM is constructed and trained. The design of SleepFM was developed through an empirical and iterative process, informed by domain knowledge and guided by practical training considerations. While we did not perform an exhaustive hyperparameter search, we systematically evaluated architectural variants through trial-and-error by monitoring loss convergence, training stability, and downstream performance.

Each 5-second segment of raw PSG signals (640 timepoints at 128Hz) is passed through a tokenizer composed of six convolutional layers with increasing feature maps: 1 → 4 → 8 → 16 → 32 → 64 → 128. Each convolutional block includes BatchNorm, ELU activation, and LayerNorm. After convolution, adaptive average pooling reduces the temporal axis to 1, and a linear layer projects the features to a fixed 128-dimensional token embedding. The resulting output shape is (B, C, S, 128), where C is the number of channels and S is the number of 5-second tokens.

To accommodate variability in the number and ordering of channels across different PSG datasets, we introduced an attention-based spatial pooling layer that operates across channels using a transformer encoder. This design makes the model robust to inconsistencies in recording configurations across sites. Specifically, embeddings from multiple channels within a modality are pooled using multi-head self-attention, producing a modality-specific sequence of shape (B, S, 128).

To capture long-range temporal dependencies in sleep signals, the pooled token sequence is passed through three transformer encoder layers (each with 8 heads, batch-first configuration, and a dropout rate of 0.3), along with sinusoidal positional encoding and LayerNorm. This component enables modeling of contextual relationships across the sleep sequence. The output shape remains (B, S, 128).

An additional attention-based pooling layer aggregates the temporal sequence across time steps, resulting in a single 128-dimensional embedding for each modality (e.g., BAS, ECG, EMG, or respiratory). These fixed-size modality-specific embeddings are used for pretraining with a self-supervised contrastive learning objective.

For downstream disease prediction, 5-second token embeddings spanning a standardized 9-hour window are processed by a fine-tuning head. This head includes spatial pooling followed by a two-layer bidirectional LSTM (hidden size = 64). Temporal mean pooling is applied across valid time steps, and normalized age and sex features are concatenated with the pooled output. The combined vector is then passed through a final linear layer to generate hazard scores for each disease. The total number of learnable parameters in this setup is approximately 0.91 million.

The supervised baseline model uses the same architecture as SleepFM but is trained from scratch without pretraining. The demographics-only baseline passes four input features—age, sex, BMI, and race/ethnicity—through a shallow MLP with dimensions 4 → 128 → output.

### Implementation Details

All implementations were carried out using PyTorch, a widely used library for deep learning. The PSG data was gathered and processed within a HIPAA-compliant and secure compute cluster on Google Cloud Platform. Patient electronic health record data was likewise stored and analyzed exclusively within this secure environment.

For pre-training, the model was trained with a batch size of 32, a learning rate of 0.001, 8 pooling heads, 3 transformer layers, and a dropout rate of 0.3. As previously described, each patch size corresponds to a 5-second segment, and the total sequence length is 5 minutes for the transformer model. The total parameter count for the model was approximately 4.44 million. Pre-training was performed on 432,000 hours of sleep data collected from 48,000 participants for one epoch, using an NVIDIA A100 GPU. The entire pre-training process took approximately 15 hours.

For fine-tuning, the batch size was also set to 32, with a learning rate of 0.001, 4 pooling heads, 2 LSTM layers, and a dropout rate of 0.3. The fine-tuned model had approximately 0.91 million learnable parameters. Training was conducted on patient data, with each token embedding represented as a 128-dimensional vector, over 10 epochs. The fine-tuning process was performed on an NVIDIA A100 GPU, with the total training time per epoch ranging from 2 to 5 minutes, depending on the task.

All data analysis and preprocessing were performed using Python (version 3.10.14) and its data analysis libraries, including Pandas (2.1.1), NumPy (1.25.2), SciPy (1.11.3), scikit-survival (0.23.0), scikit-learn (1.5.2), and PyTorch (2.0.1).

## Data Availability

Of the five data sources used in this study, four datasets are publicly available and can be accessed at the following links: SHHS (https://sleepdata.org/datasets/shhs), MrOS (https://sleepdata.org/ datasets/mros), MESA (https://sleepdata.org/datasets/mesa), and SSC (https://sleepdata.org/ datasets/ssc). The BioSerenity dataset is proprietary and owned by BioSerenity, which has granted Stanford University access under a research and development agreement; please contact BioSerenity directly for data agreement. Stanford sleep data is available upon publication at this link: https://bdsp.io/content/hsp/ 2.0/. Access to these data is provided solely for research purposes and is subject to data use restrictions that prohibit redistribution or sharing with third parties.

## Code Availability

All of the SleepFM code is open source and available at https://github.com/zou-group/sleepfm-clinical

## Extended Tables and Figures

**Extended Data Table 1.**
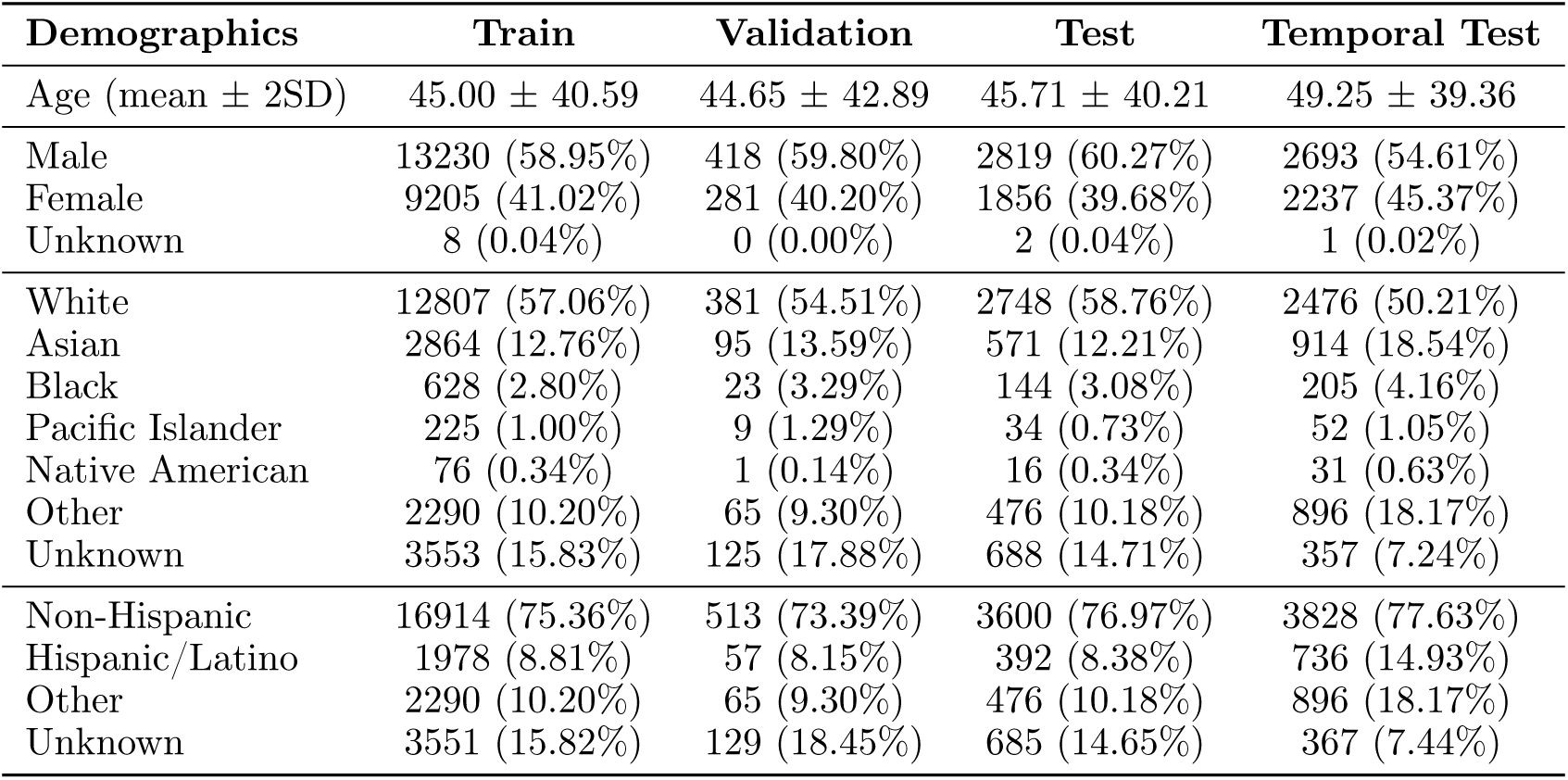
Demographic characteristics of the Stanford Sleep Clinic (SSC) cohort. Values are shown by dataset split and reported as mean ± 2 standard deviations for age, and as counts (percentages) for categorical variables.

| Demographics | Train | Validation | Test | Temporal Test |
| --- | --- | --- | --- | --- |
| Age (mean ± 2SD) | 45.00 ± 40.59 | 44.65 ± 42.89 | 45.71 ± 40.21 | 49.25 ± 39.36 |
| Male | 13230 (58.95%) | 418 (59.80%) | 2819 (60.27%) | 2693 (54.61%) |
| Female | 9205 (41.02%) | 281 (40.20%) | 1856 (39.68%) | 2237 (45.37%) |
| Unknown | 8 (0.04%) | 0 (0.00%) | 2 (0.04%) | 1 (0.02%) |
| White | 12807 (57.06%) | 381 (54.51%) | 2748 (58.76%) | 2476 (50.21%) |
| Asian | 2864 (12.76%) | 95 (13.59%) | 571 (12.21%) | 914 (18.54%) |
| Black | 628 (2.80%) | 23 (3.29%) | 144 (3.08%) | 205 (4.16%) |
| Pacific Islander | 225 (1.00%) | 9 (1.29%) | 34 (0.73%) | 52 (1.05%) |
| Native American | 76 (0.34%) | 1 (0.14%) | 16 (0.34%) | 31 (0.63%) |
| Other | 2290 (10.20%) | 65 (9.30%) | 476 (10.18%) | 896 (18.17%) |
| Unknown | 3553 (15.83%) | 125 (17.88%) | 688 (14.71%) | 357 (7.24%) |
| Non-Hispanic | 16914 (75.36%) | 513 (73.39%) | 3600 (76.97%) | 3828 (77.63%) |
| Hispanic/Latino | 1978 (8.81%) | 57 (8.15%) | 392 (8.38%) | 736 (14.93%) |
| Other | 2290 (10.20%) | 65 (9.30%) | 476 (10.18%) | 896 (18.17%) |
| Unknown | 3551 (15.82%) | 129 (18.45%) | 685 (14.65%) | 367 (7.44%) |

**Extended Data Table 2.**
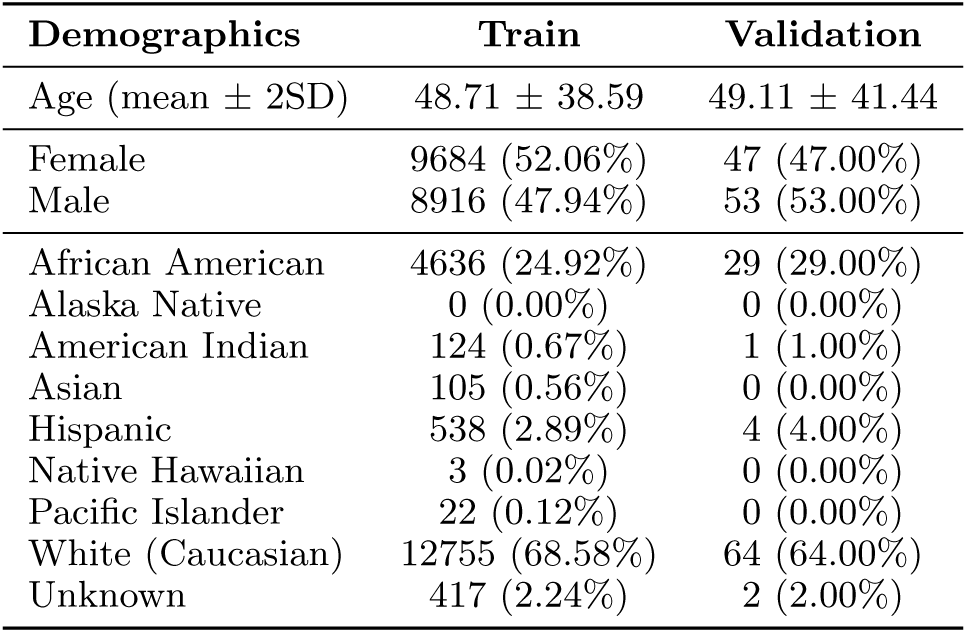
Demographic characteristics of the Bioserenity cohort. Values are presented as mean ± 2 standard deviations for age, and as counts (percentages) for categorical variables.

| Demographics | Train | Validation |
| --- | --- | --- |
| Age (mean ± 2SD) | 48.71 ± 38.59 | 49.11 ± 41.44 |
| Female | 9684 (52.06%) | 47 (47.00%) |
| Male | 8916 (47.94%) | 53 (53.00%) |
| African American | 4636 (24.92%) | 29 (29.00%) |
| Alaska Native | 0 (0.00%) | 0 (0.00%) |
| American Indian | 124 (0.67%) | 1 (1.00%) |
| Asian | 105 (0.56%) | 0 (0.00%) |
| Hispanic | 538 (2.89%) | 4 (4.00%) |
| Native Hawaiian | 3 (0.02%) | 0 (0.00%) |
| Pacific Islander | 22 (0.12%) | 0 (0.00%) |
| White (Caucasian) | 12755 (68.58%) | 64 (64.00%) |
| Unknown | 417 (2.24%) | 2 (2.00%) |

**Extended Data Table 3.** Per–sleep-stage F1 performance of SleepFM across four cohorts. Per–sleep-stage F1 scores for SleepFM across four cohorts (SSC, MESA, MrOS and SHHS), with comparison to U-Sleep. Values for SleepFM are mean F1 with 95% confidence intervals from 1,000 bootstrap resamples of test recordings (CIs shown in parentheses beneath each estimate). The bold “Mean” row reports the macro-average across stages. U-Sleep values are the corresponding F1 scores reported for the same cohorts; confidence intervals were not available.

| Stage | SleepFM |  |  |  | U-Sleep |  |  |  |
| --- | --- | --- | --- | --- | --- | --- | --- | --- |
|  | SSC | MESA | MrOS | SHHS | SSC | MESA | MrOS | SHHS |
| Wake | 0.92<sub>(0.92, 0.93)</sub> | 0.94<sub>(0.94, 0.94)</sub> | 0.94<sub>(0.94, 0.94)</sub> | 0.92<sub>(0.92, 0.92)</sub> | 0.81 | 0.92 | 0.93 | 0.93 |
| Stage 1 | 0.48<sub>(0.48, 0.48)</sub> | 0.56<sub>(0.56, 0.56)</sub> | 0.41<sub>(0.40, 0.41)</sub> | 0.49<sub>(0.48, 0.49)</sub> | 0.35 | 0.59 | 0.46 | 0.51 |
| Stage 2 | 0.87<sub>(0.87, 0.87)</sub> | 0.83<sub>(0.83, 0.83)</sub> | 0.86<sub>(0.86, 0.86)</sub> | 0.84<sub>(0.84, 0.84)</sub> | 0.75 | 0.87 | 0.87 | 0.87 |
| Stage 3 | 0.39<sub>(0.38, 0.39)</sub> | 0.68<sub>(0.68, 0.69)</sub> | 0.65<sub>(0.64, 0.65)</sub> | 0.72<sub>(0.72, 0.72)</sub> | 0.51 | 0.65 | 0.68 | 0.76 |
| REM | 0.86<sub>(0.86, 0.86)</sub> | 0.88<sub>(0.88, 0.88)</sub> | 0.90<sub>(0.90, 0.90)</sub> | 0.91<sub>(0.91, 0.91)</sub> | 0.84 | 0.90 | 0.88 | 0.92 |
| **Mean** | **0.70** | **0.78** | **0.75** | **0.78** | **0.65** | **0.79** | **0.77** | **0.80** |

**Extended Data Table 4.** Sleep staging performance on the SSC cohort. Sleep staging results (F1) for SleepFM, U-Sleep, YASA, GSSC, and STAGES on the SSC cohort. The **bold** bottom row reports the macro-average (mean) across stages.

| Stage | SleepFM | U-Sleep | YASA | GSSC | STAGES |
| --- | --- | --- | --- | --- | --- |
| Wake | 0.92 | 0.81 | 0.61 | 0.84 | 0.73 |
| Stage 1 | 0.48 | 0.35 | 0.05 | 0.39 | 0.33 |
| Stage 2 | 0.87 | 0.75 | 0.69 | 0.79 | 0.82 |
| Stage 3 | 0.39 | 0.51 | 0.56 | 0.56 | 0.63 |
| REM | 0.86 | 0.84 | 0.71 | 0.85 | 0.79 |
| **Mean** | **0.70** | **0.65** | **0.52** | **0.68** | **0.66** |

**Extended Data Table 5.** Comparison of category-averaged C-Index across SleepFM variants and baselines. Category-averaged C-Index (mean with 95% CI) comparing SleepFM variants with two baselines across disease categories on Stanford cohort (n=5,019). The Demographics baseline (“Demo”) uses only structured clinical features (age, gender, BMI, and race/ethnicity). The End-to-End PSG baseline (“E2E-PSG”) is trained directly on raw PSG signals with age and gender, without any pretraining. SleepFM-1 denotes SleepFM-LSTM (w/o Demo), using two LSTM layers in the fine-tuning prediction module and no demographic features. SleepFM-2 denotes SleepFM-Linear, a linear prediction module on SleepFM embeddings with age and gender. SleepFM-3 denotes SleepFM-LSTM, which uses two LSTM layers in the fine-tuning prediction module with age and gender. Values are averaged within each category across conditions. Uncertainty is estimated via nonparametric bootstrapping (n=1000 resamples): for each resample, conditions within a category are sampled with replacement and the category mean is computed; 95% CIs are the 2.5th–97.5th percentiles across resamples.

| Category | Demo | E2E PSG | SleepFM-1 | SleepFM-2 | SleepFM-3 |
| --- | --- | --- | --- | --- | --- |
| Circulatory System | 0.72<sub>(0.72, 0.73)</sub> | 0.72<sub>(0.72, 0.73)</sub> | 0.75<sub>(0.75, 0.75)</sub> | 0.76<sub>(0.75, 0.76)</sub> | 0.75<sub>(0.75, 0.76)</sub> |
| Dermatologic | 0.63<sub>(0.62, 0.64)</sub> | 0.62<sub>(0.61, 0.63)</sub> | 0.65<sub>(0.64, 0.65)</sub> | 0.66<sub>(0.65, 0.66)</sub> | 0.66<sub>(0.65, 0.66)</sub> |
| Digestive | 0.62<sub>(0.62, 0.63)</sub> | 0.63<sub>(0.62, 0.63)</sub> | 0.66<sub>(0.65, 0.67)</sub> | 0.67<sub>(0.66, 0.68)</sub> | 0.67<sub>(0.66, 0.67)</sub> |
| Endocrine/Metabolic | 0.68<sub>(0.67, 0.68)</sub> | 0.66<sub>(0.66, 0.67)</sub> | 0.70<sub>(0.70, 0.71)</sub> | 0.71<sub>(0.71, 0.72)</sub> | 0.71<sub>(0.71, 0.72)</sub> |
| Hematopoietic | 0.65<sub>(0.64, 0.66)</sub> | 0.66<sub>(0.65, 0.67)</sub> | 0.70<sub>(0.69, 0.71)</sub> | 0.70<sub>(0.69, 0.72)</sub> | 0.70<sub>(0.69, 0.71)</sub> |
| Infectious Diseases | 0.62<sub>(0.61, 0.63)</sub> | 0.61<sub>(0.60, 0.62)</sub> | 0.65<sub>(0.64, 0.66)</sub> | 0.66<sub>(0.64, 0.67)</sub> | 0.66<sub>(0.64, 0.67)</sub> |
| Injuries & Poisonings | 0.62<sub>(0.61, 0.63)</sub> | 0.63<sub>(0.62, 0.64)</sub> | 0.65<sub>(0.64, 0.66)</sub> | 0.66<sub>(0.65, 0.67)</sub> | 0.66<sub>(0.65, 0.67)</sub> |
| Mental Disorders | 0.66<sub>(0.65, 0.66)</sub> | 0.66<sub>(0.65, 0.67)</sub> | 0.69<sub>(0.68, 0.70)</sub> | 0.69<sub>(0.69, 0.70)</sub> | 0.70<sub>(0.70, 0.71)</sub> |
| Musculoskeletal | 0.67<sub>(0.66, 0.67)</sub> | 0.67<sub>(0.66, 0.68)</sub> | 0.67<sub>(0.67, 0.68)</sub> | 0.68<sub>(0.67, 0.69)</sub> | 0.68<sub>(0.67, 0.69)</sub> |
| Neoplasms | 0.72<sub>(0.71, 0.73)</sub> | 0.72<sub>(0.71, 0.73)</sub> | 0.71<sub>(0.70, 0.72)</sub> | 0.73<sub>(0.72, 0.74)</sub> | 0.73<sub>(0.72, 0.74)</sub> |
| Neurological | 0.62<sub>(0.61, 0.63)</sub> | 0.63<sub>(0.62, 0.63)</sub> | 0.66<sub>(0.66, 0.67)</sub> | 0.67<sub>(0.66, 0.67)</sub> | 0.67<sub>(0.66, 0.68)</sub> |
| Respiratory | 0.63<sub>(0.62, 0.64)</sub> | 0.63<sub>(0.63, 0.64)</sub> | 0.66<sub>(0.65, 0.66)</sub> | 0.66<sub>(0.65, 0.66)</sub> | 0.66<sub>(0.65, 0.67)</sub> |
| Sense Organs | 0.65<sub>(0.65, 0.66)</sub> | 0.66<sub>(0.66, 0.67)</sub> | 0.68<sub>(0.67, 0.68)</sub> | 0.68<sub>(0.67, 0.68)</sub> | 0.68<sub>(0.67, 0.68)</sub> |
| Symptoms | 0.64<sub>(0.63, 0.64)</sub> | 0.64<sub>(0.64, 0.65)</sub> | 0.68<sub>(0.67, 0.69)</sub> | 0.69<sub>(0.68, 0.69)</sub> | 0.68<sub>(0.68, 0.69)</sub> |

**Extended Data Figure 1.**
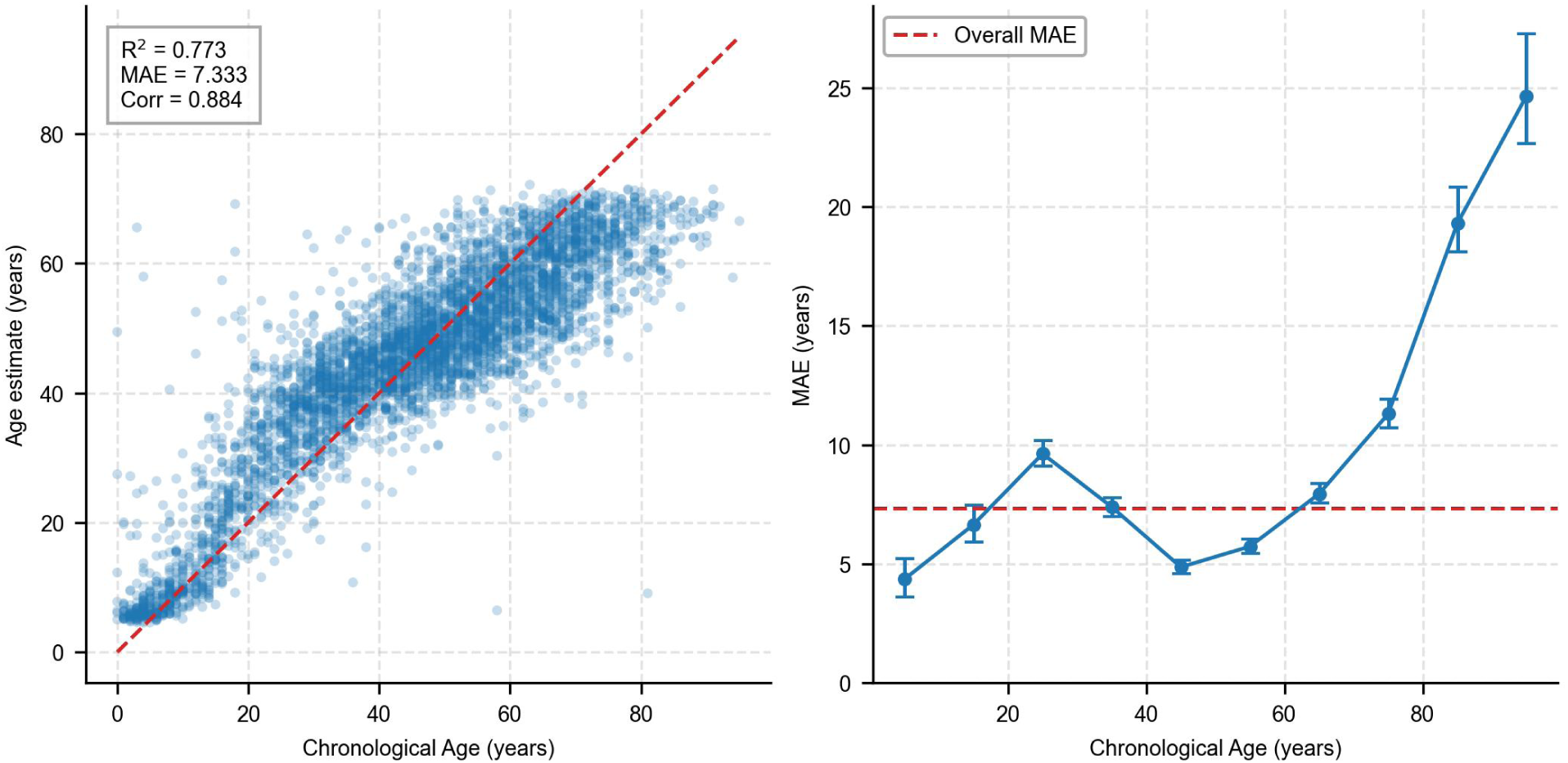
Age estimation performance on the Stanford cohort. Left: Scatter plot showing predicted versus chronological age across all patients (n = 5,019), with the diagonal line representing perfect prediction. The coefficient of determination (R^2^), mean absolute error (MAE), and Pearson correlation coefficient (Corr) are shown in the top left corner. Right: Mean Absolute Error (MAE) across chronological age groups, with vertical error bars indicating the standard error of the mean (SEM) within each age bin. The horizontal dashed line represents the overall MAE. Our model achieves an MAE comparable to state-of-the-art models and demonstrates improved age estimation performance for younger age groups compared to older ones.

**Extended Data Figure 2.**
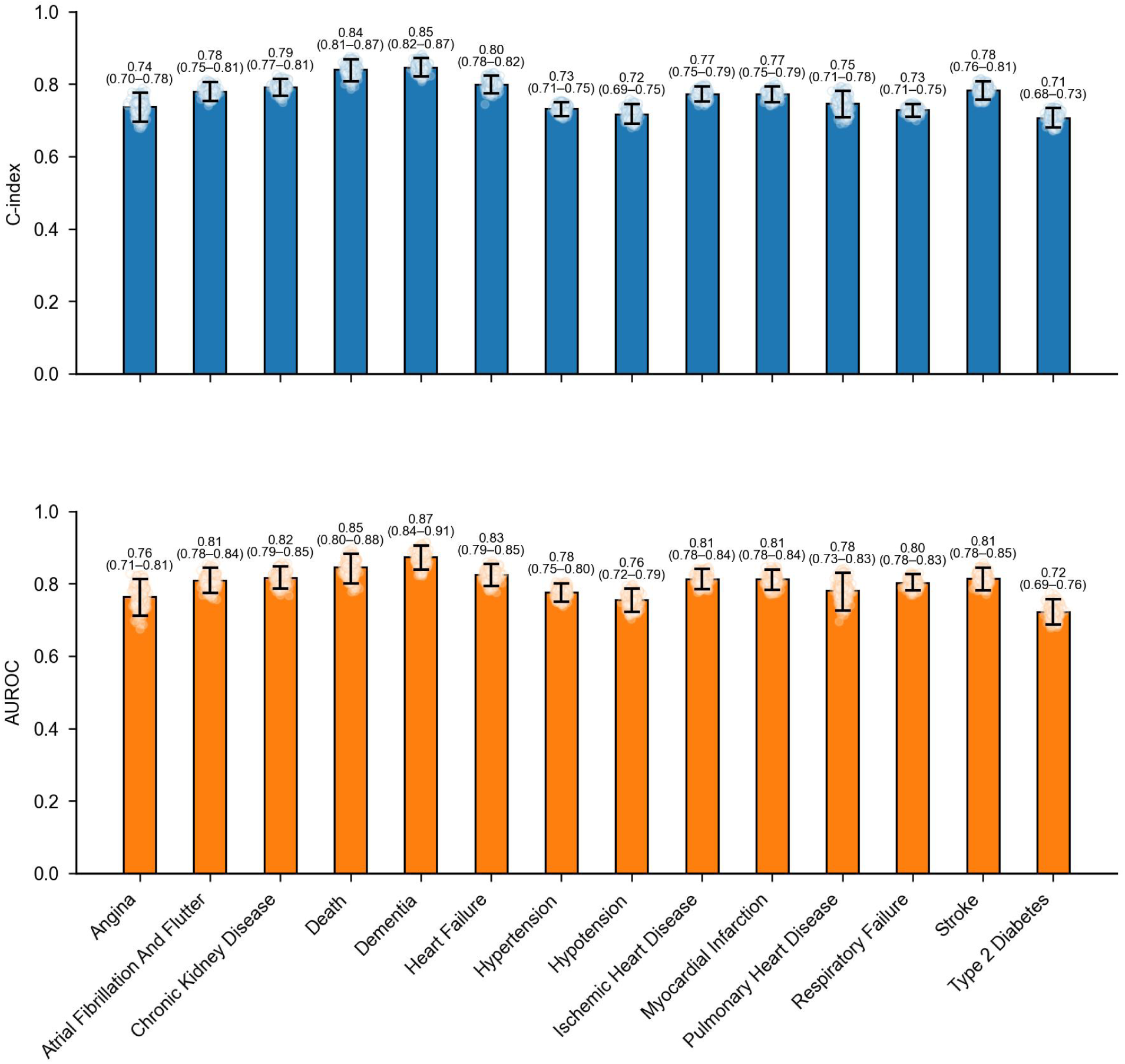
Performance across clinically relevant diseases evaluated on the Stanford test set (n=5019). Performance is evaluated using multiple metrics: C-Index and AUROC. The selected conditions include critical health outcomes such as death, heart failure, stroke, and dementia. Each panel uses violin/point plots derived from 1000 patient-level bootstrapping: the violin encodes the distribution of bootstrap estimates, faint points are individual bootstrap draws, the filled dot is the mean, and the vertical line with end caps marks the 95% bootstrap percentile CI. Numbers above violins report the mean. Metrics are C-index (top) and AUROC at 6 years (bottom).

**Extended Data Figure 3.**
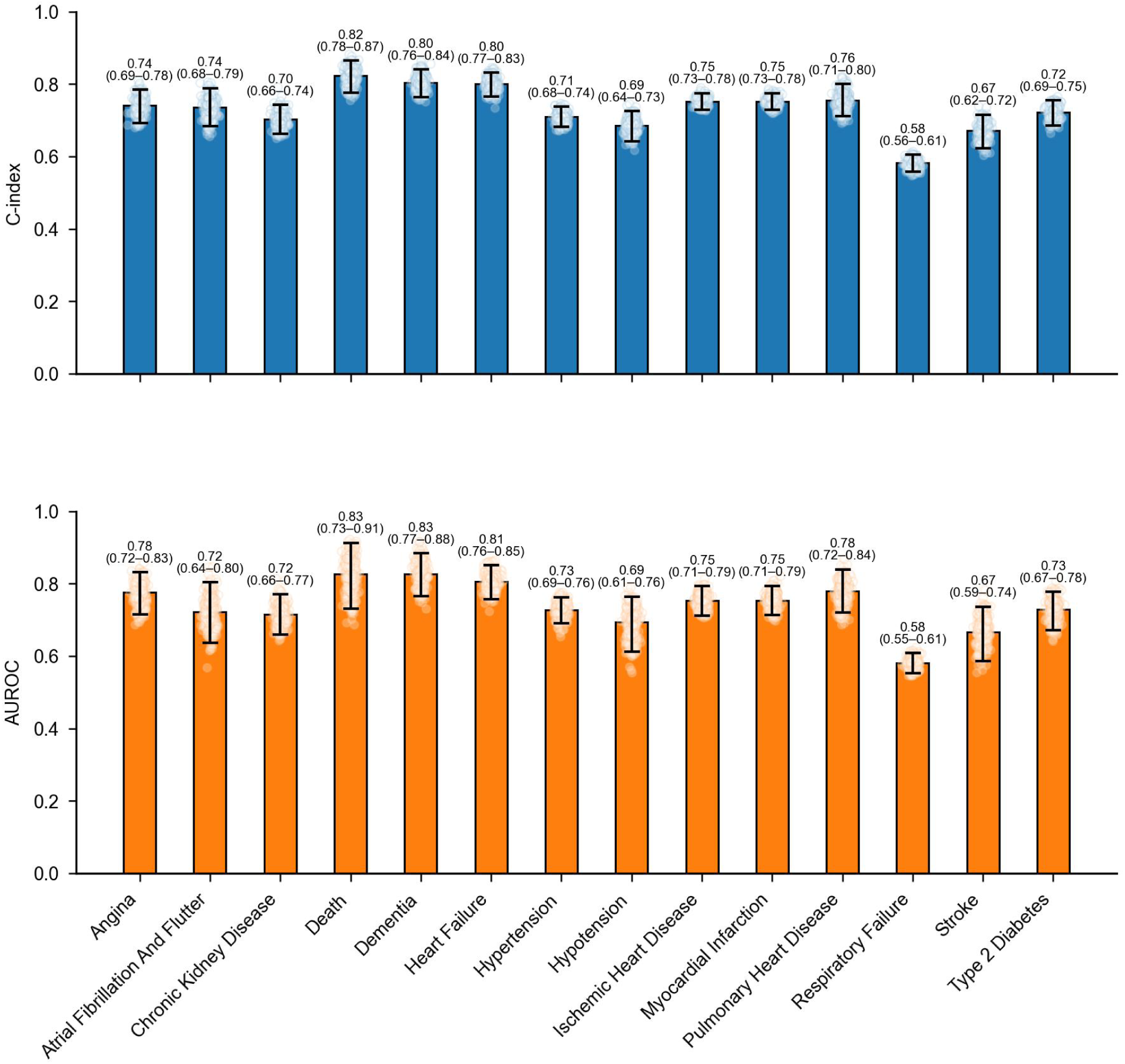
Prediction performance of SleepFM for key clinical outcomes on the temporal test set, including C-Index and AUROC metrics for critical conditions such as death, heart failure, chronic kidney disease, dementia, and stroke. Each panel uses violin/point plots derived from 1000 patient-level bootstrapping: the violin encodes the distribution of bootstrap estimates, faint points are individual bootstrap draws, the filled dot is the mean, and the vertical line with end caps marks the 95% bootstrap percentile CI. Numbers above violins report the mean. Metrics are C-index (top) and AUROC (bottom). All conditions are statistically significant with a p-value < 0.01 after Bonferroni correction.

**Extended Data Figure 4.**
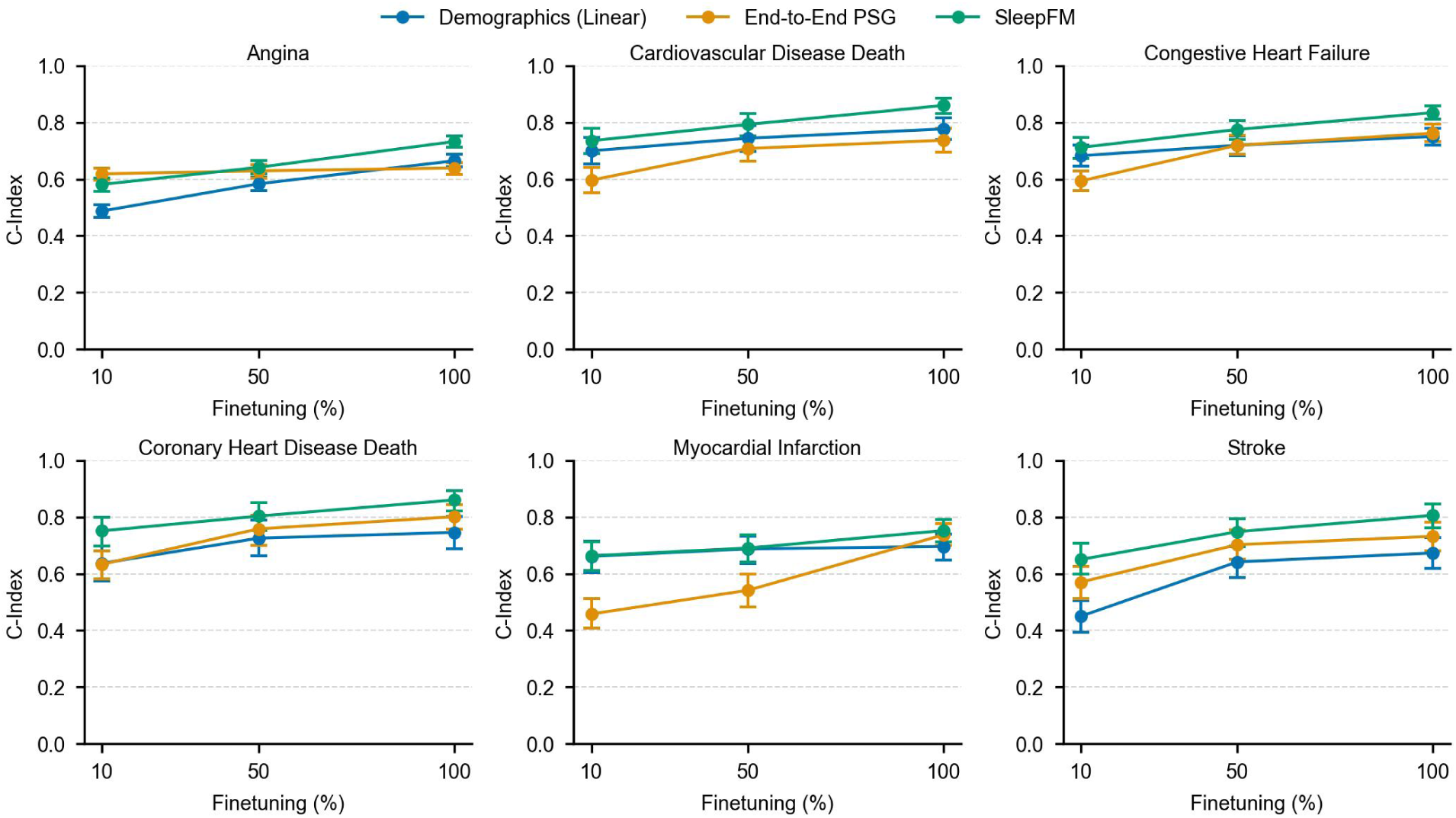
Scaling behavior of fine-tuning SleepFM on the SHHS dataset. Scaling behavior of fine-tuning SleepFM on the SHHS dataset (test size = 2,000 participants). We progressively increased the percentage of labeled SHHS data used during fine-tuning from 10% to 100%. The plots show C-Index performance across six cardiovascular outcomes, comparing SleepFM with Demographics and End-to-End PSG baselines. Error bars indicate 95% confidence intervals derived from 1,000 participant-level bootstrap resamples with replacement. Even with as little as 10% of training data (∼330 samples), SleepFM demonstrates strong predictive accuracy and consistent performance improvements as more labeled data becomes available. SleepFM outperforms both baseline models in most conditions, particularly when the dataset size is smaller, and its performance scaling is more stable across all outcomes.

**Extended Data Figure 5.**
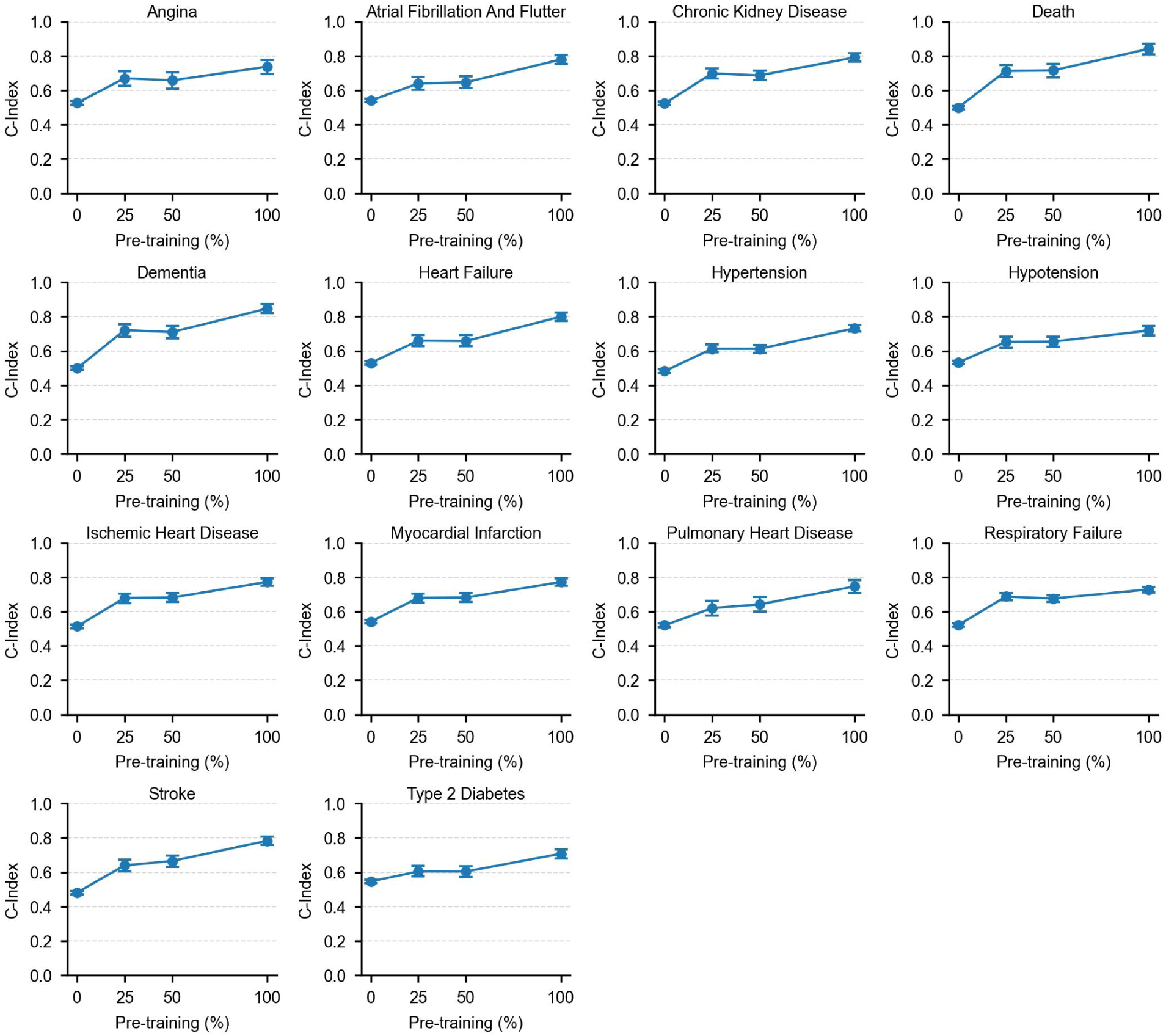
Impact of pretraining dataset size on downstream performance on the Stanford cohort. Each subplot shows C-Index performance for a specific disease as a function of the percentage of pretraining data used (0%, 25%, 50%, 100%). The downstream fine-tuning and test datasets are held constant. Error bars represent 95% confidence intervals estimated via 1,000 participant-level bootstrap resamples with replacement. The 100% mark corresponds to a full epoch of pretraining on the entire dataset (n = 24, 137). Intermediate checkpoints at 25% and 50% represent models saved partway through that epoch, while the 0% point denotes a model with no pretraining, resulting in near-random performance. Performance improves consistently with more pretraining data, highlighting the value of large-scale self-supervised pretraining across diverse phenotypes, including cardiovascular, metabolic, neurological, and respiratory conditions.

## Supplementary Section

**Supplementary Figure 1.**
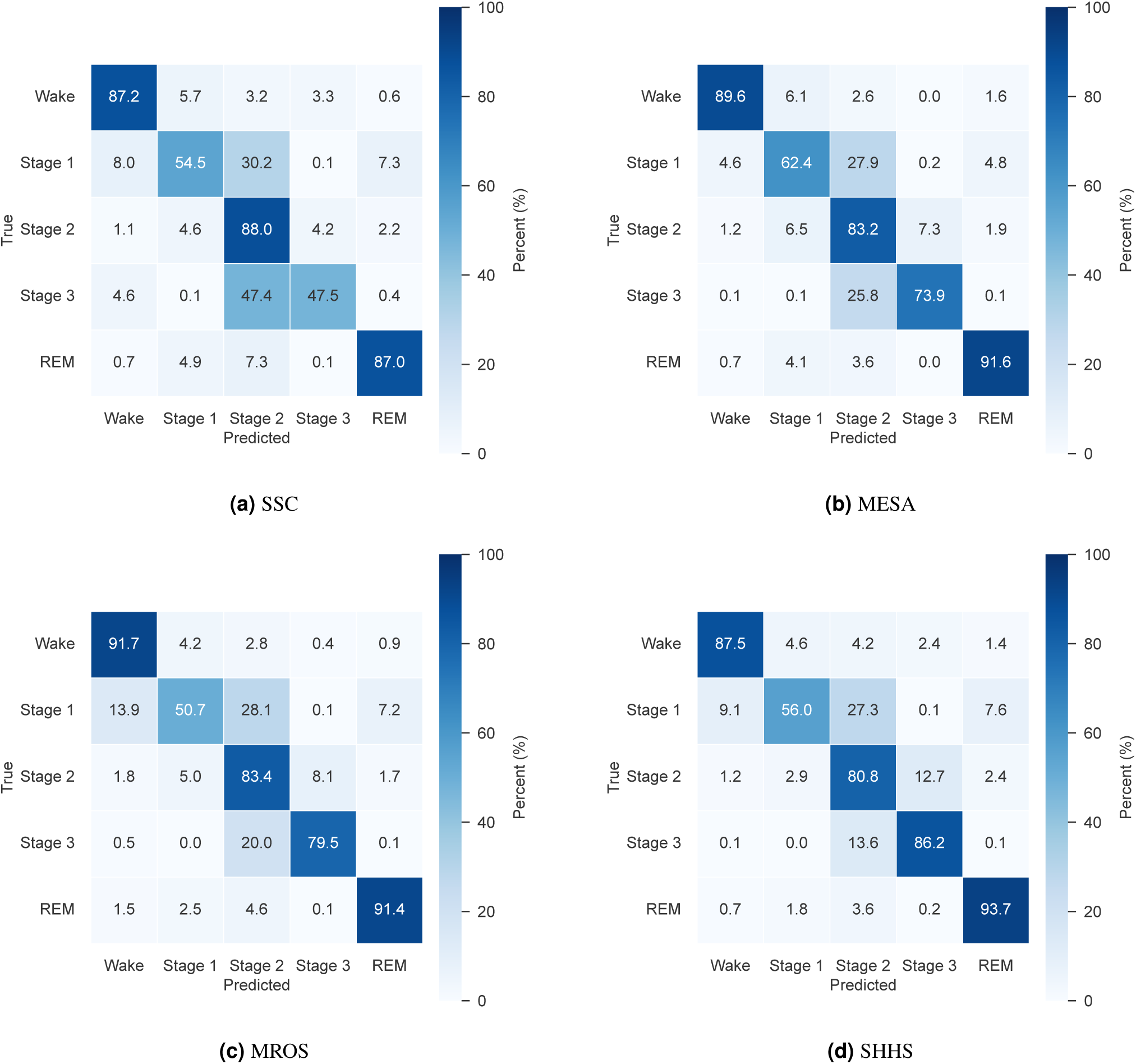
SleepFM sleep staging confusion matrices for the Stanford, MESA, MROS, and SHHS cohorts. SleepFM generally performs well in differentiating between sleep stages, with mild confusion observed between Stage 1 and Stage 2, as well as between Stage 2 and Stage 3.

**Supplementary Figure 2.**
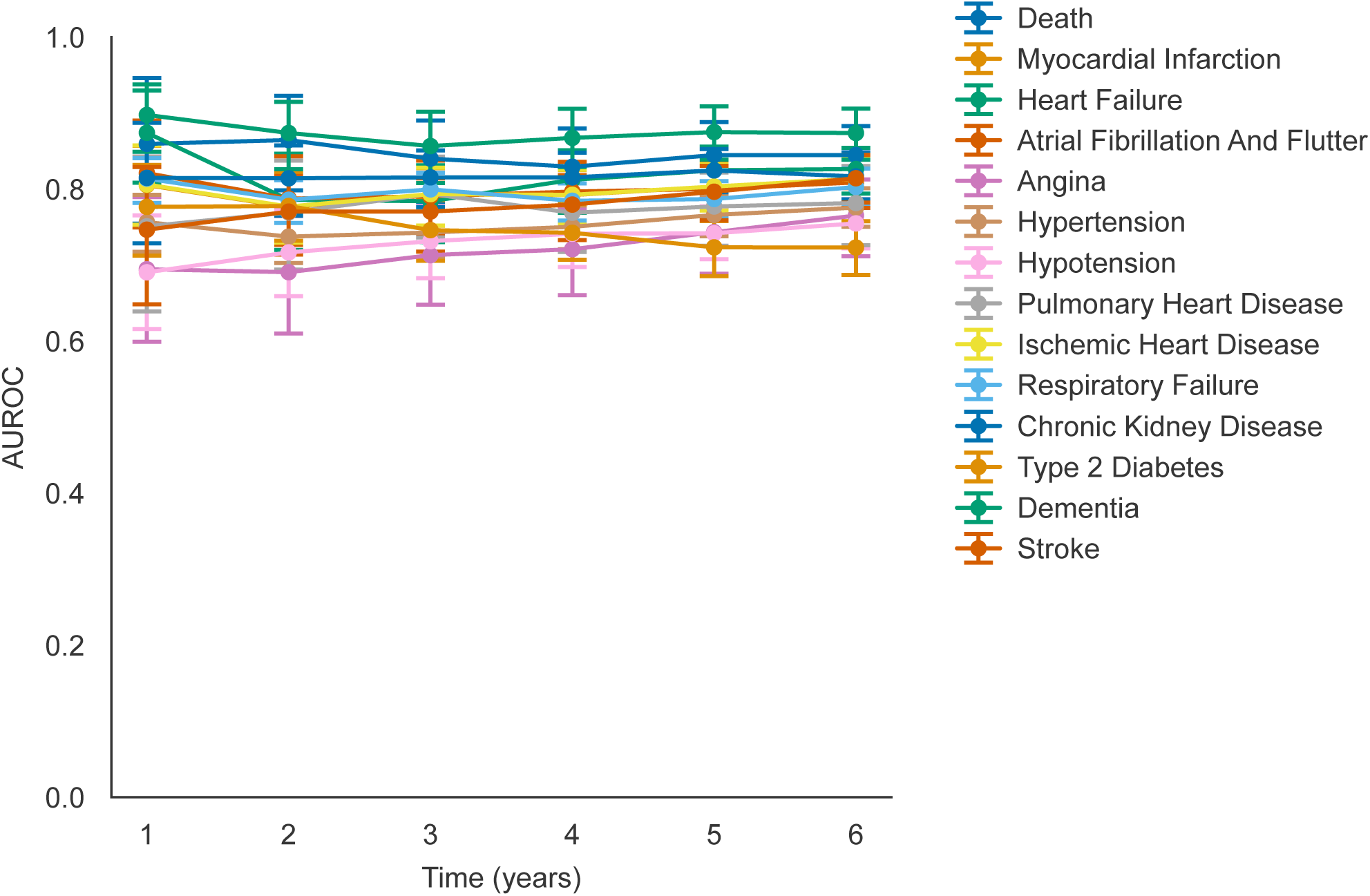
AUROC trends across prediction windows from 1 to 6 years. While we report 6-year AUROC as our primary metric to accommodate the range of conditions—both acute and chronic—we acknowledge that different time horizons may be more appropriate for specific conditions. The plot above demonstrates that AUROC remains robust across multiple conditions over the 1–6 year range.

**Supplementary Figure 3.**
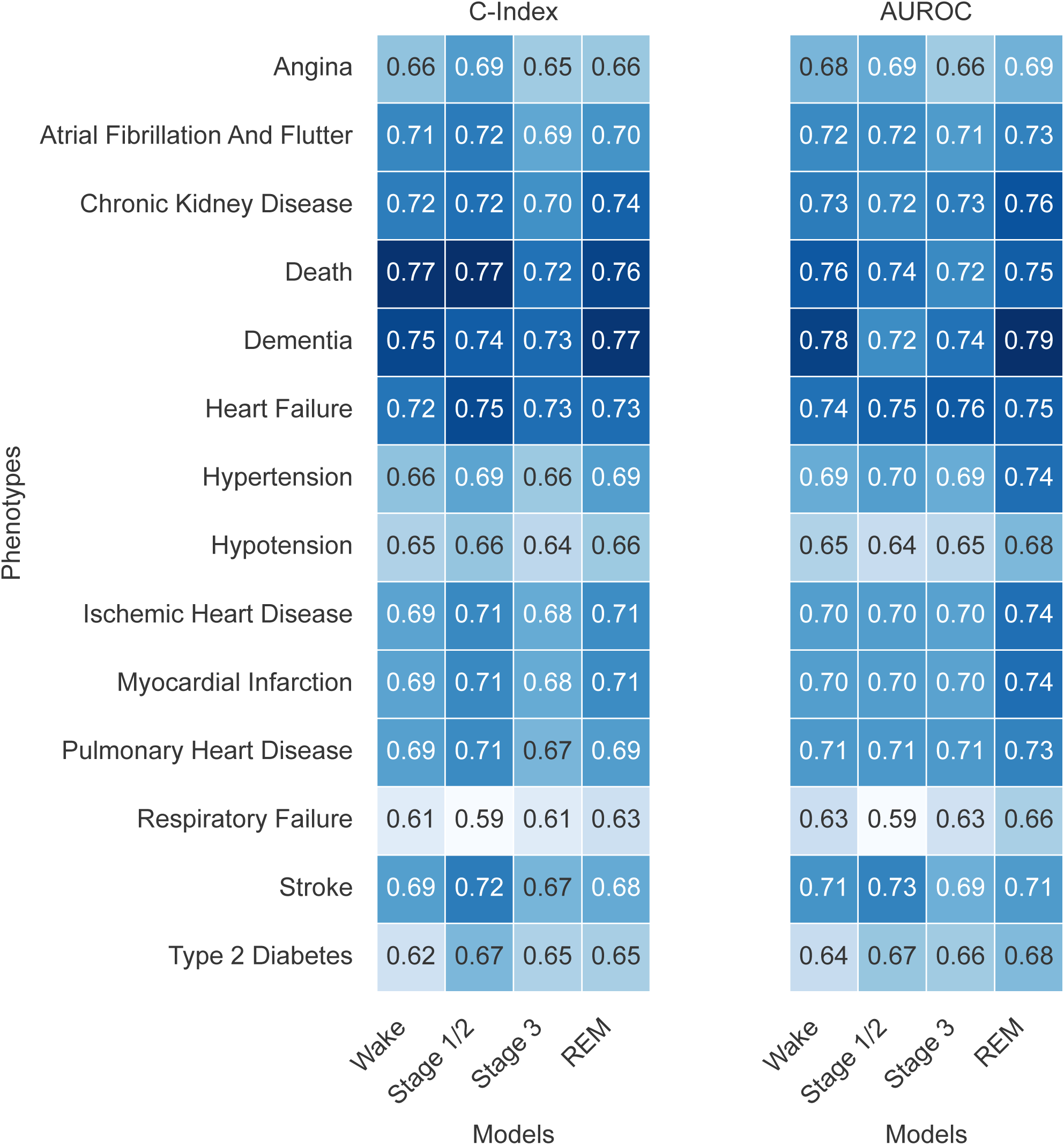
Performance comparison of models trained on different sleep stages across selected conditions. Metrics presented are C-Index and AUROC.

**Supplementary Figure 4.**
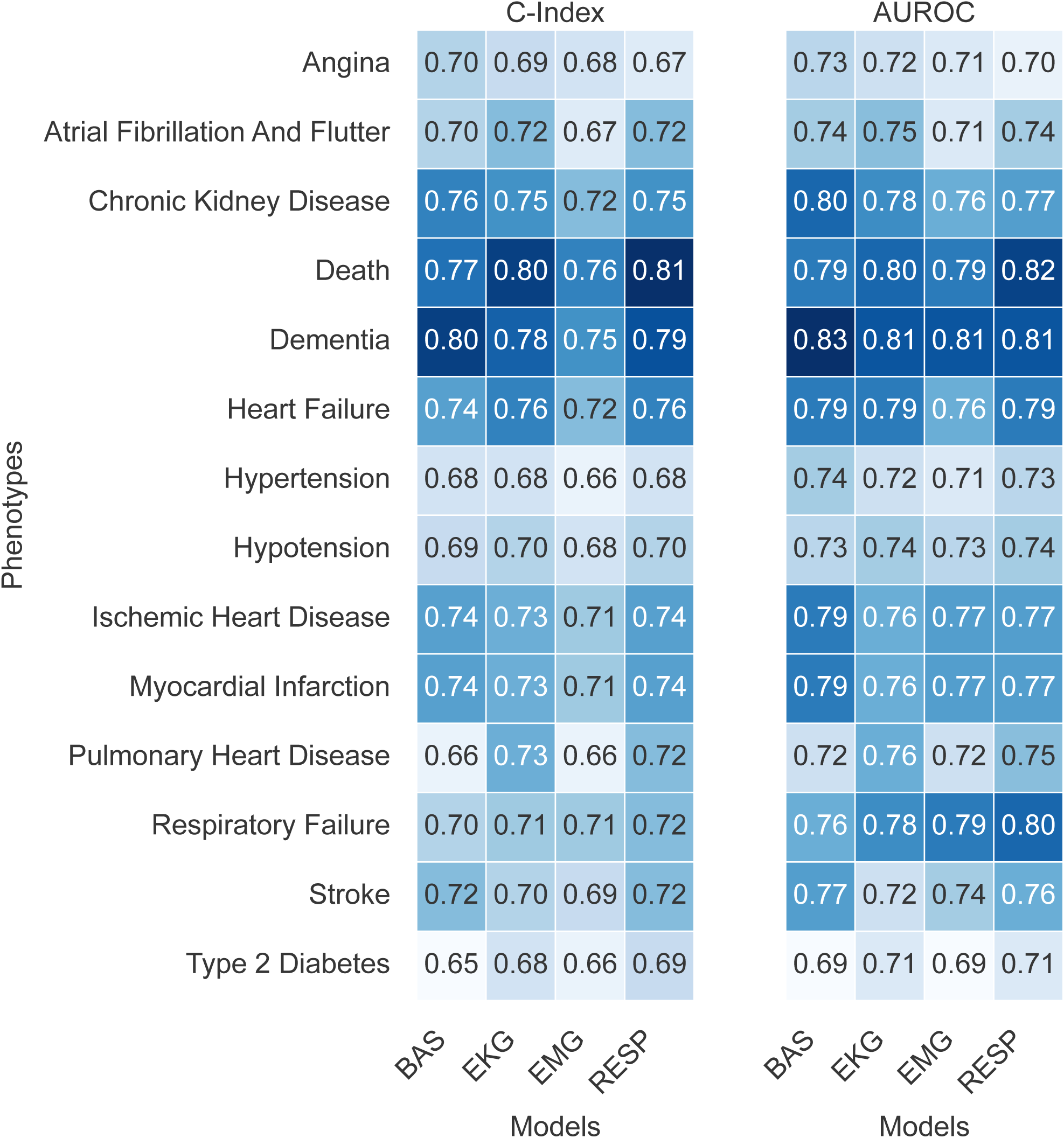
Performance comparison of models trained on different sleep modalities across selected conditions. BAS, EKG, and RESP exhibit similar predictive performance across various conditions, while EMG is the least predictive.

**Supplementary Figure 5.**
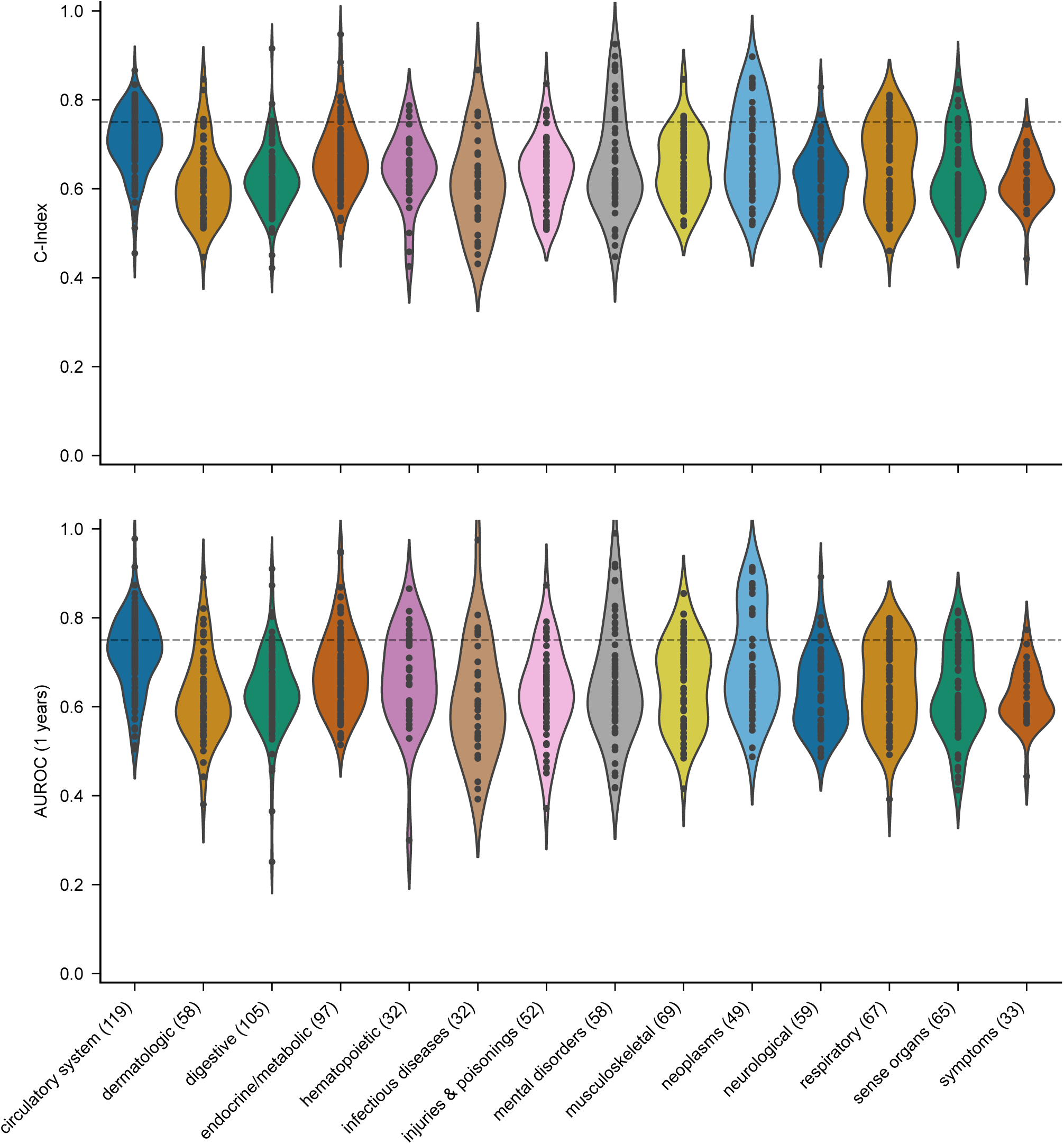
SleepFM performance across disease categories on the temporal test set (Stanford data from 2020 onwards). Performance is measured using C-Index and 6-year AUROC metrics to assess the model’s robustness to temporal distribution shifts. Despite these shifts, SleepFM maintains strong performance across a broad spectrum of diseases.

**Supplementary Figure 6.**
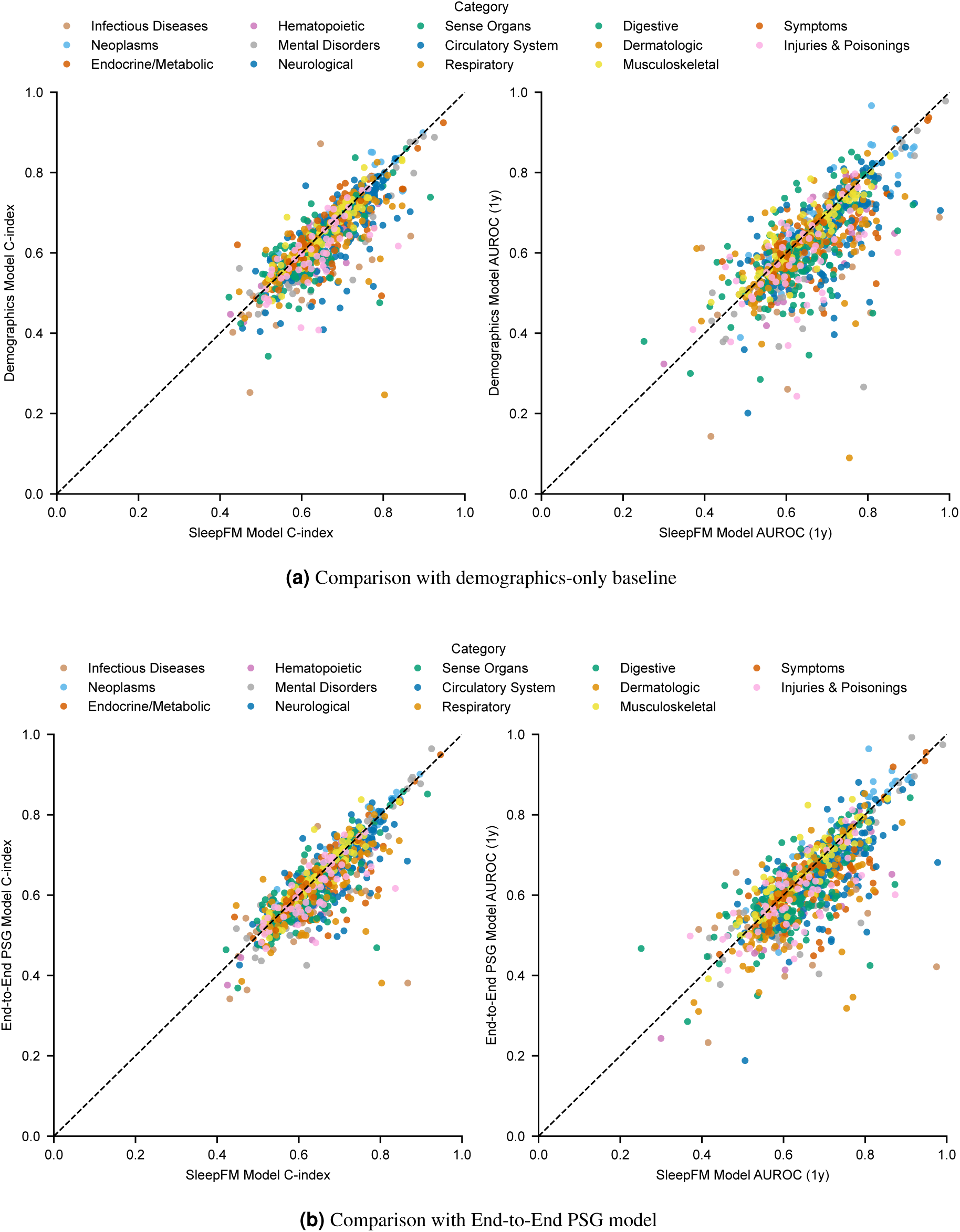
Performance comparison on the temporal test set (Stanford data from 2020 onwards) using C-Index and 6-year AUROC metrics. Each point represents a disease phenotype. (a) Comparison against demographics baseline shows points below the diagonal, indicating SleepFM’s superior performance. (b) Comparison against End-to-End PSG model demonstrates the benefits of foundation model pre-training even under temporal distribution shift. These results highlight SleepFM’s robust generalization to more recent patients. Overall, most points fall below the diagonal, demonstrating that SleepFM outperforms the baseline models for the majority of conditions.

**Supplementary Figure 7.**
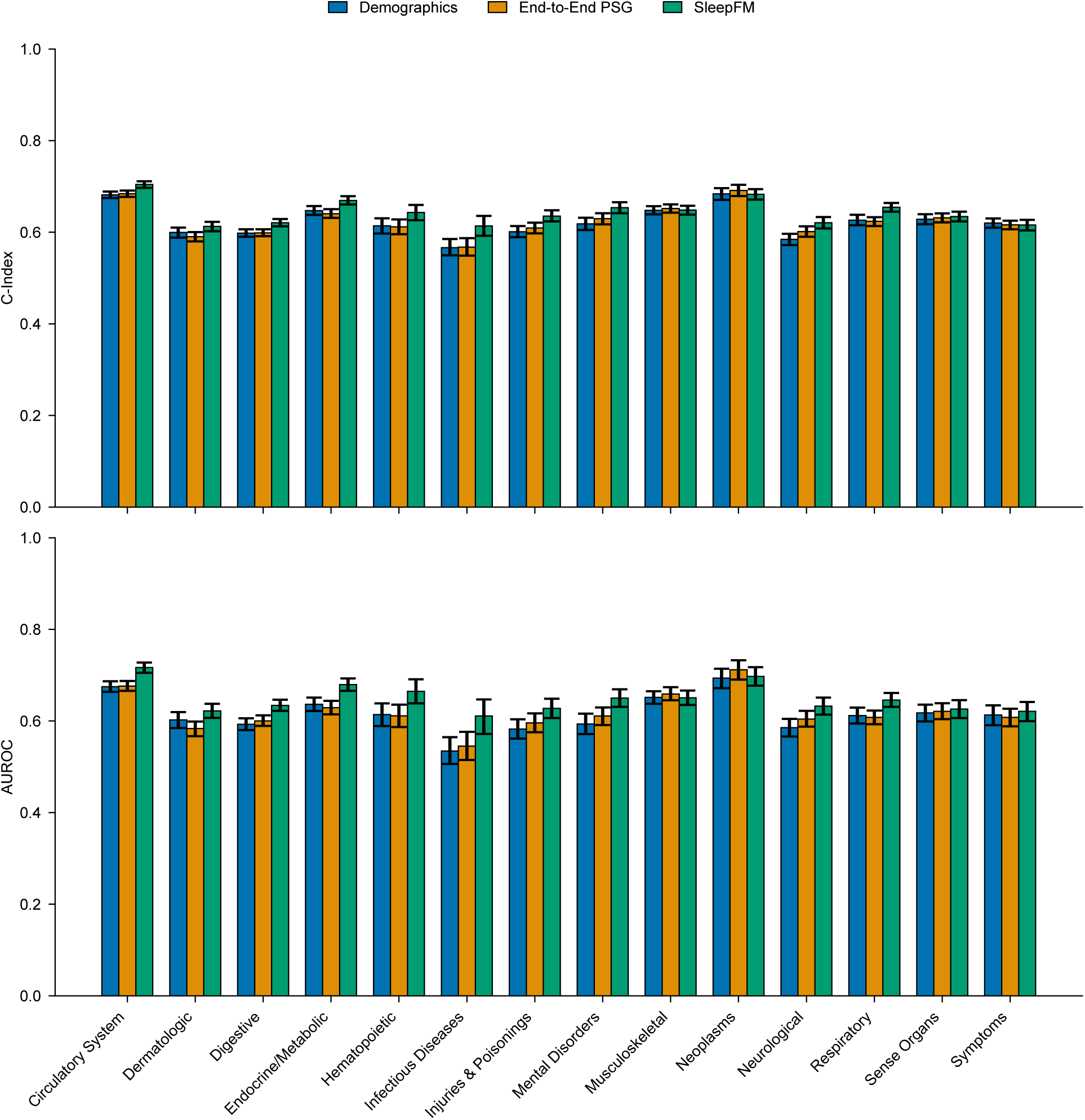
Performance comparison of SleepFM with baseline models on the temporal test set (2020 onwards), showing average C-Index and 6-year AUROC metrics across disease categories. The demographics baseline uses only clinical features (age, gender, BMI, and race/ethnicity), while the End-to-End PSG baseline is trained directly on raw PSG signals without pre-training. Despite the temporal distribution shift, SleepFM maintains its performance advantages over both baselines across most disease categories, demonstrating robust generalization to more recent patients.

**Supplementary Figure 8.**
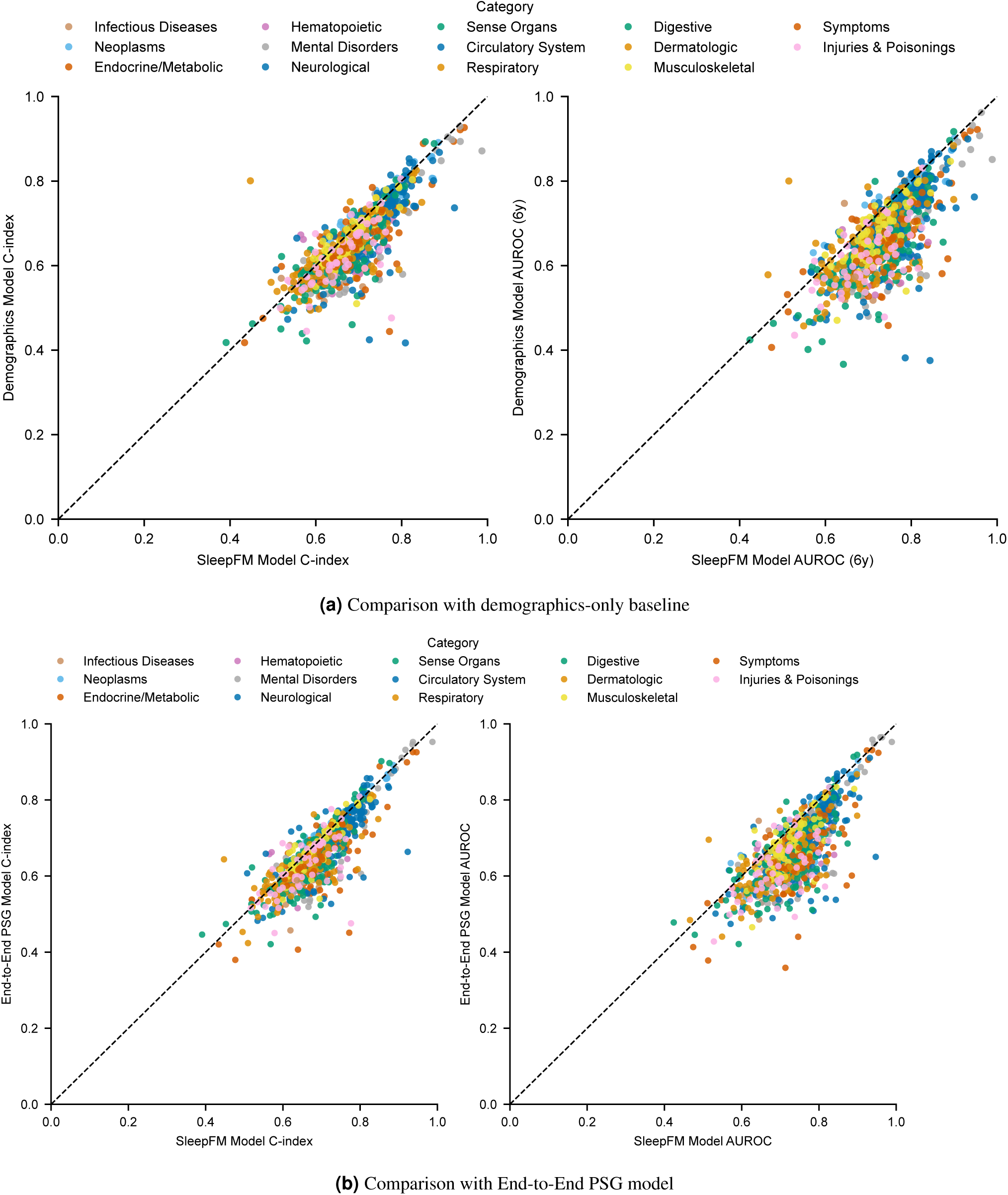
Performance comparison of SleepFM with baseline models on the test set using C-Index and 6-year AUROC metrics. Each point represents a disease phenotype. (a) Comparison against the demographics-only baseline shows points consistently below the diagonal, indicating superior performance by SleepFM. (b) Comparison against the End-to-End PSG model highlights the advantages of foundation model pre-training over direct supervised learning from raw PSG signals. Overall, most points fall below the diagonal, demonstrating that SleepFM outperforms the baseline models for the majority of conditions.

**Supplementary Figure 9.**
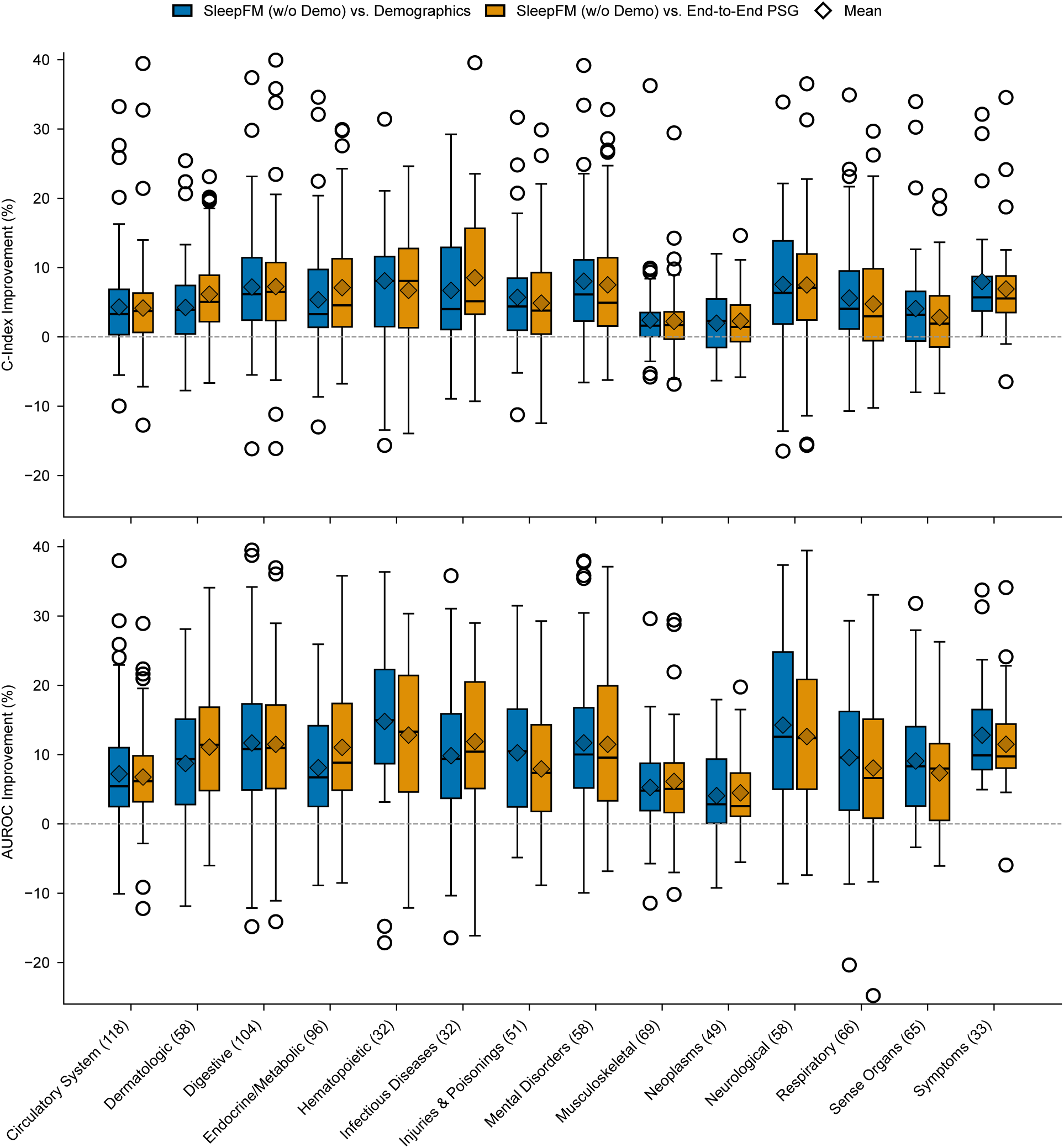
Performance improvements of SleepFM over baseline models across disease categories. This version of SleepFM excludes demographic inputs, while the End-to-End PSG model includes age and sex. Results show percentage improvements in C-Index and 6-year AUROC on the test set. Compared with the demographics-only model (age, sex, BMI, and race/ethnicity), SleepFM achieves broad gains, especially for neurological and hematopoietic conditions. The comparison with the End-to-End PSG model underscores the value of foundation model pre-training for robust sleep representations. Each box shows the distribution of disease-level percentage improvements by category, with C-Index (top) and AUROC (bottom). Boxes indicate interquartile ranges (IQR), whiskers extend to 1.5×IQR, outliers are points, and diamonds mark mean improvements. The dashed line at zero denotes no improvement.

**Supplementary Figure 10.**
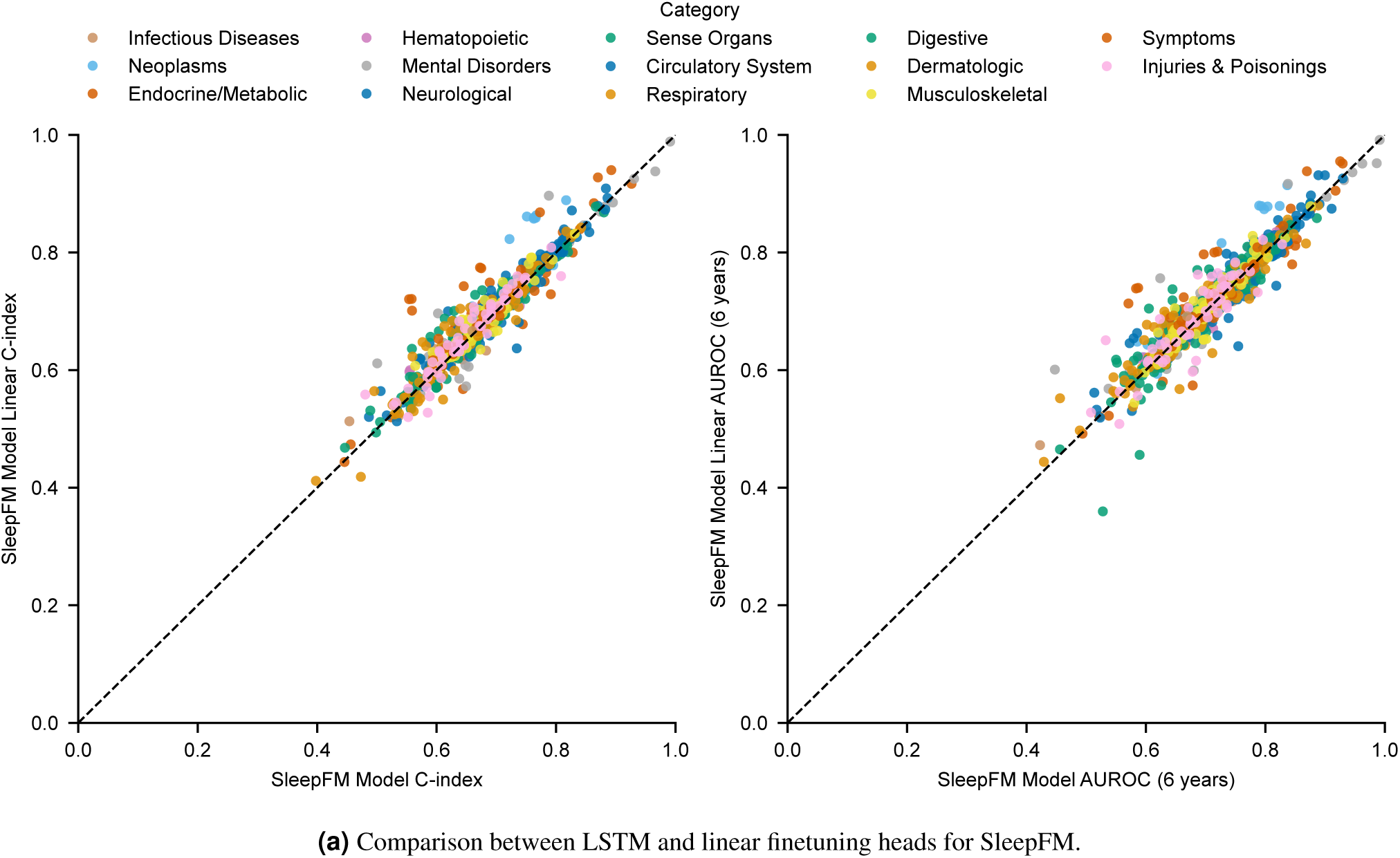
Performance comparison between different finetuning heads for SleepFM using C-Index and 6-year AUROC metrics. Each point represents a disease phenotype. The LSTM-based head shows only a modest advantage over the linear classifier, with an average improvement of 1.98% in C-Index and 1.72% in AUROC. This suggests that the strength of SleepFM lies primarily in the expressiveness of its pretrained embeddings, rather than complexity in the downstream architecture.

**Supplementary Figure 11.**
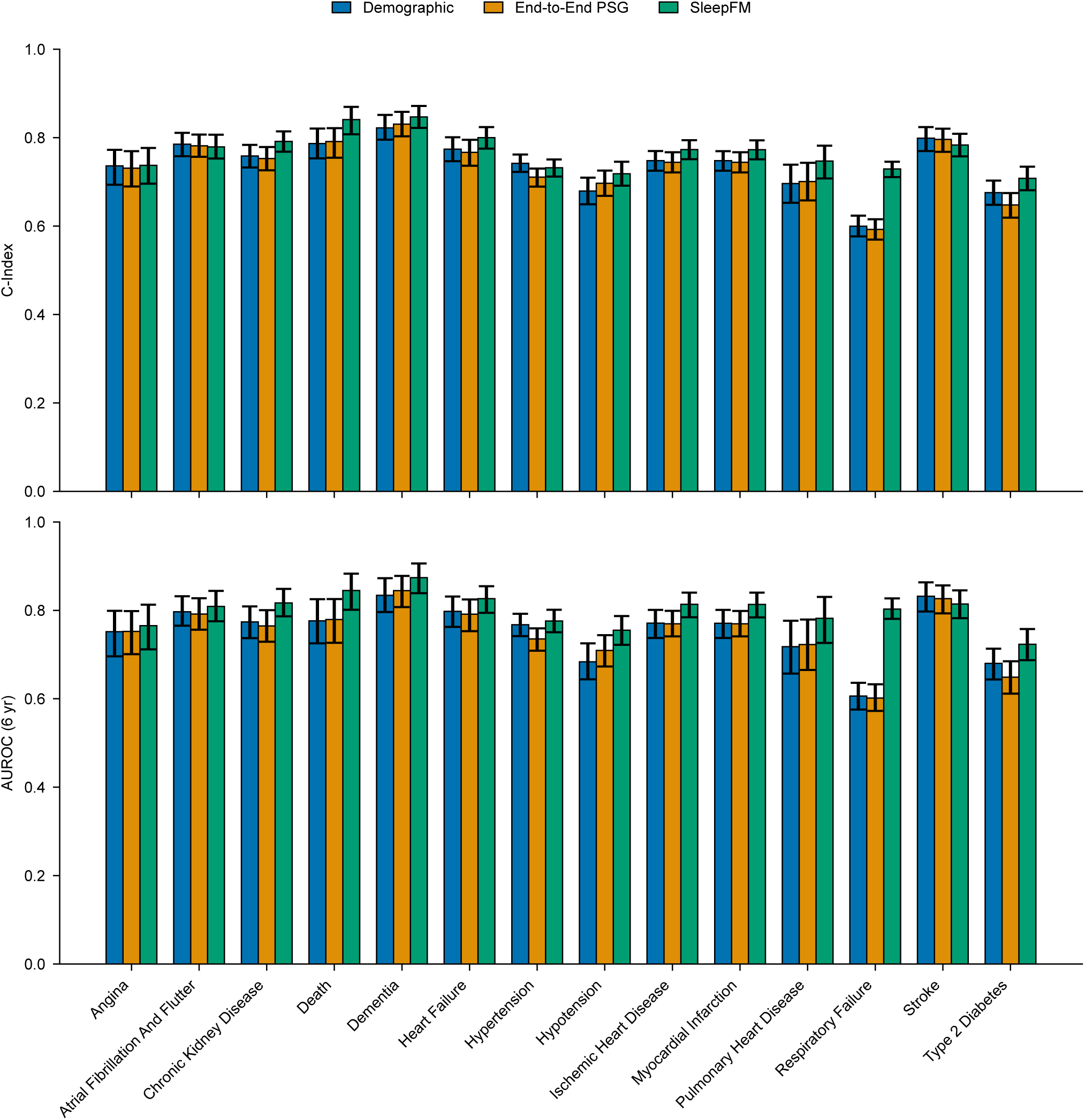
Performance comparison across clinically relevant diseases selected in consultation with sleep experts. The plot compares SleepFM against two baselines: a demographics-only model using clinical features (age, gender, BMI, and race/ethnicity) and an End-to-End PSG model trained directly on raw PSG signals. Performance is evaluated on the test set using multiple metrics: C-Index and 6-year AUROC. The selected conditions include critical health outcomes such as death, heart failure, stroke, and dementia. SleepFM demonstrates superior predictive performance across these important clinical endpoints compared to both baseline approaches.

**Supplementary Figure 12.**
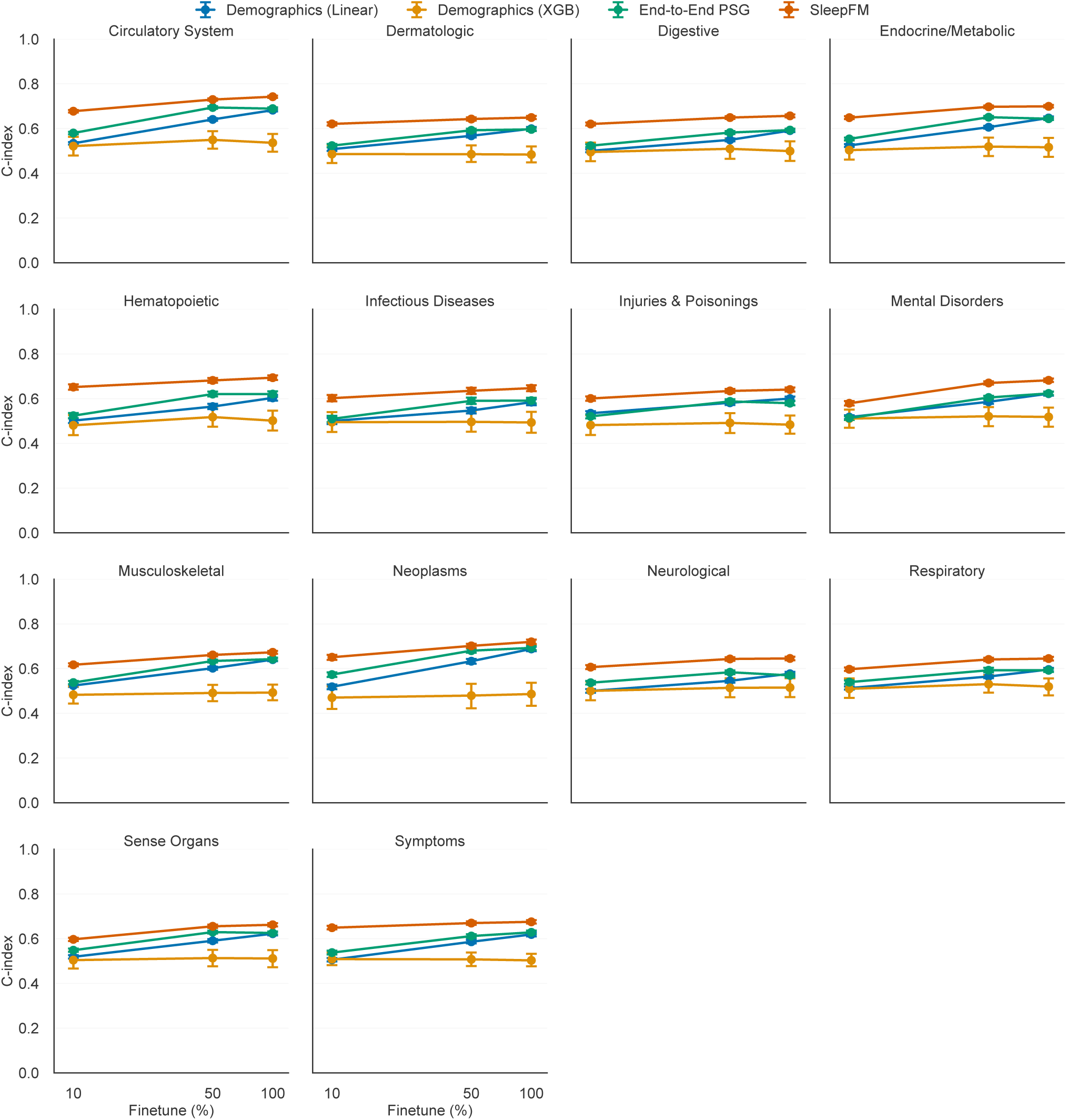
Scaling behavior of fine-tuning SleepFM on the SSC dataset. We progressively increased the proportion of labeled SSC data used during fine-tuning, from 10% to 100%. The plots show C-Index performance across multiple outcome categories. Notably, SleepFM achieves strong predictive performance even with just 10% of the training data, with performance improving consistently as more labeled data is used. Here, 100% corresponds to 6,000 SSC examples—substantially fewer than the full fine-tuning dataset of 24,137 examples. Note that while the same SSC data was used during fine-tuning and seen by the SleepFM model during pretraining, the pretraining objective was entirely different and lacked disease supervision, minimizing potential bias.

**Supplementary Table 1.** F1 scores for SleepFM and PhysioEx models on DCSM and HMC in an external validation setting (no site-specific fine-tuning).

|  | DCSM | HMC |
| --- | --- | --- |
| SeqSleepNet | 0.62 | 0.64 |
| Chambon2018 | 0.43 | 0.65 |
| TinySleepNet | 0.34 | 0.63 |
| SleepFM | 0.68 | 0.55 |

**Supplementary Table 2.** Sleep staging results (macro F1 score) when pretrained on different cohorts, finetuned and tested of SHHS.

| Pretraining data set |  |  |  |
| --- | --- | --- | --- |
| SSC | MESA | MROS | Bioserenity |
| 0.65 | 0.66 | 0.66 | 0.65 |

**Supplementary Table 3.** Cross-dataset generalization results (macro F1 score) for sleep staging models trained on one cohort and tested on others. Diagonal values represent within-dataset performance.

| Pretrain + Finetune | MESA (test) | MrOS (test) | Stanford (test) | Avg. transfer |
| --- | --- | --- | --- | --- |
| **MESA** | 0.64 | 0.55 | 0.32 | 0.44 |
| **MrOS** | 0.40 | 0.65 | 0.39 | 0.40 |
| **Stanford** | 0.30 | 0.38 | 0.54 | 0.34 |
| **Avg. test reception** | 0.35 | 0.47 | 0.36 | – |

**Supplementary Table 4.** Sleep staging results (macro F1 score) using multi-dataset pretraining and fine-tuning. All models were pretrained on MESA, MrOS, Stanford, and Bioserenity, and fine-tuned on MESA, MrOS, Stanford, and SHHS.

| Test dataset | MESA | MrOS | Stanford |
| --- | --- | --- | --- |
| **Macro F1** | 0.74 | 0.74 | 0.67 |

**Supplementary Table 5.**
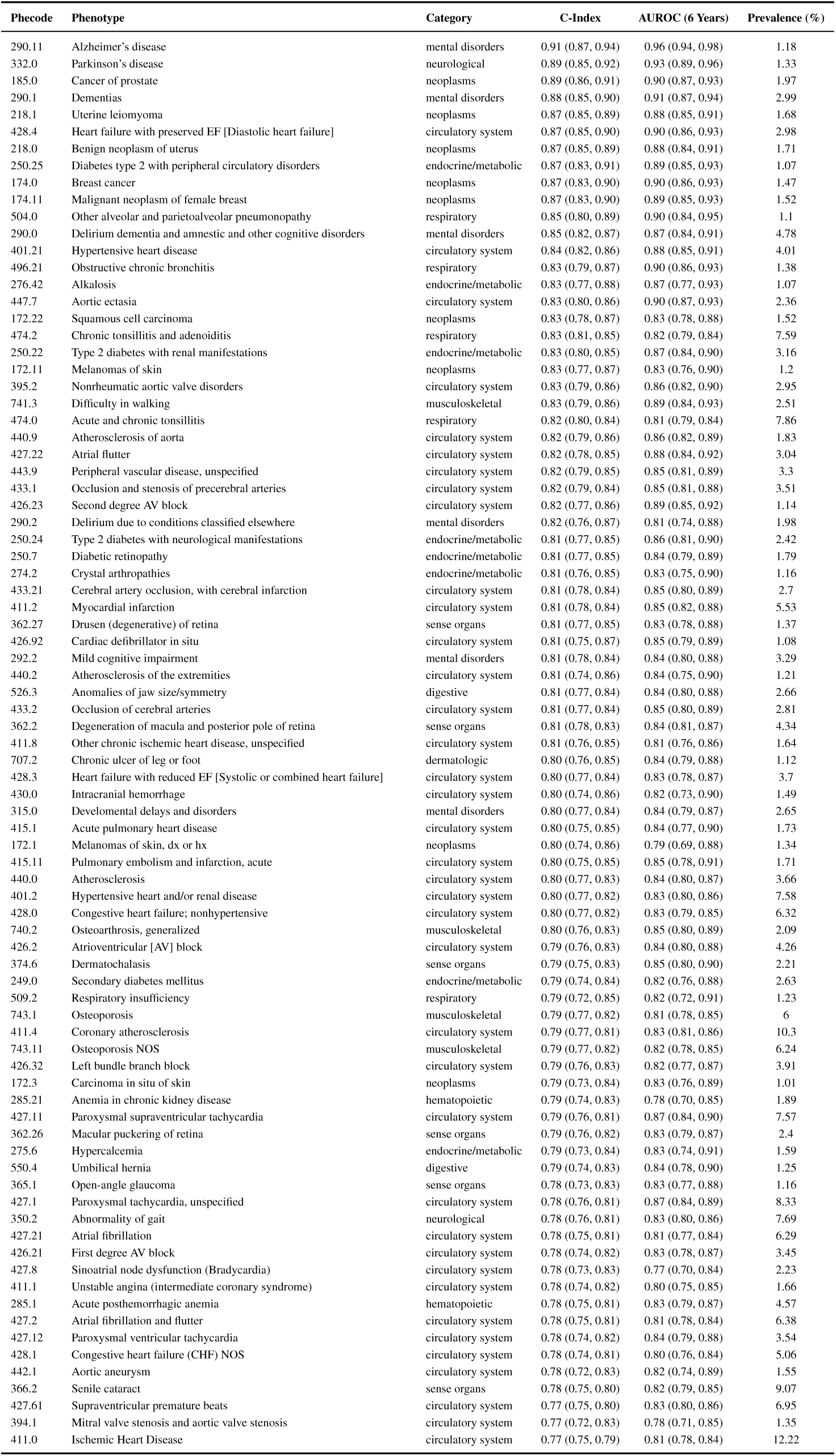
Summary of SleepFM performance metrics, including C-Index and AUROC (6 years) on the Stanford dataset. Results are sorted by C-Index, ensuring that both AUROC and C-Index exceed 0.75, with 95% confidence intervals that do not overlap with 0.5 (random chance). All conditions have statistically significant p-values (<0.01) after Bonferroni correction. Notable top conditions include Alzheimer’s disease, Parkinson’s disease, and dementias.

| Phecode | Phenotype | Category | C-Index | AUROC (6 Years) | Prevalence (%) |
| --- | --- | --- | --- | --- | --- |
| 290.11 | Alzheimer's disease | mental disorders | 0.91 (0.87, 0.94) | 0.96 (0.94, 0.98) | 1.18 |
| 332.0 | Parkinson's disease | neurological | 0.89 (0.85, 0.92) | 0.93 (0.89, 0.96) | 1.33 |
| 185.0 | Cancer of prostate | neoplasms | 0.89 (0.86, 0.91) | 0.90 (0.87, 0.93) | 1.97 |
| 290.1 | Dementias | mental disorders | 0.88 (0.85, 0.90) | 0.91 (0.87, 0.94) | 2.99 |
| 218.1 | Uterine leiomyoma | neoplasms | 0.87 (0.85, 0.89) | 0.88 (0.85, 0.91) | 1.68 |
| 428.4 | Heart failure with preserved EF [Diastolic heart failure] | circulatory system | 0.87 (0.85, 0.90) | 0.90 (0.86, 0.93) | 2.98 |
| 218.0 | Benign neoplasm of uterus | neoplasms | 0.87 (0.85, 0.89) | 0.88 (0.84, 0.91) | 1.71 |
| 250.25 | Diabetes type 2 with peripheral circulatory disorders | endocrine/metabolic | 0.87 (0.83, 0.91) | 0.89 (0.85, 0.93) | 1.07 |
| 174.0 | Breast cancer | neoplasms | 0.87 (0.83, 0.90) | 0.90 (0.86, 0.93) | 1.47 |
| 174.11 | Malignant neoplasm of female breast | neoplasms | 0.87 (0.83, 0.90) | 0.89 (0.85, 0.93) | 1.52 |
| 504.0 | Other alveolar and parietoalveolar pneumonopathy | respiratory | 0.85 (0.80, 0.89) | 0.90 (0.84, 0.95) | 1.1 |
| 290.0 | Delirium dementia and amnesic and other cognitive disorders | mental disorders | 0.85 (0.82, 0.87) | 0.87 (0.84, 0.91) | 4.78 |
| 401.21 | Hypertensive heart disease | circulatory system | 0.84 (0.82, 0.86) | 0.88 (0.85, 0.91) | 4.01 |
| 496.21 | Obstructive chronic bronchitis | respiratory | 0.83 (0.79, 0.87) | 0.90 (0.86, 0.93) | 1.38 |
| 276.42 | Alkalosis | endocrine/metabolic | 0.83 (0.77, 0.88) | 0.87 (0.77, 0.93) | 1.07 |
| 447.7 | Aortic ectasia | circulatory system | 0.83 (0.80, 0.86) | 0.90 (0.87, 0.93) | 2.36 |
| 172.22 | Squamous cell carcinoma | neoplasms | 0.83 (0.78, 0.87) | 0.83 (0.78, 0.88) | 1.52 |
| 474.2 | Chronic tonsillitis and adenoiditis | respiratory | 0.83 (0.81, 0.85) | 0.82 (0.79, 0.84) | 7.59 |
| 250.22 | Type 2 diabetes with renal manifestations | endocrine/metabolic | 0.83 (0.80, 0.85) | 0.87 (0.84, 0.90) | 3.16 |
| 172.11 | Melanomas of skin | neoplasms | 0.83 (0.77, 0.87) | 0.83 (0.76, 0.90) | 1.2 |
| 395.2 | Nonrheumatic aortic valve disorders | circulatory system | 0.83 (0.79, 0.86) | 0.86 (0.82, 0.90) | 2.95 |
| 741.3 | Difficulty in walking | musculoskeletal | 0.83 (0.79, 0.86) | 0.89 (0.84, 0.93) | 2.51 |
| 474.0 | Acute and chronic tonsillitis | respiratory | 0.82 (0.80, 0.84) | 0.81 (0.79, 0.84) | 7.86 |
| 440.9 | Atherosclerosis of aorta | circulatory system | 0.82 (0.79, 0.86) | 0.86 (0.82, 0.89) | 1.83 |
| 427.22 | Atrial flutter | circulatory system | 0.82 (0.78, 0.85) | 0.88 (0.84, 0.92) | 3.04 |
| 443.9 | Peripheral vascular disease, unspecified | circulatory system | 0.82 (0.79, 0.85) | 0.85 (0.81, 0.89) | 3.3 |
| 433.1 | Occlusion and stenosis of precerebral arteries | circulatory system | 0.82 (0.79, 0.84) | 0.85 (0.81, 0.88) | 3.51 |
| 426.23 | Second degree AV block | circulatory system | 0.82 (0.77, 0.86) | 0.89 (0.85, 0.92) | 1.14 |
| 290.2 | Delirium due to conditions classified elsewhere | mental disorders | 0.82 (0.76, 0.87) | 0.81 (0.74, 0.88) | 1.98 |
| 250.24 | Type 2 diabetes with neurological manifestations | endocrine/metabolic | 0.81 (0.77, 0.85) | 0.86 (0.81, 0.90) | 2.42 |
| 250.7 | Diabetic retinopathy | endocrine/metabolic | 0.81 (0.77, 0.85) | 0.84 (0.79, 0.89) | 1.79 |
| 274.2 | Crystal arthropathies | endocrine/metabolic | 0.81 (0.76, 0.85) | 0.83 (0.75, 0.90) | 1.16 |
| 433.21 | Cerebral artery occlusion, with cerebral infarction | circulatory system | 0.81 (0.78, 0.84) | 0.85 (0.80, 0.89) | 2.7 |
| 411.2 | Myocardial infarction | circulatory system | 0.81 (0.78, 0.84) | 0.85 (0.82, 0.88) | 5.53 |
| 362.27 | Drusen (degenerative) of retina | sense organs | 0.81 (0.77, 0.85) | 0.83 (0.78, 0.88) | 1.37 |
| 426.92 | Cardiac defibrillator in situ | circulatory system | 0.81 (0.75, 0.87) | 0.85 (0.79, 0.89) | 1.08 |
| 292.2 | Mild cognitive impairment | mental disorders | 0.81 (0.78, 0.84) | 0.84 (0.80, 0.88) | 3.29 |
| 440.2 | Atherosclerosis of the extremities | circulatory system | 0.81 (0.74, 0.86) | 0.84 (0.75, 0.90) | 1.21 |
| 526.3 | Anomalies of jaw size/symmetry | digestive | 0.81 (0.77, 0.84) | 0.84 (0.80, 0.88) | 2.66 |
| 433.2 | Occlusion of cerebral arteries | circulatory system | 0.81 (0.77, 0.84) | 0.85 (0.80, 0.89) | 2.81 |
| 362.2 | Degeneration of macula and posterior pole of retina | sense organs | 0.81 (0.78, 0.83) | 0.84 (0.81, 0.87) | 4.34 |
| 411.8 | Other chronic ischemic heart disease, unspecified | circulatory system | 0.81 (0.76, 0.85) | 0.81 (0.76, 0.86) | 1.64 |
| 707.2 | Chronic ulcer of leg or foot | dermatologic | 0.80 (0.76, 0.85) | 0.84 (0.79, 0.88) | 1.12 |
| 428.3 | Heart failure with reduced EF [Systolic or combined heart failure] | circulatory system | 0.80 (0.77, 0.84) | 0.83 (0.78, 0.87) | 3.7 |
| 430.0 | Intracranial hemorrhage | circulatory system | 0.80 (0.74, 0.86) | 0.82 (0.73, 0.90) | 1.49 |
| 315.0 | Developmental delays and disorders | mental disorders | 0.80 (0.77, 0.84) | 0.84 (0.79, 0.87) | 2.65 |
| 415.1 | Acute pulmonary heart disease | circulatory system | 0.80 (0.75, 0.85) | 0.84 (0.77, 0.90) | 1.73 |
| 172.1 | Melanomas of skin, dx or hx | neoplasms | 0.80 (0.74, 0.86) | 0.79 (0.69, 0.88) | 1.34 |
| 415.11 | Pulmonary embolism and infarction, acute | circulatory system | 0.80 (0.75, 0.85) | 0.85 (0.78, 0.91) | 1.71 |
| 440.0 | Atherosclerosis | circulatory system | 0.80 (0.77, 0.83) | 0.84 (0.80, 0.87) | 3.66 |
| 401.2 | Hypertensive heart and/or renal disease | circulatory system | 0.80 (0.77, 0.82) | 0.83 (0.80, 0.86) | 7.58 |
| 428.0 | Congestive heart failure; nonhypertensive | circulatory system | 0.80 (0.77, 0.82) | 0.83 (0.79, 0.85) | 6.32 |
| 740.2 | Osteoarthritis, generalized | musculoskeletal | 0.80 (0.76, 0.83) | 0.85 (0.80, 0.89) | 2.09 |
| 426.2 | Atrioventricular [AV] block | circulatory system | 0.79 (0.76, 0.83) | 0.84 (0.80, 0.88) | 4.26 |
| 374.6 | Dermatochalasis | sense organs | 0.79 (0.75, 0.83) | 0.85 (0.80, 0.90) | 2.21 |
| 249.0 | Secondary diabetes mellitus | endocrine/metabolic | 0.79 (0.74, 0.84) | 0.82 (0.76, 0.88) | 2.63 |
| 509.2 | Respiratory insufficiency | respiratory | 0.79 (0.72, 0.85) | 0.82 (0.72, 0.91) | 1.23 |
| 743.1 | Osteoporosis | musculoskeletal | 0.79 (0.77, 0.82) | 0.81 (0.78, 0.85) | 6 |
| 411.4 | Coronary atherosclerosis | circulatory system | 0.79 (0.77, 0.81) | 0.83 (0.81, 0.86) | 10.3 |
| 743.11 | Osteoporosis NOS | musculoskeletal | 0.79 (0.77, 0.82) | 0.82 (0.78, 0.85) | 6.24 |
| 426.32 | Left bundle branch block | circulatory system | 0.79 (0.76, 0.83) | 0.82 (0.77, 0.87) | 3.91 |
| 172.3 | Carcinoma in situ of skin | neoplasms | 0.79 (0.73, 0.84) | 0.83 (0.76, 0.89) | 1.01 |
| 285.21 | Anemia in chronic kidney disease | hematopoietic | 0.79 (0.74, 0.83) | 0.78 (0.70, 0.85) | 1.89 |
| 427.11 | Paroxysmal supraventricular tachycardia | circulatory system | 0.79 (0.76, 0.81) | 0.87 (0.84, 0.90) | 7.57 |
| 362.26 | Macular puckering of retina | sense organs | 0.79 (0.76, 0.82) | 0.83 (0.79, 0.87) | 2.4 |
| 275.6 | Hypercalcemia | endocrine/metabolic | 0.79 (0.73, 0.84) | 0.83 (0.74, 0.91) | 1.59 |
| 550.4 | Umbilical hernia | digestive | 0.79 (0.74, 0.83) | 0.84 (0.78, 0.90) | 1.25 |
| 365.1 | Open-angle glaucoma | sense organs | 0.78 (0.73, 0.83) | 0.83 (0.77, 0.88) | 1.16 |
| 427.1 | Paroxysmal tachycardia, unspecified | circulatory system | 0.78 (0.76, 0.81) | 0.87 (0.84, 0.89) | 8.33 |
| 350.2 | Abnormality of gait | neurological | 0.78 (0.76, 0.81) | 0.83 (0.80, 0.86) | 7.69 |
| 427.21 | Atrial fibrillation | circulatory system | 0.78 (0.75, 0.81) | 0.81 (0.77, 0.84) | 6.29 |
| 426.21 | First degree AV block | circulatory system | 0.78 (0.74, 0.82) | 0.83 (0.78, 0.87) | 3.45 |
| 427.8 | Sinoatrial node dysfunction (Bradycardia) | circulatory system | 0.78 (0.73, 0.83) | 0.77 (0.70, 0.84) | 2.23 |
| 411.1 | Unstable angina (intermediate coronary syndrome) | circulatory system | 0.78 (0.74, 0.82) | 0.80 (0.75, 0.85) | 1.66 |
| 285.1 | Acute posthemorrhagic anemia | hematopoietic | 0.78 (0.75, 0.81) | 0.83 (0.79, 0.87) | 4.57 |
| 427.2 | Atrial fibrillation and flutter | circulatory system | 0.78 (0.75, 0.81) | 0.81 (0.78, 0.84) | 6.38 |
| 427.12 | Paroxysmal ventricular tachycardia | circulatory system | 0.78 (0.74, 0.82) | 0.84 (0.79, 0.88) | 3.54 |
| 428.1 | Congestive heart failure (CHF) NOS | circulatory system | 0.78 (0.74, 0.81) | 0.80 (0.76, 0.84) | 5.06 |
| 442.1 | Aortic aneurysm | circulatory system | 0.78 (0.72, 0.83) | 0.82 (0.74, 0.89) | 1.55 |
| 366.2 | Senile cataract | sense organs | 0.78 (0.75, 0.80) | 0.82 (0.79, 0.85) | 9.07 |
| 427.61 | Supraventricular premature beats | circulatory system | 0.77 (0.75, 0.80) | 0.83 (0.80, 0.86) | 6.95 |
| 394.1 | Mitral valve stenosis and aortic valve stenosis | circulatory system | 0.77 (0.72, 0.83) | 0.78 (0.71, 0.85) | 1.35 |
| 411.0 | Ischemic Heart Disease | circulatory system | 0.77 (0.75, 0.79) | 0.81 (0.78, 0.84) | 12.22 |

**Supplementary Table 6.** Hand-selected diseases and their corresponding phecode groupings. These diseases were chosen for their relevance to sleep, in consultation with a sleep expert. All phecode groupings were reviewed and curated by a medical professional.

| Disease | Phecode | Phenotype |
| --- | --- | --- |
| Dementia | 290.0 | Delirium dementia and amnestic and other cognitive disorders |
| Dementia | 290.1 | Dementias |
| Dementia | 290.16 | Vascular dementia |
| Dementia | 290.12 | Dementia with cerebral degenerations |
| Dementia | 290.11 | Alzheimer's disease |
| Dementia | 290.13 | Senile dementia |
| Myocardial Infarction | 411.0 | Ischemic Heart Disease |
| Myocardial Infarction | 411.2 | Myocardial infarction |
| Ischemic Heart Disease | 411.0 | Ischemic Heart Disease |
| Ischemic Heart Disease | 411.8 | Other chronic ischemic heart disease, unspecified |
| Ischemic Heart Disease | 411.9 | Other acute and subacute forms of ischemic heart disease |
| Angina | 411.1 | Unstable angina (intermediate coronary syndrome) |
| Angina | 411.3 | Angina pectoris |
| Hypertension | 401.0 | Hypertension |
| Hypertension | 401.1 | Essential hypertension |
| Hypertension | 401.2 | Hypertensive heart and/or renal disease |
| Hypertension | 401.21 | Hypertensive heart disease |
| Hypertension | 401.3 | Other hypertensive complications |
| Hypertension | 401.22 | Hypertensive chronic kidney disease |
| Hypotension | 458.0 | Hypotension |
| Hypotension | 458.1 | Orthostatic hypotension |
| Hypotension | 458.2 | Iatrogenic hypotension |
| Hypotension | 458.9 | Hypotension NOS |
| Pulmonary Heart Disease | 415.0 | Pulmonary heart disease |
| Pulmonary Heart Disease | 415.2 | Chronic pulmonary heart disease |
| Pulmonary Heart Disease | 415.1 | Acute pulmonary heart disease |
| Pulmonary Heart Disease | 415.21 | Primary pulmonary hypertension |
| Pulmonary Heart Disease | 415.11 | Pulmonary embolism and infarction, acute |
| Atrial Fibrillation and Flutter | 427.0 | Cardiac dysrhythmias |
| Atrial Fibrillation and Flutter | 427.2 | Atrial fibrillation and flutter |
| Atrial Fibrillation and Flutter | 427.21 | Atrial fibrillation |
| Atrial Fibrillation and Flutter | 427.22 | Atrial flutter |
| Respiratory Failure | 509.0 | Respiratory failure, insufficiency, arrest |
| Respiratory Failure | 509.1 | Respiratory failure |
| Respiratory Failure | 509.2 | Respiratory insufficiency |
| Respiratory Failure | 509.3 | Pulmonary insufficiency or respiratory failure following trauma and surgery |
| Respiratory Failure | 509.5 | Respiratory arrest |
| Respiratory Failure | 509.8 | Dependence on respirator [Ventilator] or supplemental oxygen |

**Supplementary Table 7.** Performance comparison of models trained on different sleep stages across conditions. For each sleep stage, C-Index values are reported with 95% confidence intervals in parentheses. Only conditions where at least one sleep stage achieves a C-Index above 0.75 are included. Results are grouped by disease category and sorted alphabetically within each category. Stage 1/2 demonstrates the highest predictive performance across 39 conditions, followed by REM sleep with 15 conditions.

| Phecode | Phenotype | Category | Wake | Stage1/2 | Stage3 | REM | Best |
| --- | --- | --- | --- | --- | --- | --- | --- |
| 290.12 | Dementia With Cerebral Degenerations | Mental Disorders | 0.88 (0.82, 0.93) | 0.87 (0.78, 0.93) | 0.80 (0.68, 0.90) | 0.82 (0.69, 0.91) | Wake |
| 440.1 | Atherosclerosis Of Renal Artery | Circulatory System | 0.78 (0.65, 0.88) | 0.76 (0.53, 0.94) | 0.73 (0.50, 0.89) | 0.74 (0.55, 0.90) | Wake |
| 427.22 | Atrial Flutter | Circulatory System | 0.77 (0.73, 0.81) | 0.77 (0.72, 0.81) | 0.75 (0.70, 0.79) | 0.75 (0.72, 0.79) | Wake |
| 290.1 | Dementias | Mental Disorders | 0.77 (0.73, 0.80) | 0.75 (0.71, 0.79) | 0.75 (0.71, 0.79) | 0.75 (0.71, 0.80) | Wake |
| 306.1 | Mental Disorders Durring/After Pregnancy | Mental Disorders | 0.77 (0.68, 0.86) | 0.72 (0.53, 0.86) | 0.70 (0.51, 0.87) | 0.67 (0.48, 0.83) | Wake |
| 474.0 | Acute And Chronic Tonsillitis | Respiratory | 0.76 (0.74, 0.79) | 0.76 (0.73, 0.79) | 0.75 (0.72, 0.78) | 0.76 (0.74, 0.79) | Wake |
| 440.21 | Atherosclerosis Of Native Arteries Of The Extremities With Ulceration Or Gangrene | Circulatory System | 0.82 (0.63, 0.95) | 0.95 (0.89, 1.00) | 0.83 (0.74, 0.93) | 0.83 (0.71, 0.95) | Stage 1/2 |
| 290.13 | Senile Dementia | Mental Disorders | 0.90 (0.85, 0.97) | 0.94 (0.89, 0.99) | 0.66 (0.39, 1.00) | 0.89 (0.85, 0.94) | Stage 1/2 |
| 264.9 | Lack Of Normal Physiological Development, Unspecified | Endocrine/Metabolic | 0.87 (0.78, 0.95) | 0.91 (0.84, 0.96) | 0.91 (0.84, 0.96) | 0.86 (0.77, 0.94) | Stage 1/2 |
| 313.3 | Autism | Mental Disorders | 0.88 (0.82, 0.93) | 0.88 (0.83, 0.93) | 0.88 (0.81, 0.93) | 0.87 (0.79, 0.92) | Stage 1/2 |
| 411.8 | Other Chronic Ischemic Heart Disease, Unspecified | Circulatory System | 0.77 (0.73, 0.82) | 0.83 (0.79, 0.86) | 0.73 (0.69, 0.77) | 0.78 (0.72, 0.82) | Stage 1/2 |
| 426.24 | Atrioventricular Block, Complete | Circulatory System | 0.82 (0.76, 0.87) | 0.82 (0.75, 0.89) | 0.80 (0.74, 0.87) | 0.82 (0.76, 0.87) | Stage 1/2 |
| 250.25 | Diabetes Type 2 With Peripheral Circulatory Disorders | Endocrine/Metabolic | 0.81 (0.76, 0.87) | 0.82 (0.76, 0.88) | 0.75 (0.69, 0.80) | 0.81 (0.76, 0.86) | Stage 1/2 |
| 496.21 | Obstructive Chronic Bronchitis | Respiratory | 0.74 (0.68, 0.79) | 0.82 (0.78, 0.86) | 0.71 (0.65, 0.76) | 0.78 (0.73, 0.83) | Stage 1/2 |
| 264.0 | Lack Of Normal Physiological Development | Endocrine/Metabolic | 0.78 (0.68, 0.87) | 0.81 (0.72, 0.88) | 0.78 (0.67, 0.87) | 0.79 (0.70, 0.87) | Stage 1/2 |
| 426.92 | Cardiac Defibrillator In Situ | Circulatory System | 0.76 (0.68, 0.82) | 0.79 (0.72, 0.87) | 0.79 (0.74, 0.85) | 0.74 (0.67, 0.81) | Stage 1/2 |
| 276.42 | Alkalosis | Endocrine/Metabolic | 0.74 (0.65, 0.82) | 0.79 (0.71, 0.85) | 0.72 (0.63, 0.80) | 0.77 (0.70, 0.83) | Stage 1/2 |
| 250.22 | Type 2 Diabetes With Renal Manifestations | Endocrine/Metabolic | 0.72 (0.68, 0.76) | 0.79 (0.75, 0.82) | 0.75 (0.70, 0.79) | 0.77 (0.73, 0.81) | Stage 1/2 |
| 331.0 | Other Cerebral Degenerations | Neurological | 0.74 (0.65, 0.81) | 0.79 (0.72, 0.85) | 0.75 (0.67, 0.81) | 0.74 (0.67, 0.80) | Stage 1/2 |
| 427.4 | Cardiac Arrest And Ventricular Fibrillation | Circulatory System | 0.69 (0.60, 0.77) | 0.78 (0.70, 0.85) | 0.68 (0.58, 0.78) | 0.66 (0.58, 0.75) | Stage 1/2 |
| 256.4 | Polycystic Ovaries | Endocrine/Metabolic | 0.76 (0.66, 0.84) | 0.78 (0.70, 0.85) | 0.73 (0.66, 0.79) | 0.63 (0.54, 0.73) | Stage 1/2 |
| 250.1 | Type 1 Diabetes | Endocrine/Metabolic | 0.69 (0.61, 0.76) | 0.78 (0.72, 0.83) | 0.64 (0.56, 0.71) | 0.71 (0.65, 0.77) | Stage 1/2 |
| 440.9 | Atherosclerosis Of Aorta | Circulatory System | 0.73 (0.68, 0.79) | 0.77 (0.72, 0.81) | 0.69 (0.63, 0.75) | 0.75 (0.70, 0.80) | Stage 1/2 |
| 707.2 | Chronic Ulcer Of Leg Or Foot | Dermatologic | 0.72 (0.64, 0.79) | 0.77 (0.71, 0.82) | 0.69 (0.63, 0.75) | 0.71 (0.63, 0.77) | Stage 1/2 |
| 707.1 | Decubitus Ulcer | Dermatologic | 0.74 (0.67, 0.81) | 0.77 (0.72, 0.82) | 0.69 (0.61, 0.76) | 0.75 (0.68, 0.81) | Stage 1/2 |
| 256.0 | Ovarian Dysfunction | Endocrine/Metabolic | 0.74 (0.65, 0.81) | 0.77 (0.69, 0.84) | 0.71 (0.64, 0.77) | 0.63 (0.54, 0.71) | Stage 1/2 |
| 287.32 | Secondary Thrombocytopenia | Hematopoietic | 0.75 (0.67, 0.82) | 0.77 (0.68, 0.84) | 0.68 (0.57, 0.77) | 0.72 (0.62, 0.80) | Stage 1/2 |
| 426.9 | Cardiac Pacemaker/Device In Situ | Circulatory System | 0.72 (0.68, 0.76) | 0.76 (0.72, 0.81) | 0.75 (0.71, 0.80) | 0.71 (0.67, 0.75) | Stage 1/2 |
| 426.21 | First Degree Av Block | Circulatory System | 0.75 (0.71, 0.79) | 0.76 (0.72, 0.80) | 0.69 (0.64, 0.74) | 0.74 (0.69, 0.79) | Stage 1/2 |
| 428.3 | Heart Failure With Reduced Ef [Systolic Or Combined Heart Failure] | Circulatory System | 0.74 (0.71, 0.78) | 0.76 (0.73, 0.80) | 0.71 (0.67, 0.75) | 0.73 (0.70, 0.77) | Stage 1/2 |
| 430.0 | Intracranial Hemorrhage | Circulatory System | 0.73 (0.65, 0.80) | 0.76 (0.70, 0.81) | 0.70 (0.63, 0.77) | 0.74 (0.67, 0.81) | Stage 1/2 |
| 433.8 | Late Effects Of Cerebrovascular Disease | Circulatory System | 0.74 (0.66, 0.80) | 0.76 (0.69, 0.81) | 0.70 (0.62, 0.76) | 0.69 (0.61, 0.76) | Stage 1/2 |
| 443.9 | Peripheral Vascular Disease, Unspecified | Circulatory System | 0.75 (0.71, 0.78) | 0.76 (0.72, 0.80) | 0.69 (0.65, 0.73) | 0.76 (0.72, 0.79) | Stage 1/2 |
| 415.11 | Pulmonary Embolism And Infarction, Acute | Circulatory System | 0.72 (0.66, 0.77) | 0.76 (0.70, 0.81) | 0.74 (0.68, 0.79) | 0.70 (0.63, 0.77) | Stage 1/2 |
| 250.24 | Type 2 Diabetes With Neurological Manifestations | Endocrine/Metabolic | 0.72 (0.66, 0.77) | 0.76 (0.71, 0.81) | 0.68 (0.63, 0.72) | 0.75 (0.71, 0.79) | Stage 1/2 |
| 172.11 | Melanomas Of Skin | Neoplasms | 0.68 (0.60, 0.75) | 0.76 (0.70, 0.82) | 0.75 (0.67, 0.82) | 0.70 (0.63, 0.77) | Stage 1/2 |
| 332.0 | Parkinson'S Disease | Neurological | 0.76 (0.69, 0.81) | 0.76 (0.70, 0.81) | 0.73 (0.67, 0.78) | 0.75 (0.68, 0.82) | Stage 1/2 |
| 377.1 | Optic Atrophy | Sense Organs | 0.70 (0.64, 0.77) | 0.76 (0.70, 0.82) | 0.66 (0.59, 0.73) | 0.69 (0.63, 0.77) | Stage 1/2 |
| 415.1 | Acute Pulmonary Heart Disease | Circulatory System | 0.72 (0.66, 0.77) | 0.75 (0.69, 0.81) | 0.74 (0.68, 0.80) | 0.70 (0.63, 0.76) | Stage 1/2 |
| 440.0 | Atherosclerosis | Circulatory System | 0.72 (0.68, 0.75) | 0.75 (0.71, 0.79) | 0.69 (0.65, 0.73) | 0.71 (0.67, 0.75) | Stage 1/2 |
| 428.0 | Congestive Heart Failure; Nonhypertensive | Circulatory System | 0.72 (0.69, 0.75) | 0.75 (0.72, 0.78) | 0.72 (0.69, 0.75) | 0.73 (0.70, 0.76) | Stage 1/2 |
| 457.2 | Encounter For Long-Term (Current) Use Of Antiplatelet-s/Antithrombotics | Circulatory System | 0.71 (0.66, 0.76) | 0.75 (0.72, 0.79) | 0.71 (0.67, 0.76) | 0.72 (0.67, 0.77) | Stage 1/2 |
| 426.32 | Left Bundle Branch Block | Circulatory System | 0.74 (0.71, 0.78) | 0.75 (0.71, 0.79) | 0.72 (0.68, 0.76) | 0.74 (0.70, 0.78) | Stage 1/2 |
| 509.1 | Respiratory Failure | Respiratory | 0.70 (0.66, 0.74) | 0.75 (0.71, 0.79) | 0.68 (0.64, 0.73) | 0.72 (0.67, 0.76) | Stage 1/2 |
| 509.2 | Respiratory Insufficiency | Respiratory | 0.71 (0.64, 0.77) | 0.75 (0.70, 0.81) | 0.71 (0.63, 0.78) | 0.69 (0.61, 0.76) | Stage 1/2 |
| 426.91 | Cardiac Pacemaker In Situ | Circulatory System | 0.74 (0.69, 0.78) | 0.78 (0.73, 0.82) | 0.79 (0.74, 0.83) | 0.73 (0.68, 0.77) | Stage 3 |
| 425.2 | Secondary/Extrinsic Cardiomyopathies | Circulatory System | 0.59 (0.49, 0.69) | 0.60 (0.51, 0.68) | 0.77 (0.66, 0.86) | 0.58 (0.48, 0.69) | Stage 3 |
| 428.4 | Heart Failure With Preserved Ef [Diastolic Heart Failure] | Circulatory System | 0.80 (0.76, 0.83) | 0.81 (0.78, 0.85) | 0.76 (0.73, 0.80) | 0.82 (0.78, 0.85) | REM |
| 290.16 | Vascular Dementia | Mental Disorders | 0.80 (0.72, 0.87) | 0.76 (0.67, 0.84) | 0.77 (0.67, 0.86) | 0.80 (0.71, 0.87) | REM |
| 362.29 | Macular Degeneration (Senile) Of Retina Nos | Sense Organs | 0.77 (0.72, 0.83) | 0.78 (0.72, 0.83) | 0.70 (0.62, 0.77) | 0.79 (0.74, 0.84) | REM |
| 315.2 | Speech And Language Disorder | Mental Disorders | 0.77 (0.67, 0.85) | 0.74 (0.65, 0.82) | 0.77 (0.68, 0.84) | 0.78 (0.68, 0.85) | REM |
| 401.21 | Hypertensive Heart Disease | Circulatory System | 0.72 (0.68, 0.76) | 0.74 (0.71, 0.78) | 0.74 (0.70, 0.77) | 0.77 (0.73, 0.80) | REM |
| 411.9 | Other Acute And Subacute Forms Of Ischemic Heart Disease | Circulatory System | 0.71 (0.62, 0.79) | 0.69 (0.59, 0.79) | 0.67 (0.58, 0.77) | 0.77 (0.69, 0.85) | REM |
| 426.8 | Other Cardiac Conduction Disorders | Circulatory System | 0.76 (0.72, 0.79) | 0.75 (0.71, 0.79) | 0.69 (0.65, 0.73) | 0.77 (0.74, 0.81) | REM |
| 290.0 | Delirium Dementia And Amnesic And Other Cognitive Disorders | Mental Disorders | 0.75 (0.72, 0.79) | 0.74 (0.70, 0.77) | 0.73 (0.70, 0.76) | 0.77 (0.73, 0.80) | REM |
| 290.2 | Delirium Due To Conditions Classified Elsewhere | Mental Disorders | 0.75 (0.70, 0.80) | 0.73 (0.67, 0.79) | 0.69 (0.63, 0.75) | 0.77 (0.72, 0.82) | REM |
| 474.2 | Chronic Tonsillitis And Adenoiditis | Respiratory | 0.77 (0.74, 0.79) | 0.77 (0.74, 0.79) | 0.75 (0.72, 0.78) | 0.77 (0.74, 0.79) | REM |
| 250.7 | Diabetic Retinopathy | Endocrine/Metabolic | 0.74 (0.67, 0.79) | 0.73 (0.68, 0.78) | 0.67 (0.61, 0.73) | 0.76 (0.71, 0.81) | REM |
| 264.2 | Failure To Thrive (Childhood) | Endocrine/Metabolic | 0.74 (0.62, 0.86) | 0.73 (0.56, 0.86) | 0.66 (0.51, 0.83) | 0.75 (0.66, 0.86) | REM |
| 260.2 | Severe Protein-Calorie Malnutrition | Endocrine/Metabolic | 0.73 (0.67, 0.78) | 0.70 (0.64, 0.75) | 0.62 (0.55, 0.68) | 0.75 (0.70, 0.80) | REM |
| 333.1 | Essential Tremor | Neurological | 0.69 (0.63, 0.76) | 0.70 (0.62, 0.77) | 0.65 (0.58, 0.71) | 0.75 (0.68, 0.82) | REM |
| 504.0 | Other Alveolar And Parietoalveolar Pneumonopathy | Respiratory | 0.72 (0.65, 0.79) | 0.73 (0.65, 0.80) | 0.75 (0.67, 0.83) | 0.75 (0.67, 0.83) | REM |

**Supplementary Table 8.**
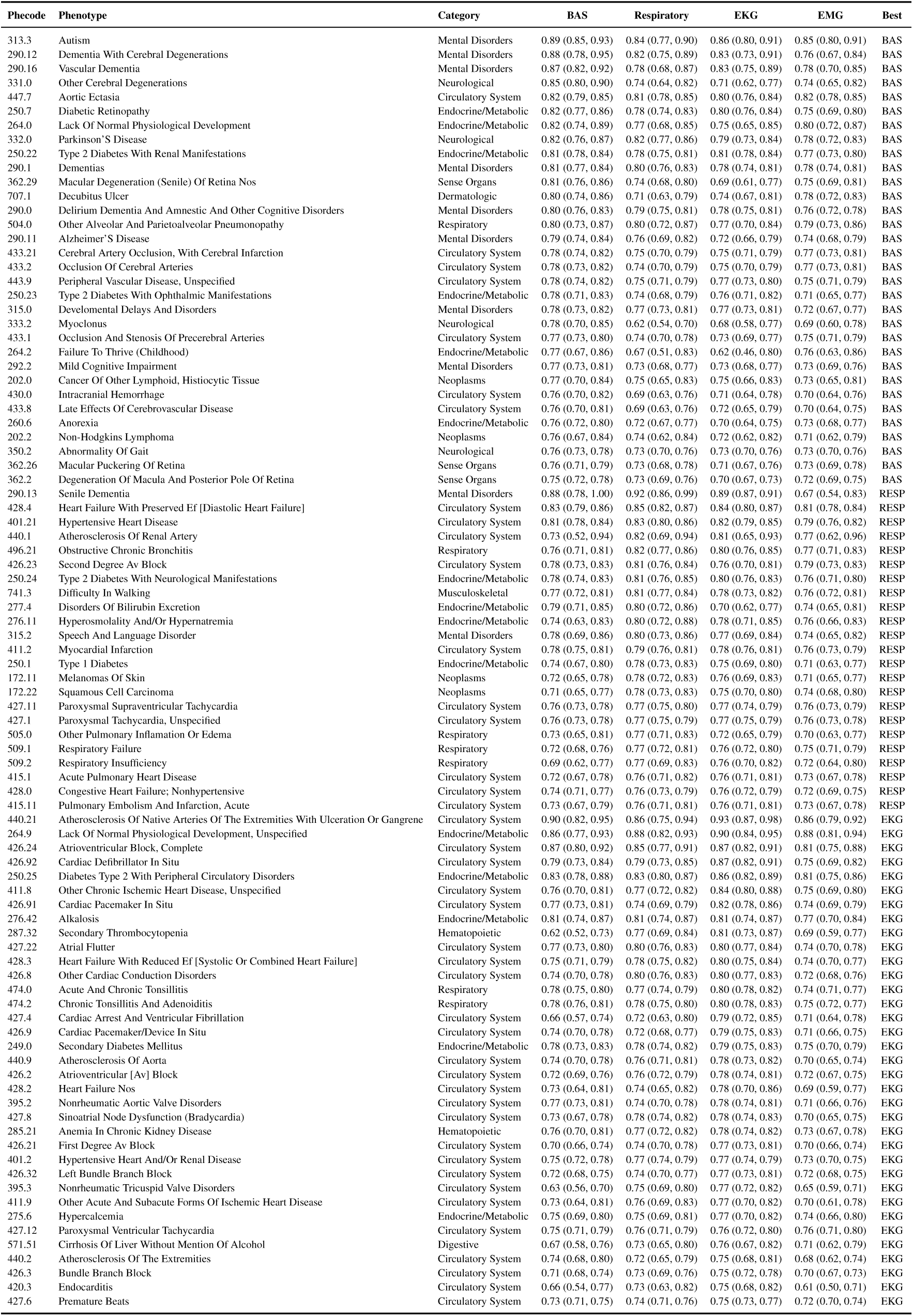
Performance comparison of different modalities across conditions. For each modality, C-Index values are reported with 95% confidence intervals in parentheses. Only conditions where at least one modality achieves a C-Index above 0.75 are included. Results are grouped by disease category and sorted alphabetically within each category. BAS and EKG demonstrate the highest predictive performance, followed closely by RESP. EMG is the least predictive, with no top conditions where it performs best.

**Supplementary Table 9.** Performance comparison between SleepFM trained on all modalities versus models trained on individual modalities alone. The table shows average C-Index across disease categories for each configuration. While certain modalities perform relatively well on their own, combining all modalities yields the highest performance, demonstrating the complementary nature of multimodal signals in PSG. These results support the benefit of leveraging multimodal data during training and highlight how different physiological signals contribute to disease prediction.

| Category | BAS | EKG | EMG | RESP | SleepFM |
| --- | --- | --- | --- | --- | --- |
| circulatory system | 0.71 | 0.73 | 0.69 | 0.72 | 0.75 |
| dermatologic | 0.63 | 0.63 | 0.63 | 0.63 | 0.66 |
| digestive | 0.63 | 0.65 | 0.64 | 0.65 | 0.67 |
| endocrine/metabolic | 0.67 | 0.68 | 0.66 | 0.68 | 0.71 |
| hematopoietic | 0.67 | 0.69 | 0.66 | 0.69 | 0.70 |
| infectious diseases | 0.63 | 0.63 | 0.63 | 0.64 | 0.66 |
| injuries & poisonings | 0.61 | 0.64 | 0.62 | 0.64 | 0.66 |
| mental disorders | 0.66 | 0.66 | 0.65 | 0.66 | 0.70 |
| musculoskeletal | 0.64 | 0.64 | 0.64 | 0.64 | 0.68 |
| neoplasms | 0.67 | 0.68 | 0.66 | 0.67 | 0.73 |
| neurological | 0.65 | 0.65 | 0.64 | 0.64 | 0.67 |
| respiratory | 0.62 | 0.64 | 0.62 | 0.64 | 0.66 |
| sense organs | 0.65 | 0.64 | 0.63 | 0.64 | 0.68 |
| symptoms | 0.66 | 0.66 | 0.66 | 0.66 | 0.68 |

**Supplementary Table 10.**
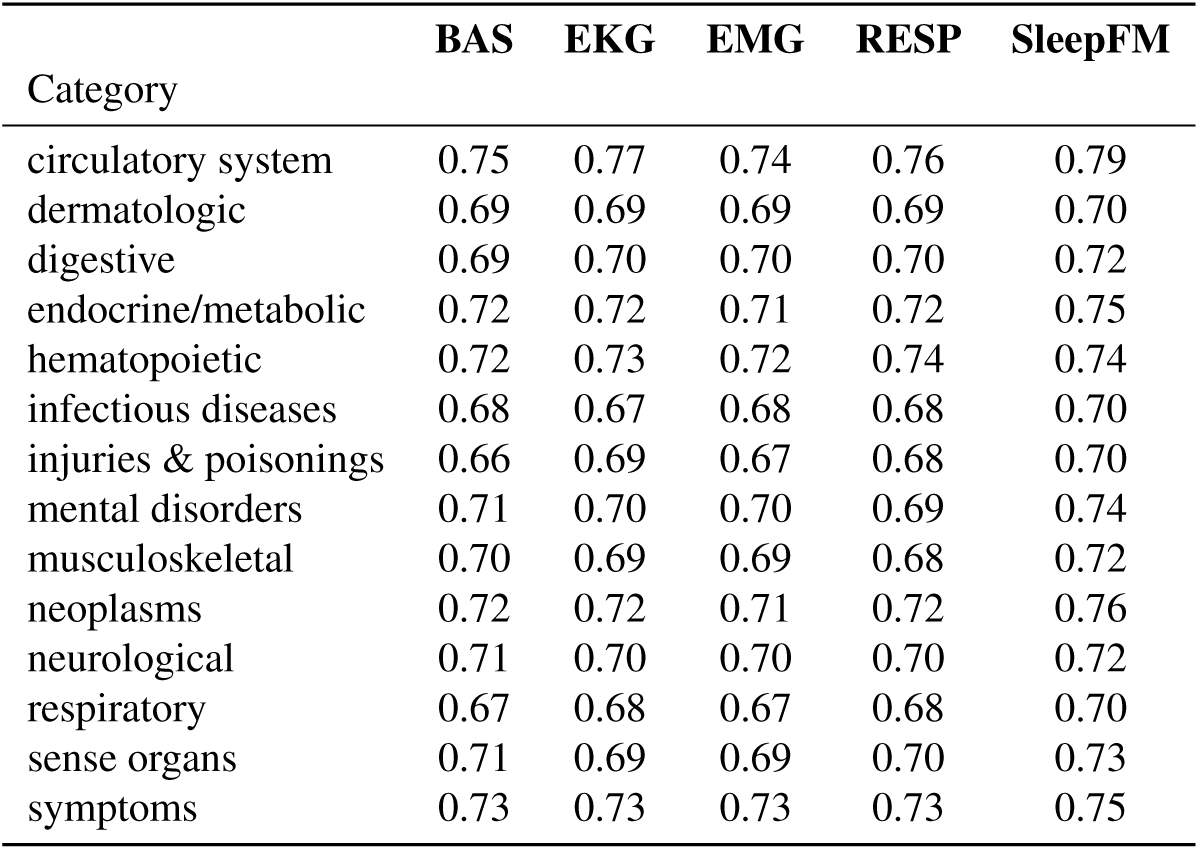
Performance comparison between SleepFM trained on all modalities versus models trained on individual modalities alone. The table shows average AUROC across disease categories for each configuration. While certain modalities perform relatively well on their own, combining all modalities yields the highest performance, demonstrating the complementary nature of multimodal signals in PSG. These results support the benefit of leveraging multimodal data during training and highlight how different physiological signals contribute to disease prediction.

**Supplementary Table 11.** Top conditions where sleep recordings provide substantial predictive value beyond demographic information alone. For each condition, we compare the predictive performance (C-Index) of SleepFM against demographic-only and end-to-end PSG models. The table highlights conditions where SleepFM achieved both strong absolute performance (C-Index > 0.75) and demonstrated meaningful improvement over demographic features (difference in C-Index > 0.5). Conditions are ranked by their relative improvement over demographics, emphasizing areas where sleep patterns contribute most significantly to prediction accuracy. Notable top conditions where SleepFM performs much better than baseline, include dementia, respiratory insufficiency, and developmental delays and disorders.

| Phcode | Phenotype | Category | SleepFM | Demographics | End-to-End PSG |
| --- | --- | --- | --- | --- | --- |
| 290.13 | Senile dementia | mental disorders | 0.99 (0.98, 1.00) | 0.87 (0.75, 0.96) | 0.95 (0.91, 0.98) |
| 440.21 | Atherosclerosis of native arteries of the extremities with ulceration or gangrene | circulatory system | 0.92 (0.88, 0.95) | 0.74 (0.64, 0.89) | 0.66 (0.50, 0.89) |
| 358.0 | Myoneural disorders | neurological | 0.81 (0.73, 0.88) | 0.42 (0.28, 0.55) | 0.60 (0.48, 0.71) |
| 264.2 | Failure to thrive (childhood) | endocrine/metabolic | 0.77 (0.68, 0.88) | 0.44 (0.26, 0.67) | 0.45 (0.26, 0.67) |
| 315.0 | Developmental delays and disorders | mental disorders | 0.80 (0.77, 0.84) | 0.58 (0.51, 0.64) | 0.61 (0.54, 0.67) |
| 509.2 | Respiratory insufficiency | respiratory | 0.79 (0.72, 0.85) | 0.59 (0.51, 0.67) | 0.62 (0.53, 0.69) |
| 277.4 | Disorders of bilirubin excretion | endocrine/metabolic | 0.79 (0.70, 0.85) | 0.60 (0.46, 0.75) | 0.61 (0.47, 0.75) |
| 344.0 | Other paralytic syndromes | neurological | 0.77 (0.72, 0.83) | 0.58 (0.48, 0.69) | 0.60 (0.48, 0.69) |
| 626.8 | Infertility, female | genitourinary | 0.89 (0.84, 0.93) | 0.80 (0.73, 0.87) | 0.88 (0.83, 0.92) |
| 614.51 | Cervicitis and endocervicitis | genitourinary | 0.88 (0.85, 0.91) | 0.79 (0.74, 0.83) | 0.86 (0.83, 0.89) |
| 619.2 | Disorders of uterus, NEC | genitourinary | 0.87 (0.84, 0.90) | 0.79 (0.76, 0.82) | 0.84 (0.81, 0.88) |
| 609.0 | Male infertility and abnormal spermatozoa | genitourinary | 0.82 (0.78, 0.86) | 0.71 (0.66, 0.76) | 0.82 (0.78, 0.86) |
| 504.0 | Other alveolar and parietoalveolar pneumonopathy | respiratory | 0.85 (0.80, 0.89) | 0.75 (0.68, 0.81) | 0.75 (0.68, 0.81) |
| 619.3 | Noninflammatory disorders of cervix | genitourinary | 0.86 (0.82, 0.89) | 0.77 (0.72, 0.81) | 0.82 (0.79, 0.85) |
| 250.25 | Diabetes type 2 with peripheral circulatory disorders | endocrine/metabolic | 0.87 (0.83, 0.91) | 0.79 (0.74, 0.85) | 0.78 (0.72, 0.84) |
| 426.24 | Atrioventricular block, complete | circulatory system | 0.87 (0.83, 0.92) | 0.80 (0.73, 0.87) | 0.83 (0.76, 0.89) |
| 348.8 | Encephalopathy, not elsewhere classified | neurological | 0.77 (0.70, 0.83) | 0.63 (0.56, 0.71) | 0.71 (0.64, 0.77) |
| 276.42 | Alkalosis | endocrine/metabolic | 0.83 (0.77, 0.88) | 0.74 (0.66, 0.80) | 0.71 (0.63, 0.78) |
| 249.0 | Secondary diabetes mellitus | endocrine/metabolic | 0.79 (0.74, 0.84) | 0.68 (0.63, 0.72) | 0.67 (0.62, 0.71) |
| 447.7 | Aortic ectasia | circulatory system | 0.83 (0.80, 0.86) | 0.74 (0.69, 0.79) | 0.78 (0.74, 0.82) |
| 280.2 | Iron deficiency anemia secondary to blood loss (chronic) | hematopoietic | 0.77 (0.73, 0.81) | 0.64 (0.59, 0.69) | 0.64 (0.60, 0.69) |
| 218.1 | Uterine leiomyoma | neoplasms | 0.87 (0.85, 0.89) | 0.81 (0.79, 0.83) | 0.87 (0.84, 0.89) |
| 300.9 | Posttraumatic stress disorder | mental disorders | 0.75 (0.70, 0.79) | 0.62 (0.56, 0.68) | 0.64 (0.59, 0.69) |
| 331.0 | Other cerebral degenerations | neurological | 0.83 (0.76, 0.88) | 0.74 (0.65, 0.82) | 0.79 (0.70, 0.87) |
| 290.2 | Delirium due to conditions classified elsewhere | mental disorders | 0.82 (0.76, 0.87) | 0.73 (0.67, 0.78) | 0.77 (0.71, 0.82) |
| 218.0 | Benign neoplasm of uterus | neoplasms | 0.87 (0.85, 0.89) | 0.81 (0.79, 0.83) | 0.87 (0.85, 0.89) |
| 276.4 | Acid-base balance disorder | endocrine/metabolic | 0.77 (0.72, 0.82) | 0.66 (0.61, 0.72) | 0.67 (0.61, 0.72) |
| 250.24 | Type 2 diabetes with neurological manifestations | endocrine/metabolic | 0.81 (0.77, 0.85) | 0.73 (0.68, 0.77) | 0.71 (0.68, 0.75) |
| 315.2 | Speech and language disorder | mental disorders | 0.81 (0.74, 0.87) | 0.73 (0.63, 0.82) | 0.73 (0.64, 0.82) |
| 530.2 | Esophageal bleeding (varices/hemorrhage) | digestive | 0.76 (0.72, 0.79) | 0.65 (0.60, 0.71) | 0.69 (0.64, 0.74) |
| 260.6 | Anorexia | endocrine/metabolic | 0.76 (0.71, 0.80) | 0.65 (0.59, 0.72) | 0.68 (0.62, 0.74) |
| 458.2 | Iatrogenic hypotension | circulatory system | 0.76 (0.68, 0.83) | 0.66 (0.58, 0.74) | 0.67 (0.60, 0.74) |
| 585.31 | Renal dialysis | genitourinary | 0.78 (0.73, 0.82) | 0.68 (0.63, 0.74) | 0.66 (0.60, 0.71) |
| 426.8 | Other cardiac conduction disorders | circulatory system | 0.79 (0.75, 0.82) | 0.70 (0.65, 0.74) | 0.71 (0.66, 0.75) |
| 276.11 | Hyperosmolality and/or hypernatremia | endocrine/metabolic | 0.76 (0.66, 0.86) | 0.67 (0.58, 0.75) | 0.68 (0.59, 0.76) |
| 264.0 | Lack of normal physiological development | endocrine/metabolic | 0.77 (0.67, 0.86) | 0.69 (0.56, 0.81) | 0.69 (0.55, 0.81) |
| 707.2 | Chronic ulcer of leg or foot | dermatologic | 0.80 (0.76, 0.85) | 0.73 (0.68, 0.79) | 0.70 (0.65, 0.75) |
| 622.1 | Polyp of corpus uteri | genitourinary | 0.85 (0.81, 0.89) | 0.79 (0.75, 0.83) | 0.85 (0.81, 0.88) |
| 594.3 | Calculus of ureter | genitourinary | 0.75 (0.69, 0.81) | 0.67 (0.60, 0.73) | 0.67 (0.62, 0.73) |
| 250.7 | Diabetic retinopathy | endocrine/metabolic | 0.81 (0.77, 0.85) | 0.75 (0.69, 0.80) | 0.74 (0.69, 0.79) |
| 513.8 | Disorders of diaphragm | respiratory | 0.75 (0.65, 0.84) | 0.67 (0.56, 0.76) | 0.68 (0.57, 0.77) |
| 592.11 | Acute cystitis | genitourinary | 0.81 (0.78, 0.83) | 0.74 (0.72, 0.77) | 0.74 (0.71, 0.77) |
| 285.1 | Acute posthemorrhagic anemia | hematopoietic | 0.78 (0.75, 0.81) | 0.71 (0.67, 0.74) | 0.72 (0.68, 0.75) |
| 440.2 | Atherosclerosis of the extremities | circulatory system | 0.81 (0.74, 0.86) | 0.75 (0.68, 0.80) | 0.75 (0.69, 0.81) |
| 362.27 | Drusen (degenerative) of retina | sense organs | 0.81 (0.77, 0.85) | 0.75 (0.69, 0.80) | 0.83 (0.79, 0.87) |
| 501.0 | Pneumonitis due to inhalation of food or vomitus | respiratory | 0.75 (0.70, 0.80) | 0.68 (0.62, 0.73) | 0.69 (0.62, 0.76) |
| 415.11 | Pulmonary embolism and infarction, acute | circulatory system | 0.80 (0.75, 0.85) | 0.74 (0.68, 0.79) | 0.75 (0.69, 0.81) |
| 427.11 | Paroxysmal supraventricular tachycardia | circulatory system | 0.79 (0.76, 0.81) | 0.72 (0.69, 0.76) | 0.73 (0.70, 0.76) |
| 415.1 | Acute pulmonary heart disease | circulatory system | 0.80 (0.75, 0.85) | 0.74 (0.68, 0.80) | 0.75 (0.69, 0.81) |
| 411.8 | Other chronic ischemic heart disease, unspecified | circulatory system | 0.81 (0.76, 0.85) | 0.75 (0.69, 0.79) | 0.77 (0.72, 0.81) |
| 440.22 | Atherosclerosis of native arteries of the extremities with intermittent claudication | circulatory system | 0.80 (0.73, 0.86) | 0.75 (0.68, 0.83) | 0.74 (0.65, 0.81) |
| 411.9 | Other acute and subacute forms of ischemic heart disease | circulatory system | 0.78 (0.72, 0.85) | 0.72 (0.64, 0.79) | 0.72 (0.66, 0.79) |
| 509.1 | Respiratory failure | respiratory | 0.77 (0.73, 0.80) | 0.70 (0.65, 0.74) | 0.69 (0.64, 0.73) |
| 740.2 | Osteoarthritis, generalized | musculoskeletal | 0.80 (0.76, 0.83) | 0.74 (0.69, 0.79) | 0.79 (0.74, 0.82) |

**Supplementary Table 12.** Top conditions where sleep recordings demonstrate strong predictive performance for 6-year risk assessment. For each condition, we compare the 6-year AUROC of SleepFM against demographic-only and end-to-end PSG models. The table includes conditions where SleepFM achieved both strong absolute performance (AUROC > 0.75) and substantial improvement over the demographic baseline. Conditions are ranked by their relative improvement over demographics, highlighting diseases where sleep patterns provide the most significant additional predictive value. Notable top conditions where SleepFM performs much better than baseline, include dementia, respiratory insufficiency, and developmental delays and disorders.

| Phencode | Phenotype | Category | SleepFM | Demographics | End-to-End PSG |
| --- | --- | --- | --- | --- | --- |
| 290.13 | Senile dementia | mental disorders | 0.99 (0.98, 1.00) | 0.87 (0.75, 0.96) | 0.95 (0.91, 0.98) |
| 440.21 | Atherosclerosis of native arteries of the extremities with ulceration or gangrene | circulatory system | 0.92 (0.88, 0.95) | 0.74 (0.64, 0.89) | 0.66 (0.50, 0.89) |
| 358.0 | Myoneural disorders | neurological | 0.81 (0.73, 0.88) | 0.42 (0.28, 0.55) | 0.60 (0.48, 0.71) |
| 264.2 | Failure to thrive (childhood) | endocrine/metabolic | 0.77 (0.68, 0.88) | 0.44 (0.26, 0.67) | 0.45 (0.26, 0.67) |
| 315.0 | Developmental delays and disorders | mental disorders | 0.80 (0.77, 0.84) | 0.58 (0.51, 0.64) | 0.61 (0.54, 0.67) |
| 509.2 | Respiratory insufficiency | respiratory | 0.79 (0.72, 0.85) | 0.59 (0.51, 0.67) | 0.62 (0.53, 0.69) |
| 277.4 | Disorders of bilirubin excretion | endocrine/metabolic | 0.79 (0.70, 0.85) | 0.60 (0.46, 0.75) | 0.61 (0.47, 0.75) |
| 344.0 | Other paralytic syndromes | neurological | 0.77 (0.72, 0.83) | 0.58 (0.48, 0.69) | 0.60 (0.48, 0.69) |
| 504.0 | Other alveolar and parietoalveolar pneumonopathy | respiratory | 0.85 (0.80, 0.89) | 0.75 (0.68, 0.81) | 0.75 (0.68, 0.81) |
| 250.25 | Diabetes type 2 with peripheral circulatory disorders | endocrine/metabolic | 0.87 (0.83, 0.91) | 0.79 (0.74, 0.85) | 0.78 (0.72, 0.84) |
| 426.24 | Atrioventricular block, complete | circulatory system | 0.87 (0.83, 0.92) | 0.80 (0.73, 0.87) | 0.83 (0.76, 0.89) |
| 348.8 | Encephalopathy, not elsewhere classified | neurological | 0.77 (0.70, 0.83) | 0.63 (0.56, 0.71) | 0.71 (0.64, 0.77) |
| 276.42 | Alkalosis | endocrine/metabolic | 0.83 (0.77, 0.88) | 0.74 (0.66, 0.80) | 0.71 (0.63, 0.78) |
| 249.0 | Secondary diabetes mellitus | endocrine/metabolic | 0.79 (0.74, 0.84) | 0.68 (0.63, 0.72) | 0.67 (0.62, 0.71) |
| 447.7 | Aortic ectasia | circulatory system | 0.83 (0.80, 0.86) | 0.74 (0.69, 0.79) | 0.78 (0.74, 0.82) |
| 280.2 | Iron deficiency anemia secondary to blood loss (chronic) | hematopoietic | 0.77 (0.73, 0.81) | 0.64 (0.59, 0.69) | 0.64 (0.60, 0.69) |
| 218.1 | Uterine leiomyoma | neoplasms | 0.87 (0.85, 0.89) | 0.81 (0.79, 0.83) | 0.87 (0.84, 0.89) |
| 300.9 | Posttraumatic stress disorder | mental disorders | 0.75 (0.70, 0.79) | 0.62 (0.56, 0.68) | 0.64 (0.59, 0.69) |
| 331.0 | Other cerebral degenerations | neurological | 0.83 (0.76, 0.88) | 0.74 (0.65, 0.82) | 0.79 (0.70, 0.87) |
| 290.2 | Delirium due to conditions classified elsewhere | mental disorders | 0.82 (0.76, 0.87) | 0.73 (0.67, 0.78) | 0.77 (0.71, 0.82) |
| 218.0 | Benign neoplasm of uterus | neoplasms | 0.87 (0.85, 0.89) | 0.81 (0.79, 0.83) | 0.87 (0.85, 0.89) |
| 276.4 | Acid-base balance disorder | endocrine/metabolic | 0.77 (0.72, 0.82) | 0.66 (0.61, 0.72) | 0.67 (0.61, 0.72) |
| 250.24 | Type 2 diabetes with neurological manifestations | endocrine/metabolic | 0.81 (0.77, 0.85) | 0.73 (0.68, 0.77) | 0.71 (0.68, 0.75) |
| 315.2 | Speech and language disorder | mental disorders | 0.81 (0.74, 0.87) | 0.73 (0.63, 0.82) | 0.73 (0.64, 0.82) |
| 530.2 | Esophageal bleeding (varices/hemorrhage) | digestive | 0.76 (0.72, 0.79) | 0.65 (0.60, 0.71) | 0.69 (0.64, 0.74) |
| 260.6 | Anorexia | endocrine/metabolic | 0.76 (0.71, 0.80) | 0.65 (0.59, 0.72) | 0.68 (0.62, 0.74) |
| 458.2 | Iatrogenic hypotension | circulatory system | 0.76 (0.68, 0.83) | 0.66 (0.58, 0.74) | 0.67 (0.60, 0.74) |
| 426.8 | Other cardiac conduction disorders | circulatory system | 0.79 (0.75, 0.82) | 0.70 (0.65, 0.74) | 0.71 (0.66, 0.75) |
| 276.11 | Hyperosmolality and/or hypernatremia | endocrine/metabolic | 0.76 (0.66, 0.86) | 0.67 (0.58, 0.75) | 0.68 (0.59, 0.76) |
| 264.0 | Lack of normal physiological development | endocrine/metabolic | 0.77 (0.67, 0.86) | 0.69 (0.56, 0.81) | 0.69 (0.55, 0.81) |
| 707.2 | Chronic ulcer of leg or foot | dermatologic | 0.80 (0.76, 0.85) | 0.73 (0.68, 0.79) | 0.70 (0.65, 0.75) |
| 250.7 | Diabetic retinopathy | endocrine/metabolic | 0.81 (0.77, 0.85) | 0.75 (0.69, 0.80) | 0.74 (0.69, 0.79) |
| 513.8 | Disorders of diaphragm | respiratory | 0.75 (0.65, 0.84) | 0.67 (0.56, 0.76) | 0.68 (0.57, 0.77) |
| 285.1 | Acute posthemorrhagic anemia | hematopoietic | 0.78 (0.75, 0.81) | 0.71 (0.67, 0.74) | 0.72 (0.68, 0.75) |
| 440.2 | Atherosclerosis of the extremities | circulatory system | 0.81 (0.74, 0.86) | 0.75 (0.68, 0.80) | 0.75 (0.69, 0.81) |
| 362.27 | Drusen (degenerative) of retina | sense organs | 0.81 (0.77, 0.85) | 0.75 (0.69, 0.80) | 0.83 (0.79, 0.87) |
| 501.0 | Pneumonitis due to inhalation of food or vomitus | respiratory | 0.75 (0.70, 0.80) | 0.68 (0.62, 0.73) | 0.69 (0.62, 0.76) |
| 415.11 | Pulmonary embolism and infarction, acute | circulatory system | 0.80 (0.75, 0.85) | 0.74 (0.68, 0.79) | 0.75 (0.69, 0.81) |
| 427.11 | Paroxysmal supraventricular tachycardia | circulatory system | 0.79 (0.76, 0.81) | 0.72 (0.69, 0.76) | 0.73 (0.70, 0.76) |
| 415.1 | Acute pulmonary heart disease | circulatory system | 0.80 (0.75, 0.85) | 0.74 (0.68, 0.80) | 0.75 (0.69, 0.81) |
| 411.8 | Other chronic ischemic heart disease, unspecified | circulatory system | 0.81 (0.76, 0.85) | 0.75 (0.69, 0.79) | 0.77 (0.72, 0.81) |
| 440.22 | Atherosclerosis of native arteries of the extremities with intermittent claudication | circulatory system | 0.80 (0.73, 0.86) | 0.75 (0.68, 0.83) | 0.74 (0.65, 0.81) |
| 411.9 | Other acute and subacute forms of ischemic heart disease | circulatory system | 0.78 (0.72, 0.85) | 0.72 (0.64, 0.79) | 0.72 (0.66, 0.79) |
| 509.1 | Respiratory failure | respiratory | 0.77 (0.73, 0.80) | 0.70 (0.65, 0.74) | 0.69 (0.64, 0.73) |
| 740.2 | Osteoarthritis, generalized | musculoskeletal | 0.80 (0.76, 0.83) | 0.74 (0.69, 0.79) | 0.79 (0.74, 0.82) |
| 427.1 | Paroxysmal tachycardia, unspecified | circulatory system | 0.78 (0.76, 0.81) | 0.73 (0.70, 0.75) | 0.73 (0.71, 0.76) |
| 427.4 | Cardiac arrest and ventricular fibrillation | circulatory system | 0.77 (0.68, 0.84) | 0.71 (0.63, 0.78) | 0.73 (0.65, 0.81) |
| 569.2 | Gastrointestinal complications | digestive | 0.76 (0.70, 0.81) | 0.69 (0.62, 0.77) | 0.66 (0.59, 0.72) |
| 427.12 | Paroxysmal ventricular tachycardia | circulatory system | 0.78 (0.74, 0.82) | 0.72 (0.68, 0.77) | 0.72 (0.67, 0.77) |
| 260.2 | severe protein-calorie malnutrition | endocrine/metabolic | 0.79 (0.74, 0.83) | 0.73 (0.68, 0.78) | 0.71 (0.67, 0.76) |
| 285.2 | Anemia of chronic disease | hematopoietic | 0.76 (0.72, 0.79) | 0.70 (0.66, 0.75) | 0.71 (0.67, 0.75) |
| 459.9 | Circulatory disease NEC | circulatory system | 0.77 (0.74, 0.79) | 0.71 (0.68, 0.74) | 0.71 (0.68, 0.75) |
| 276.13 | Hyperpotassemia | endocrine/metabolic | 0.75 (0.71, 0.80) | 0.70 (0.65, 0.75) | 0.70 (0.65, 0.74) |
| 574.2 | Calculus of bile duct | digestive | 0.76 (0.70, 0.80) | 0.70 (0.63, 0.78) | 0.70 (0.63, 0.78) |
| 250.1 | Type 1 diabetes | endocrine/metabolic | 0.75 (0.69, 0.81) | 0.70 (0.64, 0.75) | 0.68 (0.62, 0.75) |

**Supplementary Table 13.**
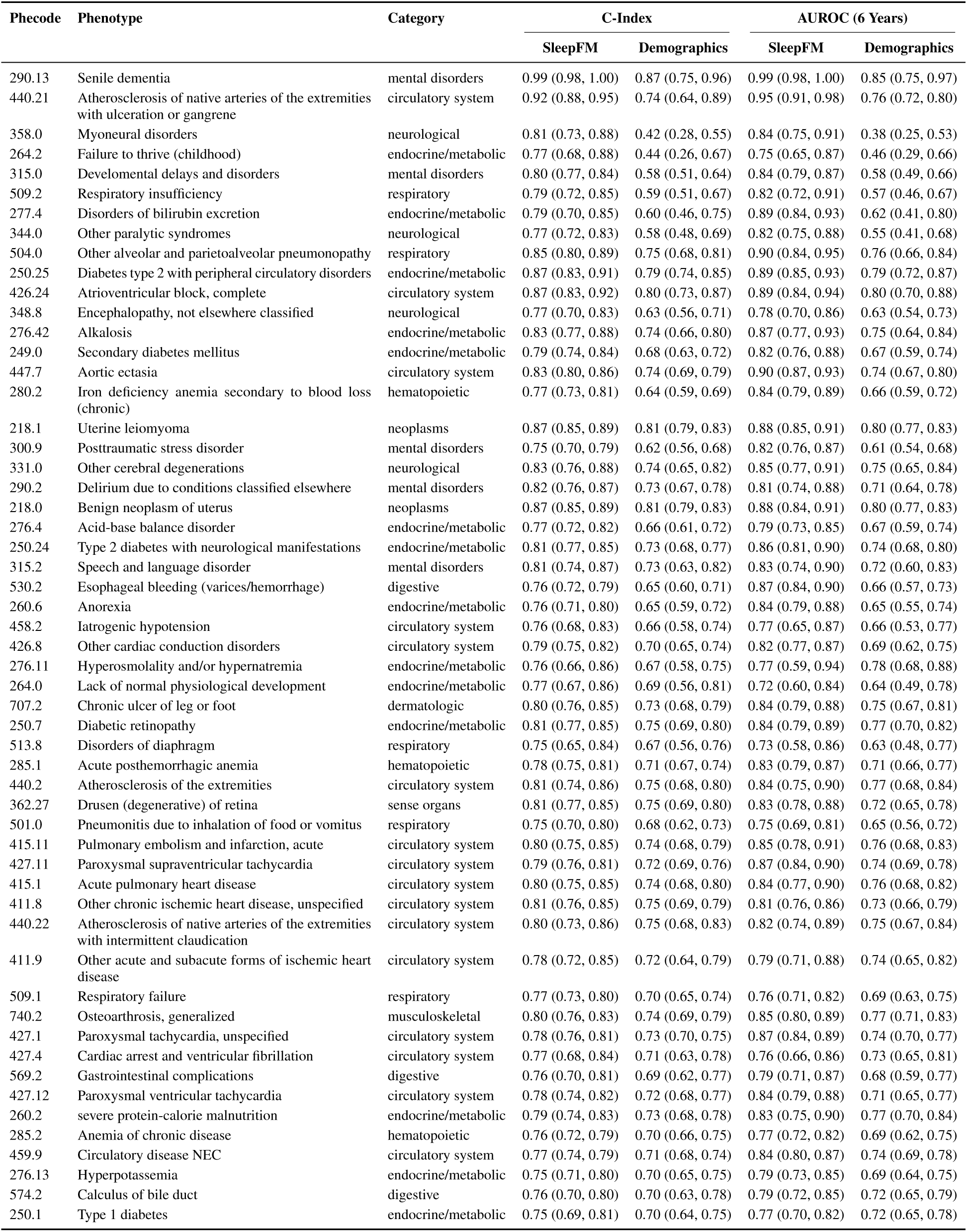
Comparison of performance metrics, including C-Index and AUROC (6 Years) for SleepFM and demographics model. Conditions are ranked by their relative improvement over demographics model, retaining only conditions where the C-Index for SleepFM exceeds 0.75. All metrics include 95% confidence intervals calculated using bootstrapping.

| Phcode | Phenotype | Category | C-Index |  | AUROC (6 Years) |  |
| --- | --- | --- | --- | --- | --- | --- |
|  |  |  | SleepFM | Demographics | SleepFM | Demographics |
| 290.13 | Senile dementia | mental disorders | 0.99 (0.98, 1.00) | 0.87 (0.75, 0.96) | 0.99 (0.98, 1.00) | 0.85 (0.75, 0.97) |
| 440.21 | Atherosclerosis of native arteries of the extremities with ulceration or gangrene | circulatory system | 0.92 (0.88, 0.95) | 0.74 (0.64, 0.89) | 0.95 (0.91, 0.98) | 0.76 (0.72, 0.80) |
| 358.0 | Myoneural disorders | neurological | 0.81 (0.73, 0.88) | 0.42 (0.28, 0.55) | 0.84 (0.75, 0.91) | 0.38 (0.25, 0.53) |
| 264.2 | Failure to thrive (childhood) | endocrine/metabolic | 0.77 (0.68, 0.88) | 0.44 (0.26, 0.67) | 0.75 (0.65, 0.87) | 0.46 (0.29, 0.66) |
| 315.0 | Develomental delays and disorders | mental disorders | 0.80 (0.77, 0.84) | 0.58 (0.51, 0.64) | 0.84 (0.79, 0.87) | 0.58 (0.49, 0.66) |
| 509.2 | Respiratory insufficiency | respiratory | 0.79 (0.72, 0.85) | 0.59 (0.51, 0.67) | 0.82 (0.72, 0.91) | 0.57 (0.46, 0.67) |
| 277.4 | Disorders of bilirubin excretion | endocrine/metabolic | 0.79 (0.70, 0.85) | 0.60 (0.46, 0.75) | 0.89 (0.84, 0.93) | 0.62 (0.41, 0.80) |
| 344.0 | Other paralytic syndromes | neurological | 0.77 (0.72, 0.83) | 0.58 (0.48, 0.69) | 0.82 (0.75, 0.88) | 0.55 (0.41, 0.68) |
| 504.0 | Other alveolar and parietoalveolar pneumonopathy | respiratory | 0.85 (0.80, 0.89) | 0.75 (0.68, 0.81) | 0.90 (0.84, 0.95) | 0.76 (0.66, 0.84) |
| 250.25 | Diabetes type 2 with peripheral circulatory disorders | endocrine/metabolic | 0.87 (0.83, 0.91) | 0.79 (0.74, 0.85) | 0.89 (0.85, 0.93) | 0.79 (0.72, 0.87) |
| 426.24 | Atrioventricular block, complete | circulatory system | 0.87 (0.83, 0.92) | 0.80 (0.73, 0.87) | 0.89 (0.84, 0.94) | 0.80 (0.70, 0.88) |
| 348.8 | Encephalopathy, not elsewhere classified | neurological | 0.77 (0.70, 0.83) | 0.63 (0.56, 0.71) | 0.78 (0.70, 0.86) | 0.63 (0.54, 0.73) |
| 276.42 | Alkalosis | endocrine/metabolic | 0.83 (0.77, 0.88) | 0.74 (0.66, 0.80) | 0.87 (0.77, 0.93) | 0.75 (0.64, 0.84) |
| 249.0 | Secondary diabetes mellitus | endocrine/metabolic | 0.79 (0.74, 0.84) | 0.68 (0.63, 0.72) | 0.82 (0.76, 0.88) | 0.67 (0.59, 0.74) |
| 447.7 | Aortic ectasia | circulatory system | 0.83 (0.80, 0.86) | 0.74 (0.69, 0.79) | 0.90 (0.87, 0.93) | 0.74 (0.67, 0.80) |
| 280.2 | Iron deficiency anemia secondary to blood loss (chronic) | hematopoietic | 0.77 (0.73, 0.81) | 0.64 (0.59, 0.69) | 0.84 (0.79, 0.89) | 0.66 (0.59, 0.72) |
| 218.1 | Uterine leiomyoma | neoplasms | 0.87 (0.85, 0.89) | 0.81 (0.79, 0.83) | 0.88 (0.85, 0.91) | 0.80 (0.77, 0.83) |
| 300.9 | Posttraumatic stress disorder | mental disorders | 0.75 (0.70, 0.79) | 0.62 (0.56, 0.68) | 0.82 (0.76, 0.87) | 0.61 (0.54, 0.68) |
| 331.0 | Other cerebral degenerations | neurological | 0.83 (0.76, 0.88) | 0.74 (0.65, 0.82) | 0.85 (0.77, 0.91) | 0.75 (0.65, 0.84) |
| 290.2 | Delirium due to conditions classified elsewhere | mental disorders | 0.82 (0.76, 0.87) | 0.73 (0.67, 0.78) | 0.81 (0.74, 0.88) | 0.71 (0.64, 0.78) |
| 218.0 | Benign neoplasm of uterus | neoplasms | 0.87 (0.85, 0.89) | 0.81 (0.79, 0.83) | 0.88 (0.84, 0.91) | 0.80 (0.77, 0.83) |
| 276.4 | Acid-base balance disorder | endocrine/metabolic | 0.77 (0.72, 0.82) | 0.66 (0.61, 0.72) | 0.79 (0.73, 0.85) | 0.67 (0.59, 0.74) |
| 250.24 | Type 2 diabetes with neurological manifestations | endocrine/metabolic | 0.81 (0.77, 0.85) | 0.73 (0.68, 0.77) | 0.86 (0.81, 0.90) | 0.74 (0.68, 0.80) |
| 315.2 | Speech and language disorder | mental disorders | 0.81 (0.74, 0.87) | 0.73 (0.63, 0.82) | 0.83 (0.74, 0.90) | 0.72 (0.60, 0.83) |
| 530.2 | Esophageal bleeding (varices/hemorrhage) | digestive | 0.76 (0.72, 0.79) | 0.65 (0.60, 0.71) | 0.87 (0.84, 0.90) | 0.66 (0.57, 0.73) |
| 260.6 | Anorexia | endocrine/metabolic | 0.76 (0.71, 0.80) | 0.65 (0.59, 0.72) | 0.84 (0.79, 0.88) | 0.65 (0.55, 0.74) |
| 458.2 | Iatrogenic hypotension | circulatory system | 0.76 (0.68, 0.83) | 0.66 (0.58, 0.74) | 0.77 (0.65, 0.87) | 0.66 (0.53, 0.77) |
| 426.8 | Other cardiac conduction disorders | circulatory system | 0.79 (0.75, 0.82) | 0.70 (0.65, 0.74) | 0.82 (0.77, 0.87) | 0.69 (0.62, 0.75) |
| 276.11 | Hyperosmolality and/or hypernatremia | endocrine/metabolic | 0.76 (0.66, 0.86) | 0.67 (0.58, 0.75) | 0.77 (0.59, 0.94) | 0.78 (0.68, 0.88) |
| 264.0 | Lack of normal physiological development | endocrine/metabolic | 0.77 (0.67, 0.86) | 0.69 (0.56, 0.81) | 0.72 (0.60, 0.84) | 0.64 (0.49, 0.78) |
| 707.2 | Chronic ulcer of leg or foot | dermatologic | 0.80 (0.76, 0.85) | 0.73 (0.68, 0.79) | 0.84 (0.79, 0.88) | 0.75 (0.67, 0.81) |
| 250.7 | Diabetic retinopathy | endocrine/metabolic | 0.81 (0.77, 0.85) | 0.75 (0.69, 0.80) | 0.84 (0.79, 0.89) | 0.77 (0.70, 0.82) |
| 513.8 | Disorders of diaphragm | respiratory | 0.75 (0.65, 0.84) | 0.67 (0.56, 0.76) | 0.73 (0.58, 0.86) | 0.63 (0.48, 0.77) |
| 285.1 | Acute posthemorrhagic anemia | hematopoietic | 0.78 (0.75, 0.81) | 0.71 (0.67, 0.74) | 0.83 (0.79, 0.87) | 0.71 (0.66, 0.77) |
| 440.2 | Atherosclerosis of the extremities | circulatory system | 0.81 (0.74, 0.86) | 0.75 (0.68, 0.80) | 0.84 (0.75, 0.90) | 0.77 (0.68, 0.84) |
| 362.27 | Drusen (degenerative) of retina | sense organs | 0.81 (0.77, 0.85) | 0.75 (0.69, 0.80) | 0.83 (0.78, 0.88) | 0.72 (0.65, 0.78) |
| 501.0 | Pneumonitis due to inhalation of food or vomitus | respiratory | 0.75 (0.70, 0.80) | 0.68 (0.62, 0.73) | 0.75 (0.69, 0.81) | 0.65 (0.56, 0.72) |
| 415.11 | Pulmonary embolism and infarction, acute | circulatory system | 0.80 (0.75, 0.85) | 0.74 (0.68, 0.79) | 0.85 (0.78, 0.91) | 0.76 (0.68, 0.83) |
| 427.11 | Paroxysmal supraventricular tachycardia | circulatory system | 0.79 (0.76, 0.81) | 0.72 (0.69, 0.76) | 0.87 (0.84, 0.90) | 0.74 (0.69, 0.78) |
| 415.1 | Acute pulmonary heart disease | circulatory system | 0.80 (0.75, 0.85) | 0.74 (0.68, 0.80) | 0.84 (0.77, 0.90) | 0.76 (0.68, 0.82) |
| 411.8 | Other chronic ischemic heart disease, unspecified | circulatory system | 0.81 (0.76, 0.85) | 0.75 (0.69, 0.79) | 0.81 (0.76, 0.86) | 0.73 (0.66, 0.79) |
| 440.22 | Atherosclerosis of native arteries of the extremities with intermittent claudication | circulatory system | 0.80 (0.73, 0.86) | 0.75 (0.68, 0.83) | 0.82 (0.74, 0.89) | 0.75 (0.67, 0.84) |
| 411.9 | Other acute and subacute forms of ischemic heart disease | circulatory system | 0.78 (0.72, 0.85) | 0.72 (0.64, 0.79) | 0.79 (0.71, 0.88) | 0.74 (0.65, 0.82) |
| 509.1 | Respiratory failure | respiratory | 0.77 (0.73, 0.80) | 0.70 (0.65, 0.74) | 0.76 (0.71, 0.82) | 0.69 (0.63, 0.75) |
| 740.2 | Osteoarthritis, generalized | musculoskeletal | 0.80 (0.76, 0.83) | 0.74 (0.69, 0.79) | 0.85 (0.80, 0.89) | 0.77 (0.71, 0.83) |
| 427.1 | Paroxysmal tachycardia, unspecified | circulatory system | 0.78 (0.76, 0.81) | 0.73 (0.70, 0.75) | 0.87 (0.84, 0.89) | 0.74 (0.70, 0.77) |
| 427.4 | Cardiac arrest and ventricular fibrillation | circulatory system | 0.77 (0.68, 0.84) | 0.71 (0.63, 0.78) | 0.76 (0.66, 0.86) | 0.73 (0.65, 0.81) |
| 569.2 | Gastrointestinal complications | digestive | 0.76 (0.70, 0.81) | 0.69 (0.62, 0.77) | 0.79 (0.71, 0.87) | 0.68 (0.59, 0.77) |
| 427.12 | Paroxysmal ventricular tachycardia | circulatory system | 0.78 (0.74, 0.82) | 0.72 (0.68, 0.77) | 0.84 (0.79, 0.88) | 0.71 (0.65, 0.77) |
| 260.2 | severe protein-calorie malnutrition | endocrine/metabolic | 0.79 (0.74, 0.83) | 0.73 (0.68, 0.78) | 0.83 (0.75, 0.90) | 0.77 (0.70, 0.84) |
| 285.2 | Anemia of chronic disease | hematopoietic | 0.76 (0.72, 0.79) | 0.70 (0.66, 0.75) | 0.77 (0.72, 0.82) | 0.69 (0.62, 0.75) |
| 459.9 | Circulatory disease NEC | circulatory system | 0.77 (0.74, 0.79) | 0.71 (0.68, 0.74) | 0.84 (0.80, 0.87) | 0.74 (0.69, 0.78) |
| 276.13 | Hyperpotassemia | endocrine/metabolic | 0.75 (0.71, 0.80) | 0.70 (0.65, 0.75) | 0.79 (0.73, 0.85) | 0.69 (0.64, 0.75) |
| 574.2 | Calculus of bile duct | digestive | 0.76 (0.70, 0.80) | 0.70 (0.63, 0.78) | 0.79 (0.72, 0.85) | 0.72 (0.65, 0.79) |
| 250.1 | Type 1 diabetes | endocrine/metabolic | 0.75 (0.69, 0.81) | 0.70 (0.64, 0.75) | 0.77 (0.70, 0.82) | 0.72 (0.65, 0.78) |

**Supplementary Table 14.** Comparison of performance metrics, including C-Index and AUROC (6 Years) for SleepFM and End-to-End PSG model. Conditions are ranked by their relative improvement over End-to-End PSG model, retaining only conditions where the C-Index for SleepFM exceeds 0.75. All metrics include 95% confidence intervals calculated using bootstrapping.

| Phecode | Phenotype | Category | C-Index |  | AUROC (6 Years) |  |
| --- | --- | --- | --- | --- | --- | --- |
|  |  |  | SleepFM | End-to-End PSG | SleepFM | End-to-End PSG |
| 440.21 | Atherosclerosis of native arteries of the extremities with ulceration or gangrene | circulatory system | 0.92 (0.88, 0.95) | 0.66 (0.50, 0.89) | 0.95 (0.92, 0.98) | 0.65 (0.61, 0.69) |
| 264.2 | Failure to thrive (childhood) | endocrine/metabolic | 0.77 (0.68, 0.88) | 0.45 (0.26, 0.67) | 0.75 (0.65, 0.87) | 0.44 (0.26, 0.66) |
| 358.0 | Myoneural disorders | neurological | 0.81 (0.73, 0.88) | 0.60 (0.48, 0.71) | 0.84 (0.75, 0.91) | 0.54 (0.40, 0.69) |
| 315.0 | Develomental delays and disorders | mental disorders | 0.80 (0.77, 0.84) | 0.61 (0.54, 0.67) | 0.84 (0.79, 0.87) | 0.61 (0.52, 0.69) |
| 509.2 | Respiratory insufficiency | respiratory | 0.79 (0.72, 0.85) | 0.62 (0.53, 0.69) | 0.82 (0.72, 0.91) | 0.64 (0.54, 0.73) |
| 277.4 | Disorders of bilirubin excretion | endocrine/metabolic | 0.79 (0.70, 0.85) | 0.61 (0.47, 0.75) | 0.89 (0.84, 0.93) | 0.60 (0.41, 0.78) |
| 344.0 | Other paralytic syndromes | neurological | 0.77 (0.72, 0.83) | 0.60 (0.48, 0.69) | 0.82 (0.75, 0.88) | 0.54 (0.40, 0.68) |
| 276.42 | Alkalosis | endocrine/metabolic | 0.83 (0.77, 0.88) | 0.71 (0.63, 0.78) | 0.87 (0.77, 0.93) | 0.71 (0.59, 0.81) |
| 250.25 | Diabetes type 2 with peripheral circulatory disorders | endocrine/metabolic | 0.87 (0.83, 0.91) | 0.78 (0.72, 0.84) | 0.89 (0.85, 0.93) | 0.79 (0.70, 0.87) |
| 504.0 | Other alveolar and parietoalveolar pneumonopathy | respiratory | 0.85 (0.80, 0.89) | 0.75 (0.68, 0.81) | 0.90 (0.84, 0.95) | 0.76 (0.66, 0.84) |
| 249.0 | Secondary diabetes mellitus | endocrine/metabolic | 0.79 (0.74, 0.84) | 0.67 (0.62, 0.71) | 0.82 (0.76, 0.88) | 0.66 (0.59, 0.72) |
| 250.24 | Type 2 diabetes with neurological manifestations | endocrine/metabolic | 0.81 (0.77, 0.85) | 0.71 (0.68, 0.75) | 0.86 (0.81, 0.90) | 0.73 (0.67, 0.78) |
| 280.2 | Iron deficiency anemia secondary to blood loss (chronic) | hematopoietic | 0.77 (0.73, 0.81) | 0.64 (0.60, 0.69) | 0.84 (0.79, 0.89) | 0.66 (0.60, 0.71) |
| 707.2 | Chronic ulcer of leg or foot | dermatologic | 0.80 (0.76, 0.85) | 0.70 (0.65, 0.75) | 0.84 (0.79, 0.88) | 0.73 (0.67, 0.79) |
| 276.4 | Acid-base balance disorder | endocrine/metabolic | 0.77 (0.72, 0.82) | 0.67 (0.61, 0.72) | 0.79 (0.73, 0.85) | 0.67 (0.60, 0.74) |
| 300.9 | Posttraumatic stress disorder | mental disorders | 0.75 (0.70, 0.79) | 0.64 (0.59, 0.69) | 0.82 (0.76, 0.87) | 0.63 (0.56, 0.69) |
| 315.2 | Speech and language disorder | mental disorders | 0.81 (0.74, 0.87) | 0.73 (0.64, 0.82) | 0.83 (0.74, 0.90) | 0.71 (0.60, 0.83) |
| 569.2 | Gastrointestinal complications | digestive | 0.76 (0.70, 0.81) | 0.66 (0.59, 0.72) | 0.79 (0.71, 0.87) | 0.65 (0.57, 0.73) |
| 426.8 | Other cardiac conduction disorders | circulatory system | 0.79 (0.75, 0.82) | 0.71 (0.66, 0.75) | 0.82 (0.77, 0.87) | 0.69 (0.62, 0.76) |
| 458.2 | Iatrogenic hypotension | circulatory system | 0.76 (0.68, 0.83) | 0.67 (0.60, 0.74) | 0.77 (0.65, 0.87) | 0.67 (0.56, 0.77) |
| 250.7 | Diabetic retinopathy | endocrine/metabolic | 0.81 (0.77, 0.85) | 0.74 (0.69, 0.79) | 0.84 (0.79, 0.89) | 0.76 (0.69, 0.82) |
| 276.11 | Hyperosmolality and/or hypernatremia | endocrine/metabolic | 0.76 (0.66, 0.86) | 0.68 (0.59, 0.76) | 0.77 (0.59, 0.94) | 0.74 (0.62, 0.85) |
| 264.0 | Lack of normal physiological development | endocrine/metabolic | 0.77 (0.67, 0.86) | 0.69 (0.55, 0.81) | 0.72 (0.60, 0.84) | 0.63 (0.48, 0.77) |
| 440.22 | Atherosclerosis of native arteries of the extremities with intermittent claudication | circulatory system | 0.80 (0.73, 0.86) | 0.74 (0.65, 0.81) | 0.82 (0.74, 0.89) | 0.74 (0.65, 0.83) |
| 274.11 | Gouty arthropathy | endocrine/metabolic | 0.76 (0.71, 0.81) | 0.68 (0.63, 0.73) | 0.77 (0.70, 0.84) | 0.67 (0.60, 0.74) |
| 509.1 | Respiratory failure | respiratory | 0.77 (0.73, 0.80) | 0.69 (0.64, 0.73) | 0.76 (0.71, 0.82) | 0.68 (0.62, 0.74) |
| 260.2 | severe protein-calorie malnutrition | endocrine/metabolic | 0.79 (0.74, 0.83) | 0.71 (0.67, 0.76) | 0.83 (0.75, 0.90) | 0.72 (0.64, 0.80) |
| 550.4 | Umbilical hernia | digestive | 0.79 (0.74, 0.83) | 0.72 (0.64, 0.79) | 0.84 (0.78, 0.90) | 0.75 (0.66, 0.83) |
| 513.8 | Disorders of diaphragm | respiratory | 0.75 (0.65, 0.84) | 0.68 (0.57, 0.77) | 0.73 (0.58, 0.86) | 0.63 (0.48, 0.79) |
| 260.6 | Anorexia | endocrine/metabolic | 0.76 (0.71, 0.80) | 0.68 (0.62, 0.74) | 0.84 (0.79, 0.88) | 0.68 (0.58, 0.77) |
| 530.2 | Esophageal bleeding (varices/hemorrhage) | digestive | 0.76 (0.72, 0.79) | 0.69 (0.64, 0.74) | 0.87 (0.84, 0.90) | 0.69 (0.61, 0.75) |
| 440.2 | Atherosclerosis of the extremities | circulatory system | 0.81 (0.74, 0.86) | 0.75 (0.69, 0.81) | 0.84 (0.75, 0.90) | 0.78 (0.71, 0.85) |
| 411.9 | Other acute and subacute forms of ischemic heart disease | circulatory system | 0.78 (0.72, 0.85) | 0.72 (0.66, 0.79) | 0.79 (0.71, 0.88) | 0.75 (0.68, 0.82) |
| 227.0 | Benign neoplasm of other endocrine glands and related structures | neoplasms | 0.75 (0.66, 0.84) | 0.69 (0.59, 0.77) | 0.82 (0.69, 0.93) | 0.72 (0.59, 0.83) |
| 250.1 | Type 1 diabetes | endocrine/metabolic | 0.75 (0.69, 0.81) | 0.68 (0.62, 0.75) | 0.77 (0.70, 0.82) | 0.70 (0.64, 0.78) |
| 285.1 | Acute posthemorrhagic anemia | hematopoietic | 0.78 (0.75, 0.81) | 0.72 (0.68, 0.75) | 0.83 (0.79, 0.87) | 0.73 (0.68, 0.78) |
| 415.1 | Acute pulmonary heart disease | circulatory system | 0.80 (0.75, 0.85) | 0.75 (0.69, 0.81) | 0.84 (0.77, 0.90) | 0.76 (0.69, 0.83) |
| 348.8 | Encephalopathy, not elsewhere classified | neurological | 0.77 (0.70, 0.83) | 0.71 (0.64, 0.77) | 0.78 (0.70, 0.86) | 0.70 (0.62, 0.78) |
| 427.11 | Paroxysmal supraventricular tachycardia | circulatory system | 0.79 (0.76, 0.81) | 0.73 (0.70, 0.76) | 0.87 (0.84, 0.90) | 0.75 (0.70, 0.79) |
| 427.12 | Paroxysmal ventricular tachycardia | circulatory system | 0.78 (0.74, 0.82) | 0.72 (0.67, 0.77) | 0.84 (0.79, 0.88) | 0.72 (0.65, 0.78) |
| 428.2 | Heart failure NOS | circulatory system | 0.77 (0.70, 0.84) | 0.71 (0.64, 0.78) | 0.81 (0.74, 0.88) | 0.72 (0.64, 0.80) |
| 427.1 | Paroxysmal tachycardia, unspecified | circulatory system | 0.78 (0.76, 0.81) | 0.73 (0.71, 0.76) | 0.87 (0.84, 0.89) | 0.75 (0.71, 0.79) |
| 501.0 | Pneumonitis due to inhalation of food or vomitus | respiratory | 0.75 (0.70, 0.80) | 0.69 (0.62, 0.76) | 0.75 (0.69, 0.81) | 0.66 (0.57, 0.74) |
| 276.13 | Hyperpotassemia | endocrine/metabolic | 0.75 (0.71, 0.80) | 0.70 (0.65, 0.74) | 0.79 (0.73, 0.85) | 0.69 (0.63, 0.74) |
| 459.9 | Circulatory disease NEC | circulatory system | 0.77 (0.74, 0.79) | 0.71 (0.68, 0.75) | 0.84 (0.80, 0.87) | 0.74 (0.69, 0.78) |
| 574.2 | Calculus of bile duct | digestive | 0.76 (0.70, 0.80) | 0.70 (0.63, 0.78) | 0.79 (0.72, 0.85) | 0.72 (0.64, 0.80) |

**Supplementary Table 15.**
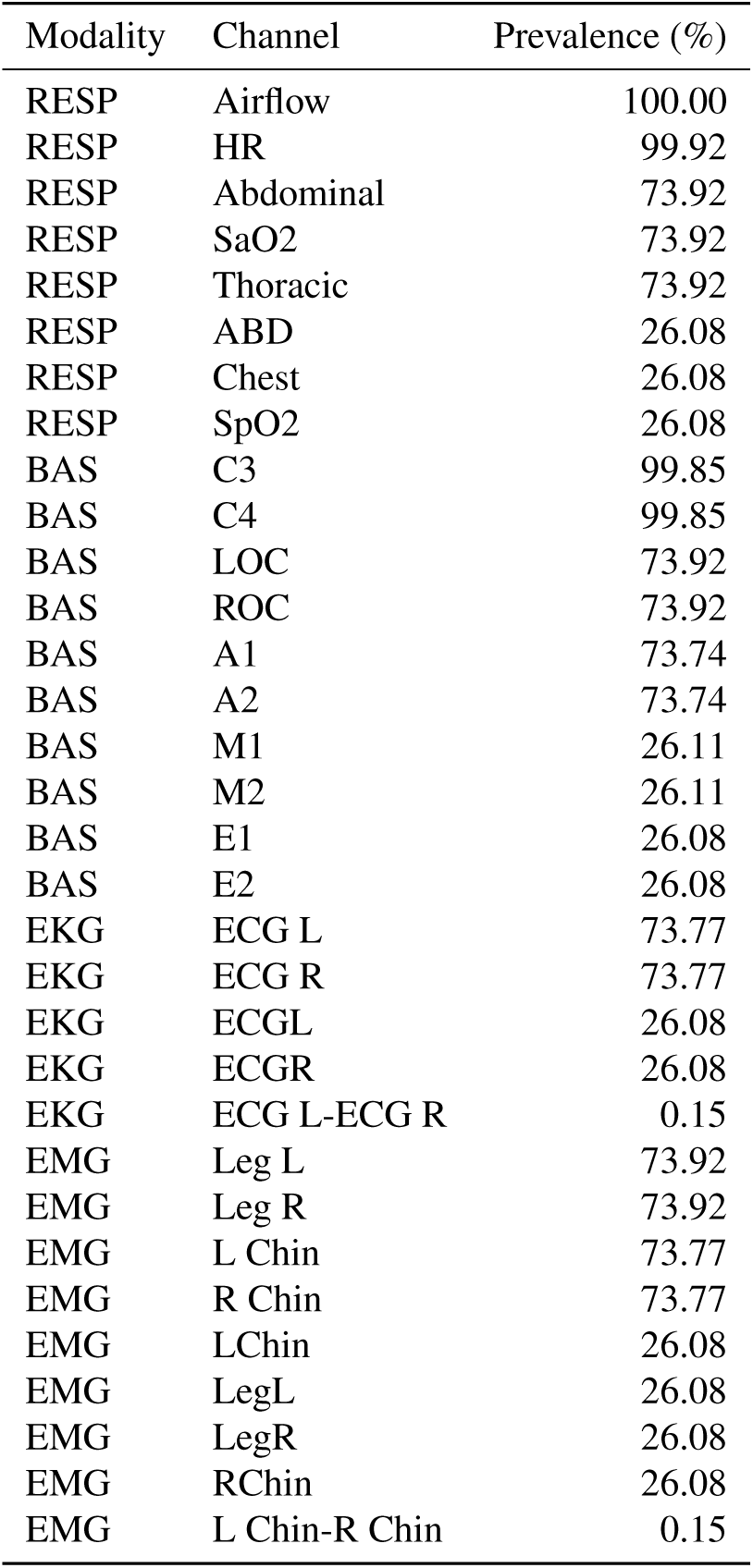
Channels and prevalence percentages by modality for MROS.

**Supplementary Table 16.**
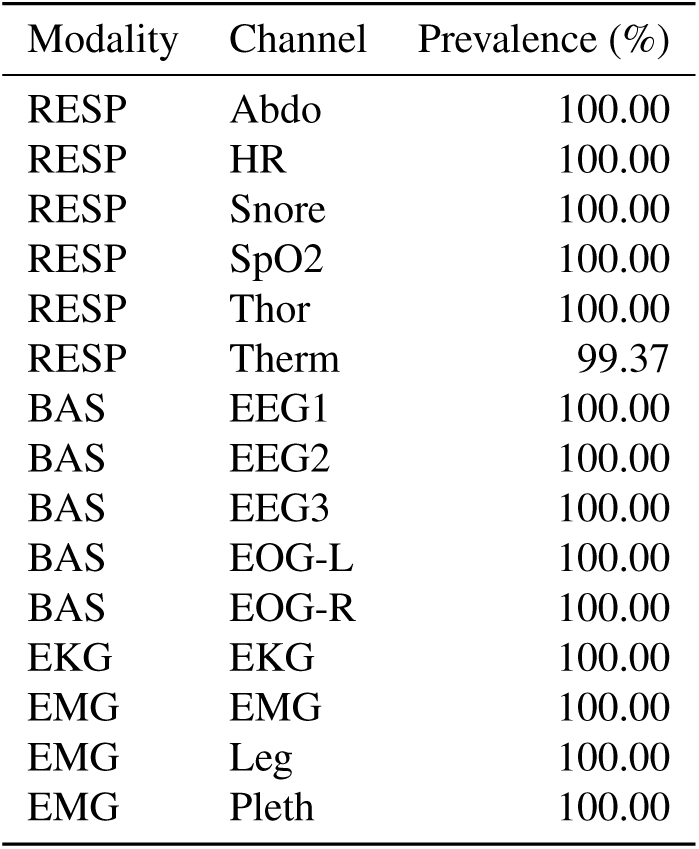
Channels and prevalence percentages by modality for MESA.

**Supplementary Table 17.** Channels and prevalence percentages by modality for SHHS.

| Modality | Channel | Prevalence (%) |
| --- | --- | --- |
| RESP | ABDO RES | 100.00 |
| RESP | SaO2 | 100.00 |
| RESP | THOR RES | 100.00 |
| RESP | AIRFLOW | 75.94 |
| RESP | H.R. | 68.69 |
| RESP | NEW AIR | 46.64 |
| RESP | New Air | 9.43 |
| RESP | NEWAIR | 4.14 |
| RESP | AIRFLOW-0 | 1.08 |
| RESP | AIRFLOW-1 | 1.08 |
| EKG | ECG | 100.00 |
| BAS | EEG | 100.00 |
| BAS | EOG(L) | 100.00 |
| BAS | EOG(R) | 100.00 |
| BAS | EEG(sec) | 97.95 |
| BAS | EEG 2 | 1.15 |
| BAS | EEG2 | 0.65 |
| BAS | EEG sec | 0.15 |
| BAS | EEG(SEC) | 0.09 |
| EMG | EMG | 100.00 |
| EMG | LEG(L) | 0.02 |
| EMG | LEG(R) | 0.02 |

**Supplementary Table 18.**
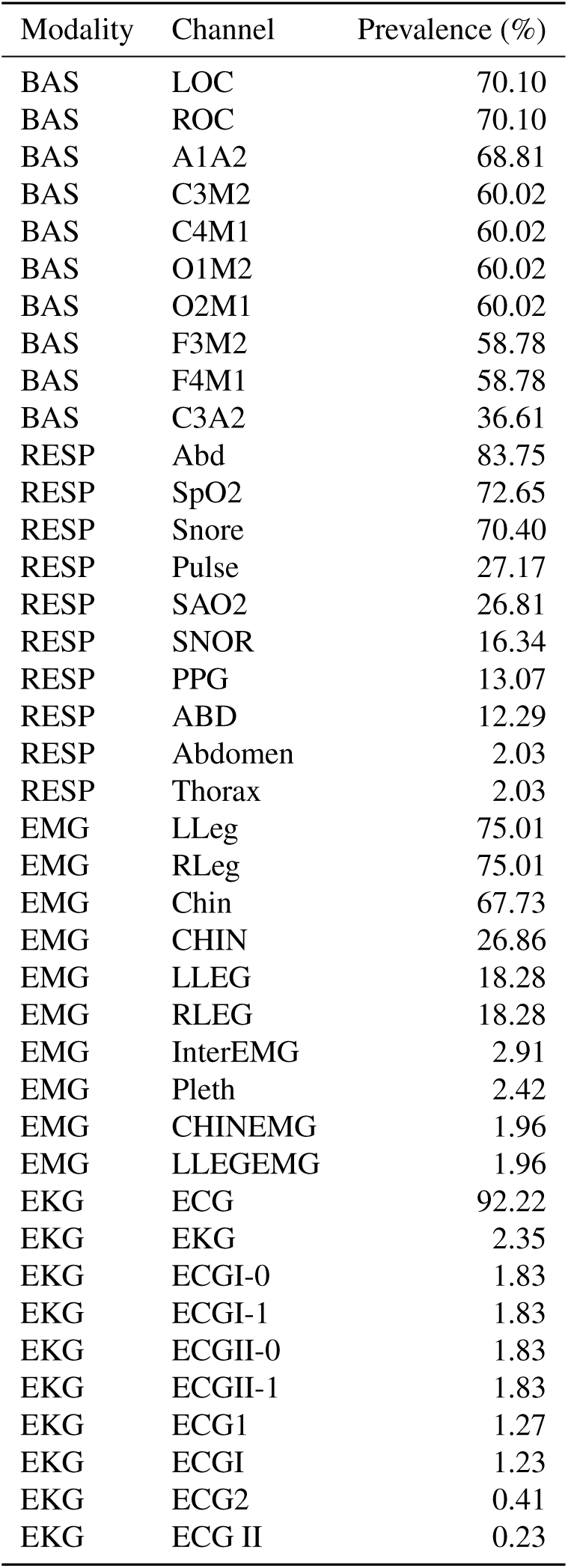
Channels and prevalence percentages by modality for BioSerenity.

| Modality | Channel | Prevalence (%) |
| --- | --- | --- |
| BAS | LOC | 70.10 |
| BAS | ROC | 70.10 |
| BAS | A1A2 | 68.81 |
| BAS | C3M2 | 60.02 |
| BAS | C4M1 | 60.02 |
| BAS | O1M2 | 60.02 |
| BAS | O2M1 | 60.02 |
| BAS | F3M2 | 58.78 |
| BAS | F4M1 | 58.78 |
| BAS | C3A2 | 36.61 |
| RESP | Abd | 83.75 |
| RESP | SpO2 | 72.65 |
| RESP | Snore | 70.40 |
| RESP | Pulse | 27.17 |
| RESP | SAO2 | 26.81 |
| RESP | SNOR | 16.34 |
| RESP | PPG | 13.07 |
| RESP | ABD | 12.29 |
| RESP | Abdomen | 2.03 |
| RESP | Thorax | 2.03 |
| EMG | LLeg | 75.01 |
| EMG | RLeg | 75.01 |
| EMG | Chin | 67.73 |
| EMG | CHIN | 26.86 |
| EMG | LLEG | 18.28 |
| EMG | RLEG | 18.28 |
| EMG | InterEMG | 2.91 |
| EMG | Pleth | 2.42 |
| EMG | CHINEMG | 1.96 |
| EMG | LLEGEMG | 1.96 |
| EKG | ECG | 92.22 |
| EKG | EKG | 2.35 |
| EKG | ECGI-0 | 1.83 |
| EKG | ECGI-1 | 1.83 |
| EKG | ECGII-0 | 1.83 |
| EKG | ECGII-1 | 1.83 |
| EKG | ECG1 | 1.27 |
| EKG | ECGI | 1.23 |
| EKG | ECG2 | 0.41 |
| EKG | ECG II | 0.23 |

**Supplementary Table 19.**
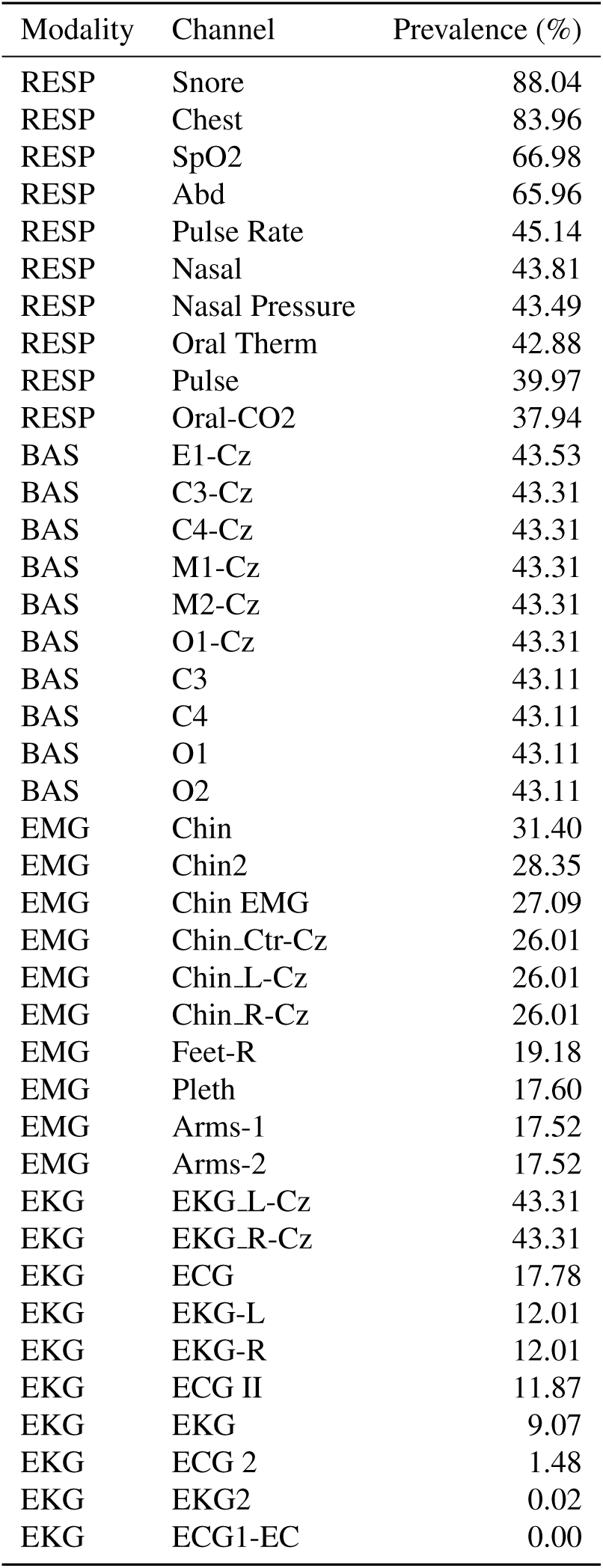
Channels and prevalence percentages by modality for SSC.

## Notes

### Competing Interest Statement

The authors have declared no competing interest.

### Funding Statement

Rahul Thapa is supported by the Knight-Hennessy Scholars funding. Drs. Mignot and Westover are supported by a grant from the National Heart, Lung, and Blood Institute of the NIH (R01HL161253). Dr. Zou is supported by funding from the Chan-Zuckerberg Biohub.

### Author Declarations

Ethics committee/IRB of Stanford University gave ethical approval for this work (Protocol Number: 69873)

### Summary of Updates

We have added additional SleepFM model version and extended its comparison with additional baseline models.

